# Health economic evaluation of strategies to eliminate *gambiense* human African trypanosomiasis in the Mandoul disease focus of Chad

**DOI:** 10.1101/2022.07.25.22278030

**Authors:** Marina Antillon, Ching-I Huang, Samuel A. Sutherland, Ronald E. Crump, Paul R. Bessell, Alexandra P.M. Shaw, Iñaki Tirados, Albert Picado, Sylvain Biéler, Paul E. Brown, Philippe Solano, Severin Mbainda, Justin Darnas, Xia Wang-Steverding, Emily H. Crowley, Mallaye Peka, Fabrizio Tediosi, Kat S. Rock

## Abstract

Human African trypanosomiasis, caused by the gambiense subspecies of *Trypanosoma brucei* (gHAT), is a deadly parasitic disease transmitted by tsetse. Partners from around the world have stepped up efforts to eliminate the disease, and the Chadian government have had a particular focus on the previously high-prevalence setting of Mandoul. In this study, we evaluate the economic efficiency of the intensified strategies that were put in place from 2014 aimed at interrupting transmission of gHAT, and we make recommendations on the best way forward based on both epidemiological projections and cost-effectiveness. In our analysis we use a dynamic transmission model fit to epidemiological data from Mandoul to evaluate the cost-effectiveness of combinations of active screening, improved passive screening (defined as an expansion of the number of health posts capable of screening for gHAT), and vector control activities (the deployment of Tiny Targets). For cost-effectiveness analyses, our primary outcome is disease burden, denominated in disability-adjusted life-years (DALYs), and costs, denominated in 2020 US$. Although active and passive screening have enabled more rapid diagnosis and accessible treatment in Mandoul, the addition of vector control provided good value-for-money (at less than $750/DALY averted) and substantially increased the probability of reaching the 2030 elimination target for gHAT as set by the World Health Organization. Our transmission modelling and economic evaluations suggests that the gains have been made could be maintained by robust passive screening, and the inclusion of active screening and vector control strategies could be considered for other foci in the country with active transmission. Our analysis speaks to comparative efficiency, and it does not take into account all possible considerations; for instance, any cessation of on-going active screening should first consider that strong surveillance activities will be critical to verify elimination of transmission and to protect against the possible importation of infection from neighbouring endemic foci.

**Author summary:** In a drive to eliminate human African trypanosomiasis (gHAT or sleeping sickness) from Chad following a peak in 2002, the National Sleeping Sickness Control programme and their partners focused on making substantial changes to interventions within the high prevalence setting of Mandoul. These included the use of vector control starting in 2014 and improved screening in health facilities starting in 2015. To explore whether these interventions were an efficient use of resources we carried out a retrospective analysis using a dynamic transmission model fit to epidemiological data from Mandoul combined with a cost model. Our analysis indicated that active and passive screening have enabled more rapid diagnosis and accessible treatment in Mandoul compared to less ambitious interventions, and furthermore the addition of vector control provided good value-for-money and substantially increased the probability of reaching the 2030 elimination of transmission target for gHAT set by the World Health Organization. Looking forwards, our prospective analysis also considers the health economics of future strategies and, as part of this, concludes that the scaleback of vertical interventions appears cost-effective if passive screening remains robust in Mandoul. This could therefore enable shifting of resources to tackle other remaining foci in Chad.

## Introduction

In 1993, after forty years without any case reports of *gambiense* human African trypanosomiasis (gHAT) in the Mandoul region of Chad, health care personnel at the newly opened Catholic Mission Health Centre diagnosed a patient with gHAT, setting off new efforts against the disease in the region [1]. Commonly referred to as “sleeping sickness”, the disease is caused by the *Trypanosoma brucei gambiense* parasite, and is characterized by undifferentiated symptoms in the initial blood-borne stage and neurological symptoms in the second stage, which are generally (but not always) fatal if untreated [2, 3]. In the first year of renewed reports in Mandoul, 212 cases were diagnosed in total, comprising most of the gHAT cases in Chad, followed by 2,291 more cases during the following fourteen years [1]. The detection of disease in the Mandoul focus paralleled an uptick in reported cases throughout the continent. The epidemic lasted more than a decade, during which 3,316 cases were reported between 1990 and 2004 in Chad [4] with 37,000 cases reported in a single year continent-wide at its peak in 1998. It is likely that many more infections went undiagnosed due to historically different constraints that limited the screening activities, with untreated cases almost invariably culminating in the death of the infected person [5–7].

In many respects gHAT presents a number of unique challenges compared to most other infections targeted for control or elimination: there is no vaccine against the disease; the treatment, which causes a number of side-effects, cannot be used for mass-drug administration [8]; until 2020, a painful lumbar puncture for disease staging was required to assign the correct treatment [2]; and control of the vector, the tsetse (*Glossina* spp.), requires alternative interventions to those popular for controlling mosquitoes [9]. Despite this, and after years of investment in screening campaigns, the appropriate diagnosis, and referral for treatment, the last decade has been characterized by optimism. In 2012 the World Health Organization (WHO) marked the disease for elimination as a public health problem by 2020 [10], and for elimination of transmission by 2030 [11]. The 2020 goal was defined for the continent as a 90% reduction in areas at moderate or high risk (defined as more than one new reported case per year per 10,000 people) compared to the 2000–2004 baseline and fewer than 2000 annual global cases [12]. The within-country indicator was given as less than one new reported case per year per 10,000 people per year in each health district by 2020 [10]. The 2030 goal was reported as a complete cessation of transmission to humans by 2030 [13]. By 2017, there were fewer than 2000 reported cases continent-wide, outpacing the case reduction indicator set for the 2020 goal, although the reduction in areas at risk slightly missed the target (achieving 83% rather than 90%) [14].

Within Chad there are three foci that are of concern: Maro and Moissala, close to the border of the Central African Republic, and Mandoul [15]. Two additional foci, Gore and Tapol – that have historically had cases – appear to no longer be active [16]. Starting in 2014, after half a decade of low but unremitting case detection, intensified screening in health facilities and a vector control project was put in place by the national gHAT control programme (Programme National de lutte contre la Trypanosomiase Humaine Africaine; PNLTHA-Chad) and international non-profit and academic partners [17–19]. Previous publications detail the impacts of these efforts on case reporting and transmission [20, 21], however, here we will focus on the economic efficiency of these interventions in Mandoul. Two other studies have looked at the costs of screening and vector control separately, but not in concert and so would miss the synergies possible in simultaneous interventions to halt transmission [19, 22]. We set out to perform a retrospective analysis for the Mandoul focus in which we look at what was done from 2014 compared to three less ambitious interventions. We examine what would have been the health economic implications if lesser strategies had been performed, and we test whether the introduction of fexinidazole in 2014 would have changed that selection. Our subsequent prospective analysis, also considers the health economic implications of what could be done against gHAT going forward from 2023 for Mandoul.

## Methods

### Setting and the historical control of gHAT

The focus of Mandoul (ca. 8.12°N, 17.11°E) spans 840 km^2^; approximately the land area of Copenhagen or Austin, Texas. It contains no paved roads and is characterized by a gallery forest along a marshland. The focus spans five cantons – Bodo, Beboto, Dilingala, Koldaga, and Bekourou – across two provinces – Logone Oriental and Mandoul (see Fig A in Supplementary Information S1 Text) [19, 20]. The focus is bisected by Niaméte river on the southern part with temporary tributaries during the wet season from May to September [23]. The river gives way to marshland towards the north.

As of 2013, the focus was dotted by 114 settlements and contained about 38,674 people of the Sara ethnic group: 1,029 people lived in 22 encampments of fewer than 100 people, 17,626 lived in hamlets of between 100–500 people, and 20,016 people lived in 27 villages of more than 500 people [20]. During the dry season, the northern marshland dries out and is populated by pastoral Bororo nomads [23]. The population participates in subsistence farming and lives in traditional huts with no modern amenities. Throughout the focus, the marshy waterway is traversed via several cross-ways, used especially during market days when people in the eastern side of the focus cross to the west, where the hamlet of Bodo is located within Logone Oriental Province. In general, this focus is relatively isolated compared to Moissala and Maro; Moissala is contiguous with the Maitikolo focus of the Central African Republic (CAR) and often receives refugees while Maro sits along the CAR border in the east of the country [24].

Screening, diagnosis and treatment for gHAT in Chad is coordinated by PNLTHA-Chad since the early 1990’s, which has headquarters in Moundou, about 150km west of the Mandoul focus [1, 23]. Until 2014, passive diagnosis of cases was only available in the Catholic Mission Hospital in the town of Bodo, which is the main service centre and transport hub of the focus. Active screening of at-risk villages was also conducted during the period but the temporal coverage of active screening varied contingent on available funding and security conditions.

Historically, PNLTHA-Chad has been funded with help from the *l’Organization de Coordination pour la Lutte contre les Endémies en Afrique centrale* (OCEAC) and the WHO, while diagnosis and treatment at the Catholic Mission Hospital is financed by user fees and private charity funds. Treatment medications have been donated via WHO to PNLTHA since 2002. In addition to the continued rounds of vehicle-based active screening that were funded by WHO, medical screening activities have been expanded since 2015 with funding from the Swiss Agency for Development and Cooperation (SDC), Germany’s Kreditanstalt für Wiederaufbau (KfW) and the BMGF, through Foundation for Innovative and Novel Diagnostics (FIND), including rapid-diagnostic tests (RDTs) in a number of healthcare facilities, additional laboratory capacity for parasitological confirmation of cases, and the expansion of active screening with additional traditional teams as well as “mini-teams” on motorcycles using RDTs. Moreover, since 2013, vector control (VC) has been supported by the Bill and Melinda Gates Foundation (BMGF) with a first deployment of Tiny Targets in 2014 following the baseline surveys in 2013. Since October 2016 the medical and entomological control activities have been integrated under the banner of the BMGF funded Trypa-NO! Partnership [17].

### Strategies

#### Strategy changes between 2014–2016

Prior to 2015, active screening (AS) was based on 12-person, 2-vehicle teams that would travel village-to-village while passive screening (PS) and parasitological diagnosis were only available at the Catholic Mission Hospital [22, 23]. Starting in 2015, the Catholic Mission Hospital was outfitted with loop-mediated isothermal amplification (LAMP), and a second hospital, the Bodo district hospital was outfitted to be able to diagnose, confirm, stage, and treat gHAT cases. Moreover, eight clinics were stocked with RDTs for screening, which then referred RDT-positive, suspected cases to the two hospitals for parasitological confirmation. In the following years, more facilities would be stocked, culminating in 31 clinics screening suspected cases in 2018. Eleven clinics were no longer stocked in 2019 in areas where no cases were detected (for more details, see Table C in Supplementary Information S1 Text).

Starting in 2014, AS based on mobile units was complemented by smaller, “mini-mobile” teams riding motorbikes and administering diagnostic algorithms with RDTs [22, 23] (see Table B in Supplementary Information S1 Text). While traditional teams confirmed cases onsite via microscopy exams of blood and cerebrospinal fluid, mini-mobile teams referred RDT+ patients to the two hospitals for confirmation with parasitology or Loop-mediated Isothermal Amplification (LAMP) and treatment [25]. The screened villages have an average population of around 800 and 40–50% of the population present for screening by mobile teams and because the structure of the teams is flexible, the team can adapt to larger villages or higher participation rates (see Tables A-B in Supplementary Information S1 Text). Transportation to the focus is limited during the rainy season of May to September due to the poor state of the roads [1, 23]. Furthermore, screening during the dry season is preferable as the population is less busy with agriculture [22]. Transportation is not much easier within the focus, and motorcycle teams have been designed to more easily reach villages and to provide transportation to suspected cases [23, 26].

To complement medical activities, in 2014 annual deployment of VC was initiated along the river and the marshland to control the tsetse population and interrupt transmission [20]. The deployment of these Tiny Targets – which attract tsetse and provide a lethal dose of insecticide through their impregnated mesh – has been very effective at reducing the local fly population density in Mandoul, with an estimated reduction of over 99% in only four months [20, 21] which has been sustained since then. The isolated nature of the Mandoul focus is believed to be one key reason for such a dramatic impact, although reductions of over 80% after one year have been found in various other regions across Africa using the Tiny Target approach [27–30]. High levels of vector reduction result in substantial disruption to the transmission cycle without the need for complete elimination of the tsetse. This vector intervention in Mandoul is further described by Rayaisse et al. [19].

#### Counterfactual strategies for retrospective analysis and prospective analysis

The intervention strategies for Mandoul are found in Table 1 and illustrated in Fig 2 for both the retrospective and prospective analysis. In the retrospective analysis, we considered outcome of the strategy that was actually implemented, with improved PS and VC (*Imp. PS & VC*) to compare against less ambitious strategies. Alternative strategies included the interventions at levels present before 2014 (*Pre-2014*), as well as strategies that either only improved PS (*Imp. PS*) or only implemented VC (*Addition of VC*) alongside AS. In all retrospective strategies, the same amount of AS took place and the passive detection rate was at least as much as before 2014.

**Table 1.** Strategies against gHAT in the Mandoul focus. Active screening (AS) coverage in the retrospective analysis is actual coverage in 2014–2019, and the recent mean coverage (2015–2019) thereafter – this is denoted as default coverage (ASd). Coverage in the prospective analysis is the recent mean (*Mean AS*) or the historical maximum for 2000–2019 (*Max AS*), which includes enhanced coverage (ASe). For passive screening (PS) PSd refers to default coverage, consistent with what was present before 2014 and PSe refers to enhanced coverage due to the clinics that were newly stocked with RDTs starting in 2015. Annually deployed vector control (VC) with around a 99% decrease in the population of tsetse in the first 4 months [21]. Treatment is offered to all cases detected. Active screening algorithm specificity under *Mean AS & VC (a)* is 99.93% and for the strategies *Mean AS & VC (b), Mean AS* and *Max AS* it is 100%. *Stop 2023 (No AS or VC)* signifies that AS and VC do not occur from 2023 onward. No P+/S+ indicates that both serological and confirmed cases must decline to 0 before scaleback. No P+ indicates that only cases confirmed by parasitology or trypanolysis must decline to 0 before scaleback.

|  | Component Interventions |  |  |  |  | Scale-back criteria |  |
| --- | --- | --- | --- | --- | --- | --- | --- |
|  | ASd | ASe | PSd | PSe | VC | AS & VC | PS |
| *Retrospective analysis* |  |  |  |  |  |  |  |
| Pre-2014 | ✓ |  | ✓ |  |  | 3y no P+ | - |
| Imp. PS | ✓ |  | ✓ | ✓ |  | 3y no P+ | 5y no P+ |
| Addition of VC | ✓ |  | ✓ |  | ✓ | 3y no P+ | - |
| Imp. PS & VC | ✓ |  | ✓ | ✓ | ✓ | 3y no P+ | 5y no P+ |
| *Prospective analysis* |  |  |  |  |  |  |  |
| Mean AS & VC (a) | ✓ |  | ✓ | ✓ | ✓ | 3y no S+ or P+ | 5y no S+ or P+ |
| Mean AS & VC (b) | ✓ |  | ✓ | ✓ | ✓ | 3y no P+ | 5y no P+ |
| Mean AS | ✓ |  | ✓ | ✓ |  | 3y no P+ | 5y no P+ |
| Max AS | ✓ | ✓ | ✓ | ✓ |  | 3y no P+ | 5y no P+ |
| Stop 2023 (No AS or VC) |  |  | ✓ | ✓ |  | - | 5y no P+ |

**Table 2.** Model Parameters. For further details and sources, see S4 Text. CIs: confidence intervals. AS & PS: active and passive screening, respectively, VC: vector control, PNLTHA: Programme de Lutte contre la Trypanosomie Humaine, NECT: nifurtimox-eflornithine combination therapy, CATT: card agglutination test for trypanosomiasis, S1 & S2: stage 1 & 2 disease, DALYs: disability-adjusted life-years, SAE: severe adverse events, RDT: Rapid Diagnostic Test

| Variable Description | Statistical Distribution | Summary Mean (95% CIs) |
| --- | --- | --- |
| **Screening parameters** |  |  |
| Population | Fixed value | 41,000 |
| PS: coverage of the population per facility | Fixed value varying by year. | See S1 Text Table C & section S1.2.2 Prospectively, 100. |
| PS: number of facilities | Fixed value varying by year. | See S1 Text Table C & section S1.2.2. Prospectively, 22. |
| AS: coverage | Fixed value varying by year. | See S1 Text Table A-B & section S1.2.2. |
| CATT algorithm: diagnostic specificity | Beta(31, 2) | 0.94 (0.84-0.99) |
| RDT algorithm: diagnostic sensitivity | Beta(230, 1) | 1.00 (0.98-1.00) |
| RDT algorithm: diagnostic specificity | Beta(226, 31) | 0.88 (0.84-0.92) |
| CATT algorithm: wastage during AS | Beta(8, 92) | 0.08 (0.03-0.14) |
| RDT algorithm: wastage during PS | Beta(1, 99) | 0.01 (<0.01-0.04) |
| **Screening cost parameters** |  |  |
| AS: capital costs of a traditional team | Gamma(200,000, 0.02) | 3192 (3178-3206) |
| AS: fixed management costs of traditional teams | Gamma(8.475-2167) | 18,436(8047-32,580) |
| AS: capital costs of a mini-team | Gamma(8.475, 1000.81) | 8490 (3823-14,910) |
| AS: fixed management costs of a mini-team | Gamma(70.05, 91.56) | 6409 (4975-7989) |
| CATT algorithm: cost per test used | Gamma(22.87, 0.02) | 0.45 (0.29-0.67) |
| Staging: lumbar puncture & lab exam | Gamma(2.42, 3.66) | 8.90 (1.45-23.20) |
| Confirmation: microscopy | Gamma(8.475, 1.27) | 10.64 (4.78-18.68) |
| RDT algorithm: costs per test used | Gamma(8.475, 0.19) | 1.60 (0.71-2.83) |
| PS: capital costs of a facility | Gamma(8.475, 216.24) | 1,822 (811-3208) |
| PS: capital costs of a facility that screens with RDT | Gamma(8.475, 31.97) | 259 (122-484) |
| PS: fixed recurrent management costs (whole focus) | Gamma(32.47, 12.18) | 396 (272-545) |
| **Treatment parameters** |  |  |
| Proportion of cases age<6 | Beta(152.53, 2427.9) | 0.06 (0.05-0.07) |
| Proportion of cases weight<35 kg & age>6 | Beta(8.3, 359.6) | 0.02 (<0.01-0.04) |
| Proportion of S2 cases that are severe | Beta(76.93, 44.87) | 0.63 (0.54-0.72) |
| Age of death from infection | Gamma(148, 0.18) | 26.63 (22.41-31.08) |
| Length of hospital stay: NECT | Fixed value | 10 |
| Length of hospital stay: fexinidazole | Fixed value | 10 |
| Pr. of relapse: pentamidine | Beta(50.3, 665.48) | 0.07 (0.05-0.09) |
| Pr. of relapse: NECT | Beta(15.87, 378.55) | 0.05 (0.02-0.08) |
| Pr. of relapse: fexinidazole | Beta(9.49, 496.54) | 0.02 (<0.01-0.03) |
| SAE: pentamidine treatment | Beta(1.43, 551.42) | 0.002 (<0.01-0.01) |
| SAE: NECT treatment | Beta(40.88, 367.8) | 0.05 (0.03-0.08) |
| SAE: fexinidazole treatment | Beta(3, 261) | 0.01 (<0.01-0.03) |
| Days lost to disability due to S1 disease | Gamma(21.89, 26.07) | 569.21 (355.50-831.29) |
| Days lost to disability due to S2 disease | Gamma(22.18, 12.38) | 275.57 (172.46-401.20) |
| Days lost to disability due to SAE | Gamma(1.22, 2.38) | 2.96 (0.14-9.99) |
| **Treatment cost parameters** |  |  |
| Hospital stay: cost per day | Gamma(5.45, 1.76) | 9.51 (3.27-19.34) |
| Outpatient consultation: cost | Uniform(2.48, 0.79) | 1.96 (0.32-5.06) |
| Course of pentamidine: cost | Fixed value | 54 |
| Course of NECT: cost | Fixed value | 360 |
| Course of fexinidazole: cost | Fixed value | 50 |
| Drug delivery mark-up | Beta(45, 55) | 0.45 (0.35-0.55) |
| **Vector control parameters** |  |  |
| Replacement rate of targets per year | Fixed value | 1 |
| **Vector cost parameters** |  |  |
| Deployment cost per year | Gamma(8.47, 81.32) | 68,847 (30,307-123,000) |
| **DALY parameters** |  |  |
| Average years lost at age of death in Chad | Linear interpolation of life expectancy at 20-35 in Chad | 45.4 (41.4-49.9) |
| Average age of death of a HAT case | Gamma(148,0.18) | 26.6 (22.4-31.0) |
| Disability weights: S1 disease | Beta(22.96, 147.21) | 0.14 (0.09-0.19) |
| Disability weights: S2 disease | Beta(18.37, 15.63) | 0.54 (0.37-0.70) |
| Disability weights: SAE | Uniform(0.04, 0.11) | 0.08 (0.04-0.11) |

**Fig 1.**
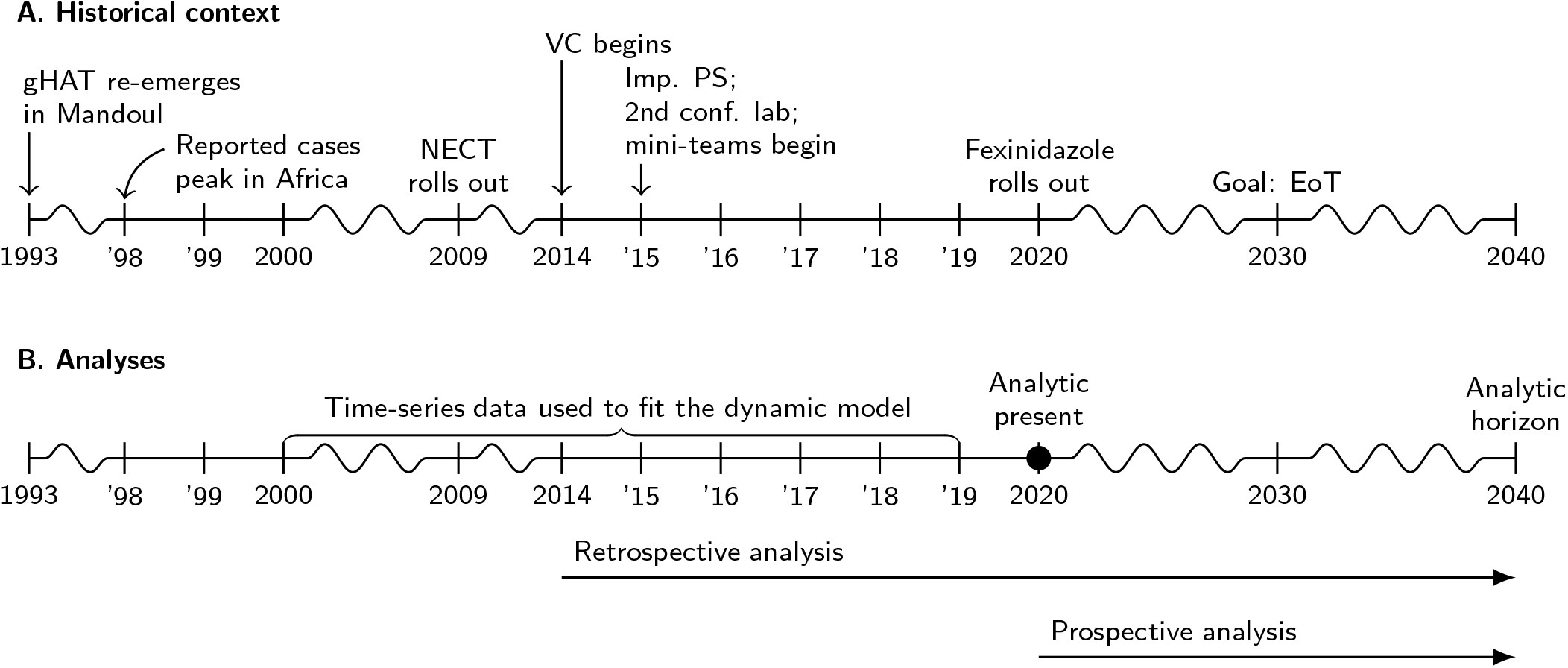
Timeline of gHAT epidemiology and analytic frame. A. Epidemiological information, start of new or improved interventions in Mandoul and the WHO 2030 goal. B. The data that were used to fit the model and the years corresponding to our retrospective and prospective analysis. NECT: Nifurtimox-eflornithine combination therapy, VC: vector control, Imp. PS: Improved passive screening, EoT: elimination of transmission.

**Fig 2.**
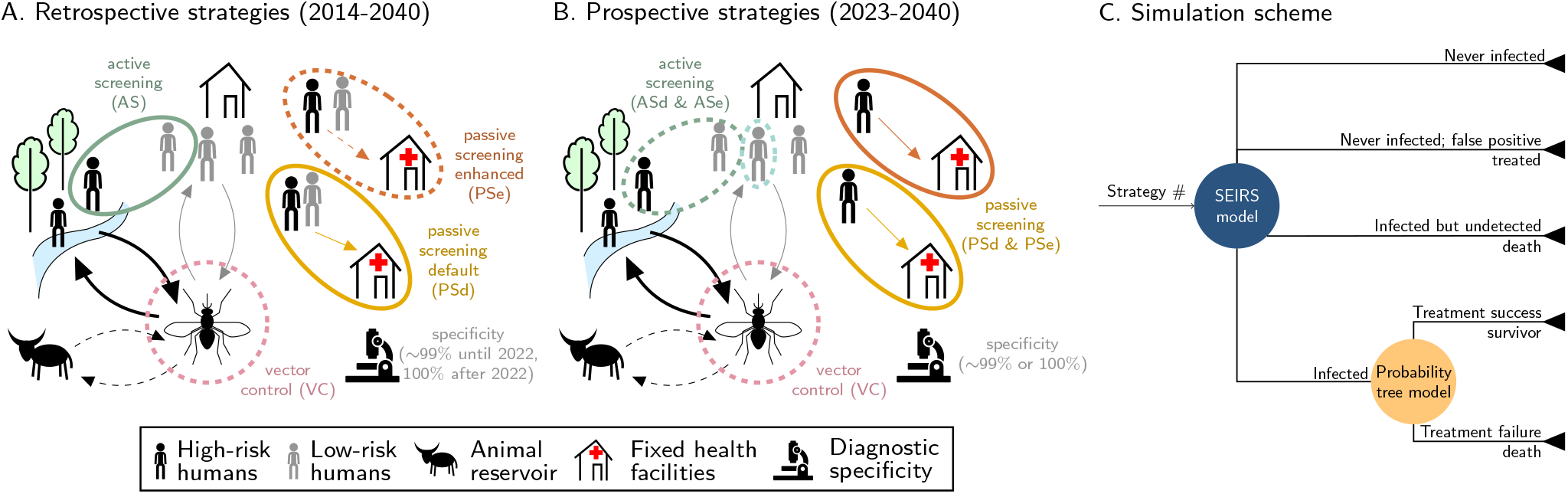
Schematic representation of the transmission models and the strategies in the (A) retrospective (2014 onward) and (B) prospective (2023 onward) analyses. Solid lines in color represent intervention components which are present across strategies (e.g. active screening (AS) in the retrospective analysis and passive screening in the prospective analysis), whereas dashed lines in color are for intervention components only present in some strategies (e.g. vector control). Arrows in black denote transmission routes which includes more transmission to high-risk individuals, and possible tsetse-animal-tsetse transmission pathways in some model variants (denoted by dashed black arrows). The imperfect specificity implies that there could be false positives that are treated. C shows the modeling scheme which combines the transmission model (SEIRS) outputs with the probability tree outcomes.

In the prospective analysis, we look to alternative strategies that can be instituted going forwards from 2023 based on the actual situation in 2020 (analytical present). Therefore all these simulations include the intensified strategies that took place from 2014 combining AS, the expanded network of PS, and VC. All prospective strategies simulate AS in 2021 and 2022 at the mean coverage of 2014–2019 (19,628 persons per year), and changes to strategies are assumed to take place from 2023.

#### Post-suppression phase

Suppression and post-elimination is defined here as the cessation of AS and VC after a pre-defined number of consecutive years of zero detected cases by any screening modality (AS or PS). After 2023, we simulated the impact of cessation of AS and VC if no cases are observed for a given period of time. For *Stop 2023 (No AS or VC)*, AS and VC stops immediately (it does not occur in 2023 onward). Under strategy *Mean AS & VC (a)*, AS and VC shut down when three years transpire of neither a serological positive (S+) nor a parasitologically confirmed (P+) case. For all other prospective strategies, AS and VC, when applicable, will shut down after three years of no detected P+ cases, even if S+ cases continue to be diagnosed and treated with fexinidazole. We rely on a realistic metric of disease suppression – either serological suspects or parasitologically confirmed cases – in order to simulate the risk and size of the resurgence if vertical interventions are ceased mistakenly early.

In the event that new P+ cases of either stage are detected by PS, an epidemiological investigation would take place to determine where the case was infected, if possible. In all strategies except for *Stop 2023 (No AS or VC)*, AS is assumed to be reactivated as reactive screening (RS) in the villages surrounding where the case was infected until there are no more P+ cases for one year. However, once VC stops it would not be restarted, even if more cases are found. The availability of RDT screening in health facilities (PS) is assumed to remain constant for five years after no P+ cases have been reported (and for five years after no S+ or P+ in the *Mean AS & VC (a)* strategy). The two hospitals providing screening, confirmation, and treatment are assumed not to rescind gHAT operations. However, no possibility of renewed expansion of RDT-stocked facilities was assumed. This is a reasonable assumption as a maximum of 0.1% of our simulations had further case reporting after five consecutive years with zero cases.

Lastly, Chadian samples are shipped to Burkina Faso for Trypanolysis for post-treatment confirmation of S+ samples. Since only a few dozen samples are sent each year and there is no documentation of the cost to do this, we have omitted Trypanolysis costs from the analysis.

### Transmission and treatment models

To evaluate the health outcomes of different strategies against gHAT, we employed a previously published and validated dynamic, transmission “susceptible-exposed-infected-recovered-susceptible (SEIRS)” model [21] coupled here with a probability tree model of treatment outcomes which are discussed in detail below.

#### Transmission model

We derived our simulations from an ensemble of models with compartments representing disease stages in humans and (possibly) non-human animals through tsetse transmission. The models are formalized through a system of ordinary differential equations (ODEs) and the ensemble is constituted of a sample of posterior predictions weighted by the model evidence for each structure. The model structure was originally developed by Rock et al. [31], modified for the Chadian context in Mahamat et al. [20], and was recently updated by Rock et al. [21]. Through numerical integration of the ODE system, we calculate the average expected dynamics. New infections and life-years lost in stage 1 and 2 are deterministic outputs. Chance events, such as case detection and deaths, were simulated by sampling from a beta-binomial distribution with a rate parameter equal to the incidence of infected individuals who reported to care and are diagnosed in fixed facilities or attend AS by mobile teams and are detected.

An improvement in detection rates in fixed care facilities was fitted to the data, as reported in Rock et al. [21]. Both true and false positives are simulated, depending on the specificity and sensitivity of the algorithm in operation for the given year. Serological positives (S+) are defined as those cases that were positive by the card agglutination test for trypansomiasis (CATT) 1:16 dilution, but for whom no trypanosomes were found in the blood. Parasitologically confirmed (P+) cases are those for whom trypanosomes can be microscopically visualised in blood. Patients who also have trypanosomes in their cerebrospinal fluid or an elevated white blood cell (WBC) count of more than 5 WBC/*µ*l are classed as stage 2 cases, otherwise they are classed as stage 1 cases. The remaining unreported prevalence was assumed to result in undetected deaths (see Section S1.2.1 in Supplementary Information S1 Text).

The population is projected to be stable. The population screened by mobile units is fixed based on the data before 2020, and according to projected levels for different strategies starting in 2023. The population screened by passive screening is projected to be the mean coverage per clinic available 2014–2020. The probability of a case being found is assumed constant, as very serious cases are likely to be tested, but suspected cases with milder symptoms will likely be categorized as other diseases. The key outputs of the transmission and detection model include deaths among undetected cases, cases that were S+ or P+ confirmed, detected cases in stages 1 and 2, and disability-adjusted life-years (DALYs) before and after presenting to care for all interventions.

Tsetse control via Tiny Targets was explicitly simulated in the model to match strategies with VC in our retrospective analysis and in all strategies from 2014 in our prospective analysis. This is done by including an additional mortality term in the tsetse model equations linked to the probability of a tsetse finding and receiving a lethal dose of insecticide during its host seeking phase of its gonotropic cycle. We assume that this probability is greatest directly after new Tiny Target deployment and gradually decreases due to a range of factors (*e*.*g*. vegetation growth, loss due to rain) over the year. The value was fitted based on the 2000–2019 case data and yielded a 99% reduction in tsetse after four months, in line with the reduction measured by tsetse traps density in the field [20, 21]. More details on modelling tsetse control in this ‘Warwick gHAT model’ framework are given elsewhere [21, 32]. Following cessation of VC (dictated by strategy choice) the tsetse population is allowed to recover in the model through removal of the additional tsetse mortality term in the equations.

#### Treatment model

P+ cases are treated as stage 1 or 2 cases according to the presence of trypanosomes or elevated WBC count in cerebrospinal fluid (stage 2 for positive findings). Patients who were positive according to CATT with dilution 1:16 but who were not confirmed by parasitology have also been treated; these serological cases have been classified as stage 1 or 2 depending on symptoms [21].

The outcome of treatment is modelled according a branching process formalized by a probability tree. The probability of treatment failure and diagnosis determines convalescence and death outcomes. Before 2020, there were only two treatments: pentamidine for stage 1 cases and nifurtimox-eflornithine combination therapy (NECT) for stage 2 (Supplementary Information S3 Text, Fig C). Starting in 2020, fexinidazole was available for inpatient and outpatient treatment of stages 1 and 2 according to patient characteristics, as delineated by WHO recommendations (Supplementary Information S3 Text, Fig C). Further details about the model are found in Supplementary Information S1 Text, Section S1.2 and in Rock *et al* [21].

### Outcome metrics

The gHAT transmission model simulates an open cohort and produces annual outputs of cases in stage 1 and stage 2 (partitioned by whether they were identified in AS or PS), person-years spent with stage 1 and stage 2 disease, undiagnosed (and therefore unreported) deaths, the number of new infections created, the number of people actively screened and whether or not vector control occurred. These were computed for each of the different analyses and strategies. Case outputs were used in turn in our probability tree model to determine which cases were given the different treatments (depending on stage and whether fexinidazole was yet available), and possibly second-line treatment based on the failure probability of first-line drugs. Additional deaths from failed treatments were also computed. Finally, to compute the DALY estimates under each strategy for each year, the total deaths (unreported and reported) were combined with the weighted person-years spend infected [33, 34]. Years lost of life posterior to the analytic horizon are counted in present-day terms by applying a 3% discounting rate.

The year of elimination of transmission (EoT) is defined in our deterministic model as the first year in which less than one case is found, in line with the proxy threshold used to determine EoT in other gHAT modelling [35]. Whilst it is imperfect, this level of new infections has been found to be an adequate approximation of the threshold to simulate local extinction as characterized in comparisons with stochastic models [36]. The probability of EoT by 2030 is the proportion of the 5000 iterations where the number of cases – detected or undetected – reaches less than one case before or in the year 2030.

### Costs

The costs were calculated from the perspective of the healthcare payers collectively, which include the government and all donors. A complete accounting of past costs for all interventions is nearly impossible because of the heterogeneous funding landscape for gHAT activities in Chad – a phenomenon that is present in many other gHAT-affected countries [37]. However, a detailed costing approach has been applied to vector control, including by contributor [19], but screening and treatment activities are not specifically designated for gHAT in national health accounts in Chad. Therefore, although we have health-related data from the field, and limited resource-related data, so we model the costs in order to come up with a holistic view of the costs involved in the endeavor from 2014. Costs are drawn from a variety of sources, from Chad whenever possible, and updated to 2020 values by taking into account the changes in the inflation rates as well as the exchange rates from local to US dollars, which we detail in the Supplementary Information S2 Text.

Moreover, we model probabilistic uncertainty around how much costs would have been in different situations, based on combining our unit cost estimates with dynamic outputs from the transmission model (number of people screened, number of vector control deployments) and probability tree model (numbers of different treatments given) (see Supplementary Information S1 Text, Section S1.2). This approach is similar to that employed in another gHAT cost-effectiveness study for the DRC [37], although each analysis is adapted to the country context. Cost components were broken down according to mean components and means of totals (since the sum of the means is not necessarily the mean of the sums). Final uncertainty was presented as the mean and the 95% predictive intervals (2.5th and 97.5th percentiles). The biggest drivers of uncertainty were determined by calculating the Expected Value of Perfect Partial Information - or the value to the program of reducing uncertainty to zero of any particular parameter. Due to the complex nature of the current model, the EVPPI was calculated by statistical approaches devised in previous literature [38].

### Cost-effectiveness analysis

Our cost-effectiveness analyses (one retrospective and one prospective) were performed from the perspective of the donors and ministry of health in Chad combined, only accounting for direct costs (not indirect costs such as out-of-pocket expenses or days of lost work accrued by patients and their families). In each analysis our default time horizon was until 2040 (2014–2040 for retrospective, and 2021–2040 for prospective) although shorter and longer time horizons were explored as sensitivity analyses. We selected 2040 as the default end point to better capture any benefits of EoT and/or cessation of activities - particularly during the 10 years following the goal year of 2030. This is one reason why we still used a default of 2040 for our time horizon in the retrospective analysis so that the assessment of cost-effective strategies factored in medium term benefits in the case that EoT occurs.

For each analysis we selected one strategy to be our comparator strategy, representing a “status quo”. For the retrospective study the comparator *(Pre-2014*) is a counterfactual strategy representing the continuation of AS and PS activities without any additional improvements to PS nor introduction of VC. For the prospective the comparator is *Mean AS & VC (a)* which assumes that, from 2023, AS and VC continues at the average level until there are three years of no parasitologically (P+) or serologically (S+) confirmed cases, alongside continued annual deployment of VC and sustained PS at the 2019 level. Although horizons run until 2040, the activities might cease before that according the the cessation criteria set forth in Table 1.

Our main metric is the incremental cost-effectiveness ratio (ICER), defined as the additional costs incurred divided by the additional DALYs averted between one strategy and the next best strategy, taking out weakly or strongly dominated strategies [39]. In these calculations, both DALYs and costs were discounted at 3% per year. We used ICERs to determine our recommendation change points depending on the policy maker’s willingness to pay (WTP) to avert disease. WTP is challenging to enumerate precisely as there are a range of financial and political factors dictating how much resource is allocated for specific disease programs. However, literature estimates have previously indicated that, in Chad, other disease programs have had WTPs of $30–518 per DALY averted [40]. In 2020, the per capita gross domestic product (GDP) in Chad was $659 [41], so based on previous WHO guidelines a strategy could have been considered to be “very cost-effective” at $659 per DALY averted or “cost-effective” at $1977 [34].

Four mechanisms drive uncertainty in our modelling framework: the uncertainty in the parameterization of the dynamic model based on fitting to historical data, the stochastic nature of disease reporting and deaths, uncertainty surrounding treatment outcomes, which were parameterized based on literature based-estimates (see in supplementary information Text S4 Text, and uncertainty in cost estimates [21]. To account for all these different types of uncertainty, DALYs and costs are simulated by sampling 5,000 iterations for each of the alternative strategies in each analysis. Our outcomes are presented as means and 95% prediction intervals (bounded by the 2.5th and 97.5th percentiles of the simulated samples).

We use the net-benefit framework, which uses the net monetary benefits (NMB) as its key metric in order to account for parameter uncertainty in the cost-effectiveness analysis:

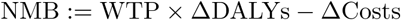

which is a simple reworking of the ICER equation so that the NMB are positive when WTP is more than the ICER 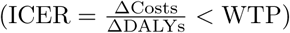. Because we have run 5,000 Monte Carlo simulations, then we have 5,000 samples of NMB. Within this framework, the probability that a strategy is optimal is the proportion of samples for which the strategy yields the highest NMB for a given WTP, and the overall optimal strategy is the strategy that yields the maximum mean NMB.

We depict the uncertainty across a range of WTP values, as recommended by conventions, using a cost-effectiveness acceptability curve (CEAC) [42–44]. The CEAC shows the probability that each strategy is optimal in a line graph against increasing values of WTP, while highlighting the optimal strategy at each WTP according to the mean NMB.

#### Sensitivity analysis

One scenario analysis was run to answer the question whether our assessment of cost-effectiveness would look qualitatively different in a similar context to Mandoul but in the present, in which fexinidazole is available.

Additional two-way sensitivity analyses were run. For the retrospective this included two shorter time horizons (10-year time horizon of 2014–2023, as well as a time horizon than ends in 2030) and one longer (2014–2050). For the prospective analysis there was one shorter (2021–2030) and one longer (2021–2050) time horizon. Under each time horizon we produce results with and without discounted benefits and costs. These are available in the project website: https://hatmepp.warwick.ac.uk/MandoulCEA/v1 and well as in the supplementary information.

### Computational considerations

The transmission model was fitted and projections simulated using Matlab 2018b. Analyses using the probability tree, cost model and cost-effectiveness were performed using R version 4.1.1. Hardware needs are detailed in the accompanying Open Science Framework repository (https://osf.io/bjxwn/) which contains the code used to perform the analyses. For interested readers and policy-makers, we created a project website to showcase results and sensitivity analysis: https://hatmepp.warwick.ac.uk/MandoulCEA/v1.

## Results

The model has indicated that there was a quickly decreasing incidence of new infections in Mandoul from 2014 which preceded the accelerated decline in the number of cases reported. Moreover, the model also shows that while infections as well as cases reported would have declined even if *Pre-2014* strategy had remained in place, the decline was accelerated by the additional activities of VC deployment and improved PS. Overall, after an initial increase in costs to set up improved PS and to start deployment of VC, costs showed a modest decrease from 2014–2020, but a more accelerated decrease will occur in the next five years, as interruptions in transmission will allow the safe scale-back of vertical activities (Table 3).

**Table 3.** Retrospective analysis. Summary of effects, costs, elimination of transmission (EoT) by 2030, and cost-effectiveness with and without uncertainty. Means are given along with 95% prediction intervals (PIs). YLL: years of life lost (to fatal disease), YLD: years of life lost to disability, DALYs: disability-adjusted life-years, PS: passive screening, AS: active screening, VC: vector control, ICER: incremental cost-effectiveness ratio, WTP: willingness to pay (USD per DALY averted), EoT: elimination of transmission.

|  | Pre-2014 | Improved PS | Addition of VC | Improved PS & VC |
| --- | --- | --- | --- | --- |
| **Health effects** |  |  |  |  |
| Reported cases | 1432 (694, 2538) | 1066 (543, 1896) | 185 (93, 315) | 216 (115, 341) |
| Deaths undetected | 1189 (520, 2365) | 533 (102, 1525) | 174 (95, 302) | 142 (62, 272) |
| Cases total | 2620 (1256, 4880) | 1598 (690, 3361) | 359 (215, 556) | 358 (208, 549) |
| Deaths detected<sup>a</sup> | 2 (0, 4) | 1 (0, 3) | 0 (0, 1) | 0 (0, 1) |
| YLD | 862 (456, 1527) | 430 (171, 909) | 111 (62, 186) | 96 (49, 173) |
| YLL | 53,572 (23,287, 105,795) | 24,029 (4,622, 69,545) | 7,862 (4,260, 13,582) | 6,409 (2,790, 12,387) |
| DALYs | 54,434 (23,771, 107,086) | 24,459 (4,902, 70,546) | 7,972 (4,361, 13,697) | 6,505 (2,881, 12,499) |
| **Costs, in thousands US\$** | | | | |
| AS costs | 1062 (774, 1436) | 973 (590, 1383) | 405 (294, 553) | 406 (294, 559) |
| PS costs | 66 (38, 106) | 1100 (714, 1548) | 66 (38, 106) | 546 (388, 751) |
| VC costs | 0 (0, 0) | 0 (0, 0) | 700 (308, 1227) | 703 (308, 1241) |
| Treatment costs | 540 (269, 944) | 441 (234, 754) | 105 (62, 161) | 122 (73, 181) |
| Costs total | 1668 (1244, 2199) | 2513 (1666, 3280) | 1277 (856, 1836) | 1777 (1305, 2394) |
| **EoT** |  |  |  |  |
| Year of EoT | After 2050 | 2043 (2029, After 2050) | 2015 (2015, 2015) | 2015 (2015, 2015) |
| Prob EoT 2030 | <0.01 | 0.06 | >0.99 | >0.99 |
| **Cost-effectiveness without uncertainty (discounted)<sup>b</sup>** |  |  |  |  |
| DALYs averted | 0 | 12,378 | 19,941 | 20,718 |
| Costs averted | 0 | 616,079 | -105,862 | 302,670 |
| ICER | Dominated | Dominated | Min Cost | 526 |
| **Cost-effectiveness with uncertainty, conditional on WTP<sup>c</sup>.** |  |  |  |  |
| WTP: \$0 | 0.32 | 0 | 0.68(p) | 0 |
| WTP: \$250 | 0 | 0.01 | 0.89(p) | 0.1 |
| WTP: \$500 | 0 | 0 | 0.53(p) | 0.47 |
| WTP: \$750 | 0 | 0 | 0.34 | 0.66(p) |
| WTP: \$1000 | 0 | 0 | 0.25 | 0.75(p) |

<sup>a</sup> Detected deaths are those that occur due to treatment failure or loss-to-follow-up.
<sup>b</sup> Cost-effectiveness results are given for discounted DALYs and costs as per convention
<sup>c</sup> (p) is the preferred strategy; the strategy with the highest mean net monetary benefits

**Table 4.**
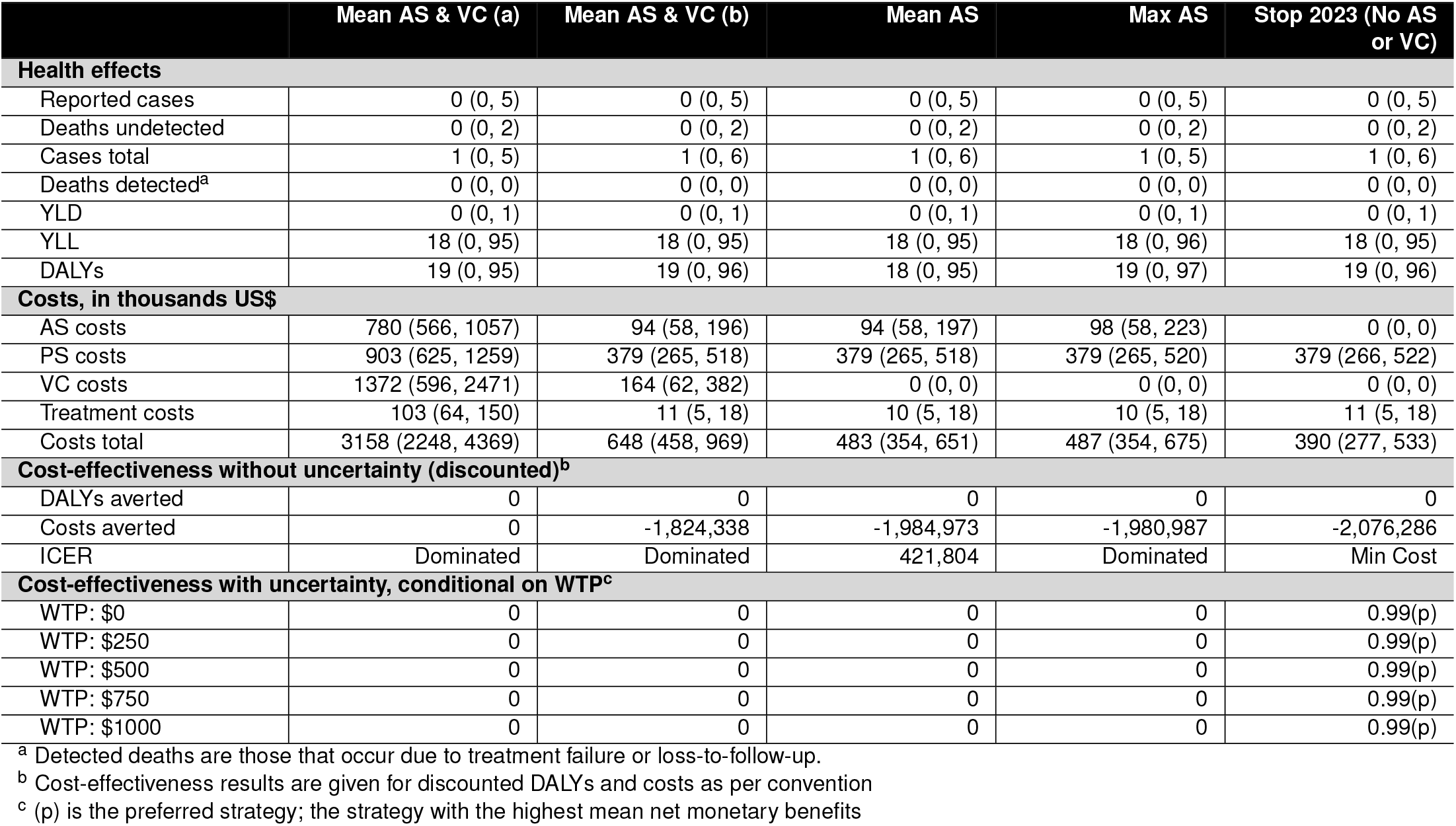
Prospective analysis. Summary of effects, costs, elimination of transmission (EoT) by 2030, and cost-effectiveness with and without uncertainty. Means are given along with 95% prediction intervals (PIs). YLL: years of life lost (to fatal disease), YLD: years of life lost to disability, DALYs: disability-adjusted life-years, PS: passive screening, AS: active screening, VC: vector control, ICER: incremental cost-effectiveness ratio, WTP: willingness to pay (USD per DALY averted)

|  | Mean AS & VC (a) | Mean AS & VC (b) | Mean AS | Max AS | Stop 2023 (No AS or VC) |
| --- | --- | --- | --- | --- | --- |
| **Health effects** |  |  |  |  |  |
| Reported cases | 0 (0, 5) | 0 (0, 5) | 0 (0, 5) | 0 (0, 5) | 0 (0, 5) |
| Deaths undetected | 0 (0, 2) | 0 (0, 2) | 0 (0, 2) | 0 (0, 2) | 0 (0, 2) |
| Cases total | 1 (0, 5) | 1 (0, 6) | 1 (0, 6) | 1 (0, 5) | 1 (0, 6) |
| Deaths detected<sup>a</sup> | 0 (0, 0) | 0 (0, 0) | 0 (0, 0) | 0 (0, 0) | 0 (0, 0) |
| YLD | 0 (0, 1) | 0 (0, 1) | 0 (0, 1) | 0 (0, 1) | 0 (0, 1) |
| YLL | 18 (0, 95) | 18 (0, 95) | 18 (0, 95) | 18 (0, 96) | 18 (0, 95) |
| DALYs | 19 (0, 95) | 19 (0, 96) | 18 (0, 95) | 19 (0, 97) | 19 (0, 96) |
| **Costs, in thousands US\$** | | | | | |
| AS costs | 780 (566, 1057) | 94 (58, 196) | 94 (58, 197) | 98 (58, 223) | 0 (0, 0) |
| PS costs | 903 (625, 1259) | 379 (265, 518) | 379 (265, 518) | 379 (265, 520) | 379 (266, 522) |
| VC costs | 1372 (596, 2471) | 164 (62, 382) | 0 (0, 0) | 0 (0, 0) | 0 (0, 0) |
| Treatment costs | 103 (64, 150) | 11 (5, 18) | 10 (5, 18) | 10 (5, 18) | 11 (5, 18) |
| Costs total | 3158 (2248, 4369) | 648 (458, 969) | 483 (354, 651) | 487 (354, 675) | 390 (277, 533) |
| **Cost-effectiveness without uncertainty (discounted)<sup>b</sup>** |  |  |  |  |  |
| DALYs averted | 0 | 0 | 0 | 0 | 0 |
| Costs averted | 0 | -1,824,338 | -1,984,973 | -1,980,987 | -2,076,286 |
| ICER | Dominated | Dominated | 421,804 | Dominated | Min Cost |
| **Cost-effectiveness with uncertainty, conditional on WTP<sup>c</sup>** |  |  |  |  |  |
| WTP: \$0 | 0 | 0 | 0 | 0 | 0.99(p) |
| WTP: \$250 | 0 | 0 | 0 | 0 | 0.99(p) |
| WTP: \$500 | 0 | 0 | 0 | 0 | 0.99(p) |
| WTP: \$750 | 0 | 0 | 0 | 0 | 0.99(p) |
| WTP: \$1000 | 0 | 0 | 0 | 0 | 0.99(p) |

<sup>a</sup> Detected deaths are those that occur due to treatment failure or loss-to-follow-up.
<sup>b</sup> Cost-effectiveness results are given for discounted DALYs and costs as per convention
<sup>c</sup> (p) is the preferred strategy; the strategy with the highest mean net monetary benefits

### Retrospective analysis

First, we ran counterfactual scenario analysis to assess whether previous increase in investment to gHAT activities in Mandoul represented a good use of resources.

#### Transmission and reporting

As outlined in our previously published transmission model analysis [21], strategies without VC would have had a very low probability of EoT by 2030: *<*1% if the strategies before 2014 had continued, or 6% if the network of RDT-based screening had been expanded (along with the motorcycle surveillance that we included). However, with VC, the probability increases to *>*99% for both strategies, and we believe that EoT has occurred since 2016, and all subsequently detected cases were infected in 2015 or before. See Table 3.

The disease burden is substantially lower than it would have been with alternative strategies. In 2014, the strategies present at the time were on track to see an additional 1,432 (95% PI: 694–2,538) detected cases by 2040 or 1,066 (95% PI: 543–1,896) detected if PS network had been improved (without use of VC). The improvement of PS would have cut not just a third of cases, demonstrating the impact of PS on transmission dynamics, but more than halved deaths thanks to expanded treatment: from 1189 (95% PI: 520–2,365) to 533 (95% PI:102–1,525). VC strategies would have additionally cut by four-fifths the number of cases detected compared to the *Improved PS* strategy; adding VC to the 2014 strategies would have meant detecting only 185 (95% PI:93–315) cases or adding both VC and improved PS would have resulted in 216 (95% PI: 115–341) detected cases. However, deaths, which come primarily from undetected cases, are fewer in the strategy that implements both improved PS clinics and VC (*Improved PS & VC*, 142 (95% PI: 62–272)) than the strategy that detects fewer cases without improved PS (*Addition of VC*. 174 (95% PI: 95–302)). Because DALYs primarily arise from fatal cases (see Table 3) the pattern on the impact of DALYs follows the pattern of deaths: continuing the *Pre-2014* strategy, there would have been 54,468 (95% PI: 23,807–106,639) DALYs lost in Mandoul alone, but the addition of improved PS would have halved those losses to 24,467 (95% PI: 4,900–69,000). The trend of DALYs across the years is illustrated in Figure A.

#### Costs

Over the 27-year period of 2014–2040, the total costs would have been highest had the *Improved PS* strategy been implemented (without VC) ($2.51M vs. $1.67M for the comparator), but the addition of VC without improved PS would have lowered the total costs ($1.28M), and the addition of both *Improved PS and VC* (actual strategy) is estimated to represent a very small increment in costs over the whole period ($1.78M). In 2015, it is estimated that costs increased about 60% in order to institute the actual strategy conducted (*Improved PS & VC*) compared to the continuation strategy (*Pre-2014*).

The investment in VC and improved PS was partially offset by averted treatment costs over a 27-year horizon, as well as the substantially reduced long-running costs of AS activities (Fig 3 and Supplementary Information S3 Text, Fig B). The costs of the strategy that was instituted (*Improved PS & VC*) were indeed higher for the period of 2014–2020, but the remaining costs in the strategy are the second lowest after the *Addition of VC* strategy (Fig 3 and Supplementary Information S3 Text, Fig B). Specific estimates of the costs from 2014–2020 compared to the whole period can be found in Supplementary Information S3 Text, Table A.

**Fig 3.**
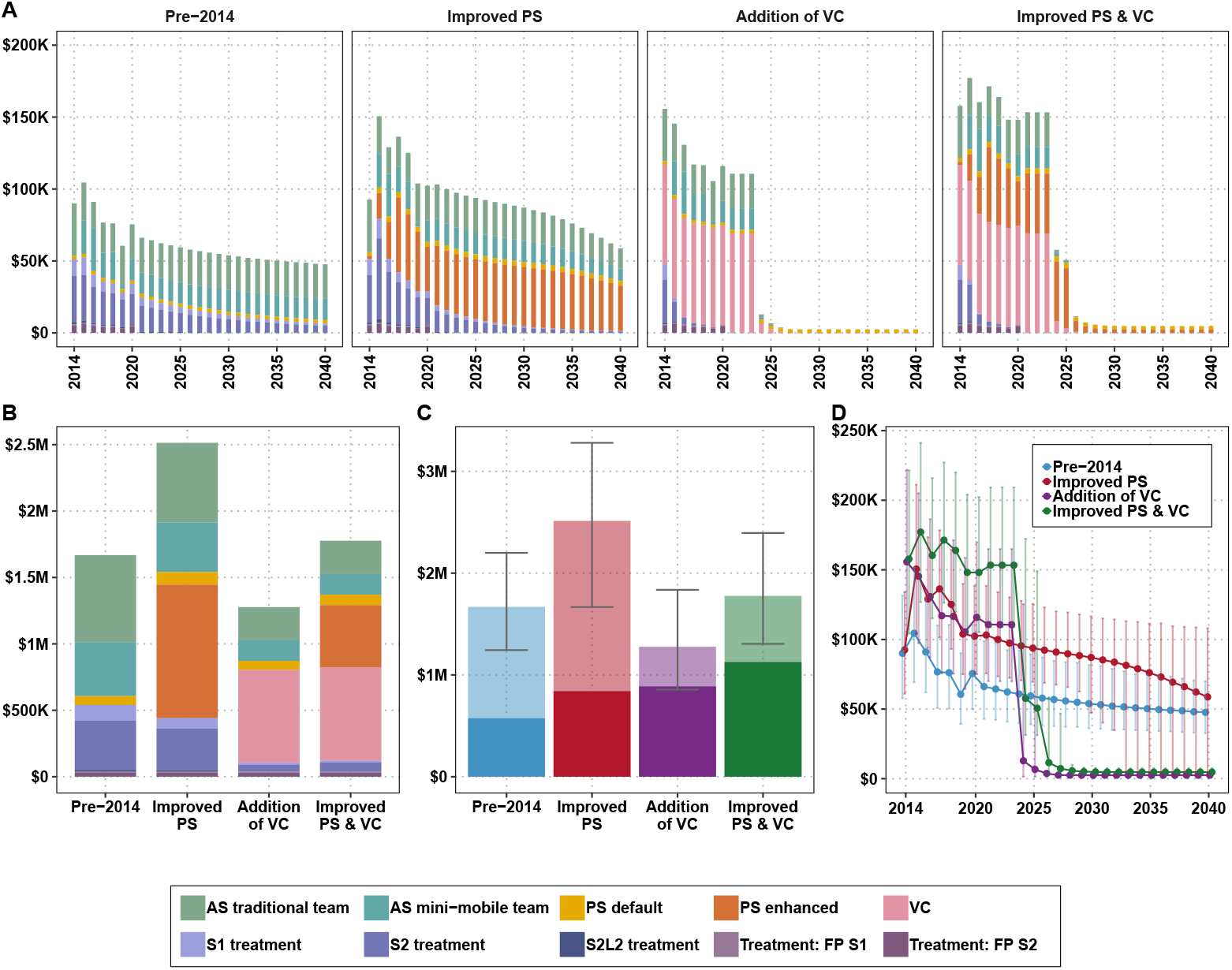
Costs by components of a strategy for the retrospective analysis: A) by year, B) for the period of 2014–2040, C) the costs for the period of 2014–2040 with uncertainty (transparent colors indicate the funds that have not been spent yet) and D) the total spent per strategy per year with 95% prediction intervals. See Supplementary Information S3 Text Table A and Fig B for total estimates and uncertainty of the costs spent between 2014–2020 compared to the whole time horizon (2014–2040). AS: active screening, PS: passive screening, VC: vector control, FP: false positive, S1: stage 1, S2: stage 2, S2L2: stage 2 rescue treatment.

**Fig 4.**
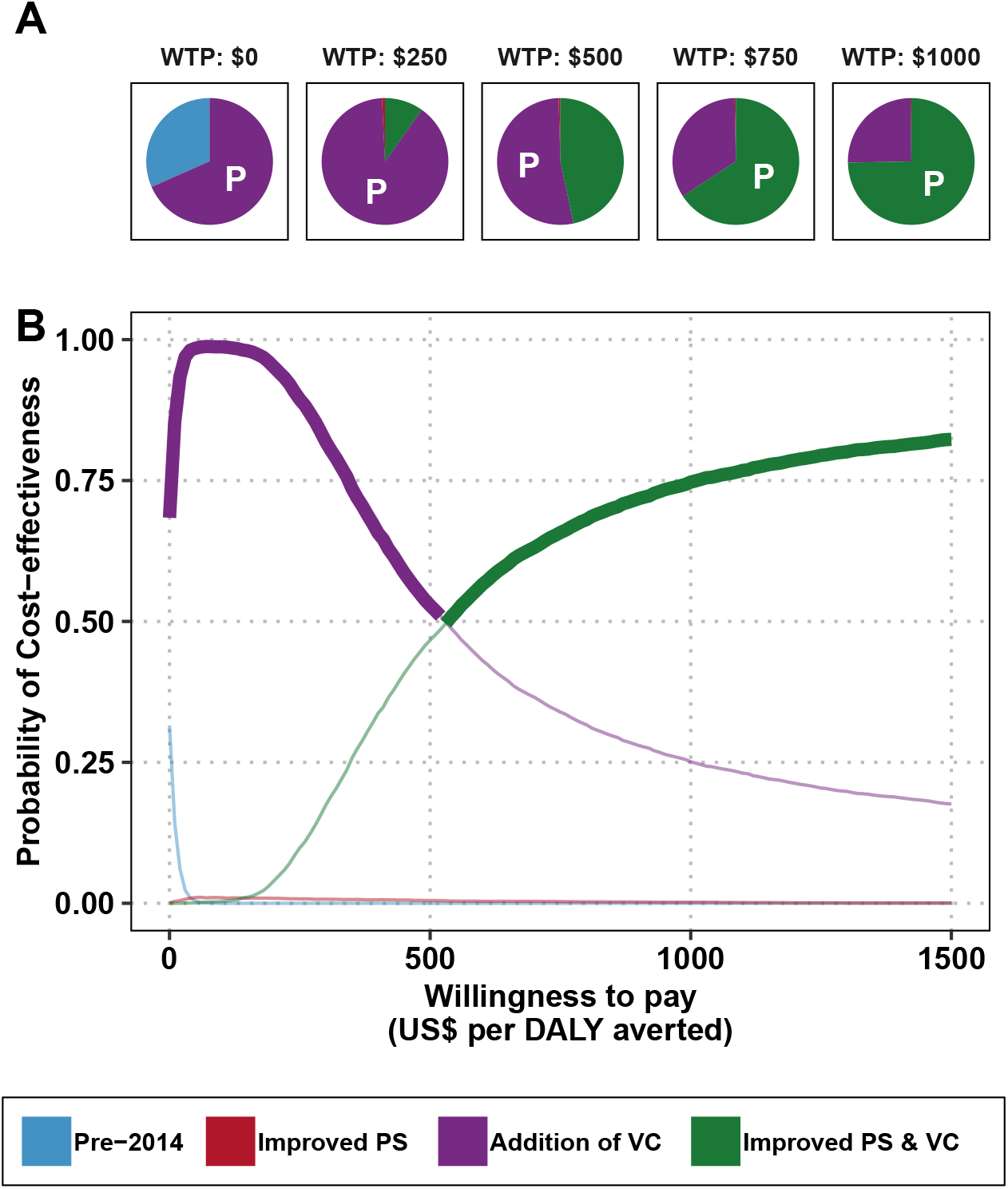
Uncertainty in cost-effectiveness for the retrospective analysis. A) Pie graphs depicting the probability that each strategy is optimal at the given willingness-to-pay (WTP) threshold. Strategies with the highest mean net monetary benefit are marked with a “P” for “preferred” strategy. B) Cost-effectiveness acceptability curves (CEACs) with the cost-effectiveness acceptability frontier (CEAFs) marked in bold. PS: passive screening, AS: active screening, VC: vector control, DALY: disability-adjusted life-years

**Fig 5.**
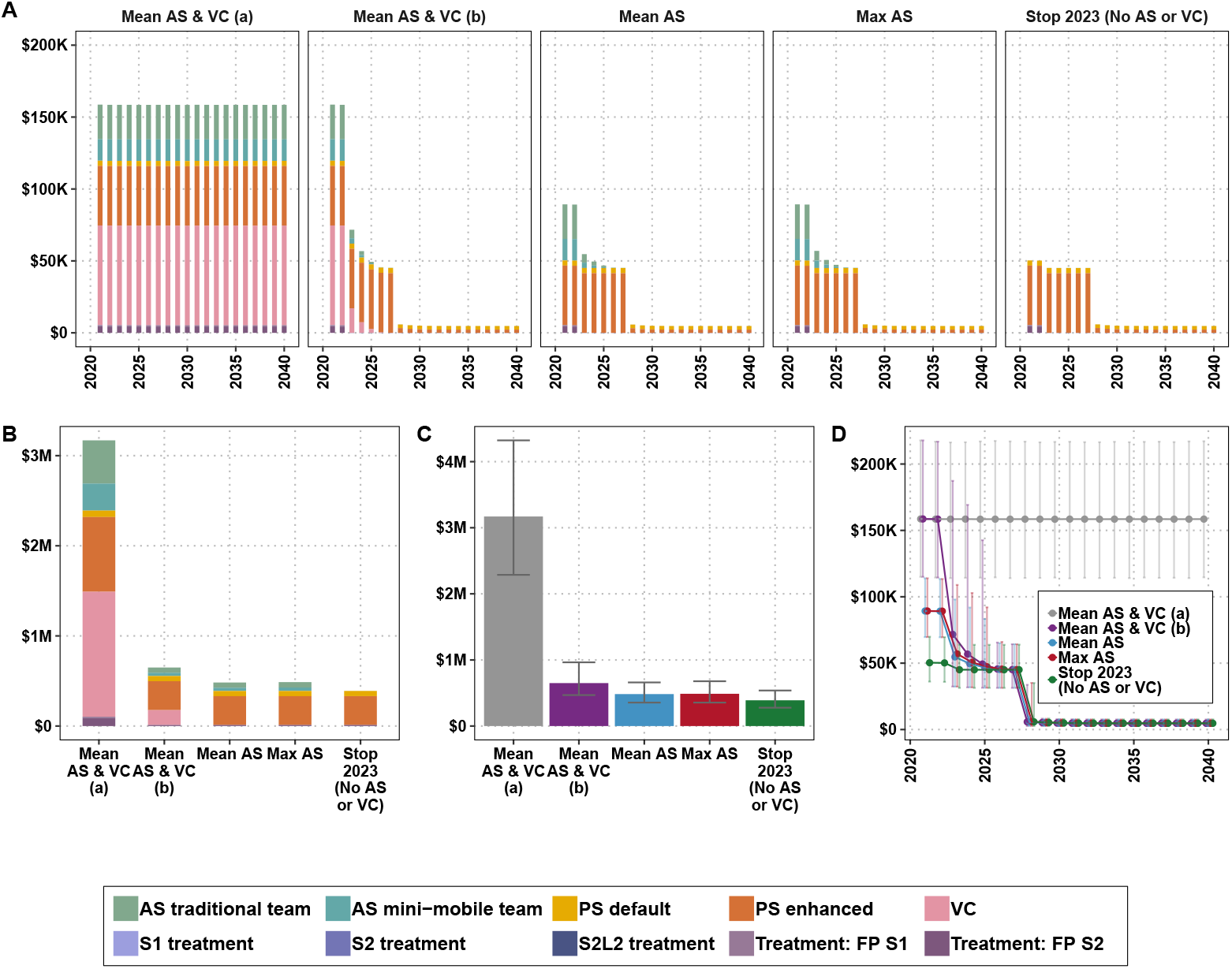
Costs by components of a strategy for the prospective analysis. A) by year, B) for the period of 2021–2040, C) the costs for the period of 2021–2040 with uncertainty, and D) the total spent per strategy per year with 95% prediction intervals. The specificity of the screening algorithm as part of the strategy *Mean AS & VC (a)* is 99.93% and for the strategies *Mean AS & VC (b), Mean AS* and *Max AS* it is 100%. *Stop 2023 (No AS or VC)* signifies that AS and VC stop immediately (it does not occur in 2023 onward). Mean AS is the coverage of people screened for 2000–2019. Max AS is the maximum coverage of people screened for 2000—2019. AS: active screening, PS: passive screening, VC: vector control, FP: false positive, S1: stage 1, S2: stage 2, S2L2: stage 2 rescue treatment.

### Cost-effectiveness

After taking a 3% discounting rate into account, the *Improved PS, Addition of VC*, and *Improved PS & VC* strategies would have averted 12,378 DALYs, 19,941 DALYs, and 20,718 DALYs, respectively. The strategy of *Improved PS* (and no VC) is *dominated*; in other words, while it costs more than any of the strategies with VC, it averts fewer DALYs. The investment of VC in addition to the continuation of the pre-2014 activities (*Addition of VC*) would have yielded cost savings by 2040, despite representing additional costs for the first few years. In fact, all strategies with VC would begin to yield cost savings in 2024, but will recover the investment completely by 2040.

Adding improved PS to the VC strategy (*Improved PS & VC*) costs an additional $408,532 (after taking into account 3% yearly discounting), for an additional 777 DALYs averted compared to *Addition of VC*; therefore, the ICER for the *Improved PS & VC* strategy is $526 per DALY averted. In short, while the investment in VC was cost-saving, the investment in improved PS was cost-effective at a willingness-to-pay of $526 per DALY averted (Table 3). At $500 per DALY averted, there is 47% probability that the implemented strategy was optimal, while at a WTP of $1000 per DALY averted, the probability that the strategy was optimal is 75%.

### Prospective analysis

Going forward, imperfect test specificity in AS will incur direct costs in over-treatment, but those costs will be overshadowed by the inability to confidently cease vertical activities (i.e. VC and AS) (Figure 2B). For all strategies with perfect diagnostic specificity, different combinations of interventions are predicted to make no difference in the number of cases detected (if there are any cases to be detected). The trend of DALYs across the years is illustrated in Supplementary Information S3 Text, Fig C.

According to this analysis, any strategies with interventions other than basic continuation of PS are not cost-effective. What we conceived as this bare-minimum strategy should cost around $390,000 if vertical interventions are stopped in 2023, or at most $648,000 for the period of 2021–2040 if VC and AS continue until no more parasitologically confirmed cases are detected.

### Scenario analyses

Different choices in times horizons or discounting of costs or disease burden would not have made a qualitative difference in our decision analysis, short of slight changes in the WTP values at which the *Improved PS & VC* strategy is cost-effective (see Figure E). Interested readers can explore the results of all scenario analysis on the project website https://hatmepp.warwick.ac.uk/MandoulCEA/v1/.

There was a difference in the results when we simulated treatment with fexinidazole for those patients who are eligible, but this different would not change the optimal strategy substantially (see Supplementary Information S3 Text, Table B). Fexinidazole treatment would have lowered the costs of treatment by 41–45% but the impact on total costs would have been a more modest reduction of 3–14%, since treatment costs constitute a small part of costs. With fexinidazole treatment for eligible cases, the strategy *Addition of VC* would no longer present the minimum cost but it would have an ICER of $2 per DALY averted. The strategy *Improved PS & VC* would have an ICER of $519 per DALY averted instead of $526 per DALY averted.

### Uncertainty analysis

When we performed an uncertainty analysis of the factors that contribute the most to decision uncertainty (see Supplementary Information S3 Text, Fig F), we found that the undetected deaths are the most influential parameter, followed by the years lost to disability before stage 2 cases are detected and the number of false positives considered stage 2 cases (S+ cases that would be P-). Other factors are the cost of microscopy, false positives considered stage 1 cases (S+ cases that would be P-), the cost of treatment for an outpatient visit, the capital cost of facilities that would be able to do screening only or screening and confirmation, and the age at the time of death of the cases. Because there is no uncertainty as to the economically optimal strategy prospectively, we did not find it beneficial to perform an uncertainty analysis for the prospective strategies.

## Discussion

Here, we present a case-study of cost-effectiveness for intensified strategies to reach EoT of gHAT and consider whether less ambitious new strategies or pre-2014 strategies would have been cost-effective. We evaluated the cost-effectiveness of the control program that was implemented from 2014 against less resource-heavy strategies to provide insights of the cost implications of such intensified elimination campaigns.

Transmission modelling and economic evaluations suggest that it may be cost-effective to halt active screening and vector control as long as passive screening remains robust. It is critical, however, to factor in data requirements for EoT to be verified and to protect against possible importation of infection from neighbouring endemic foci. It should be noted that in this analysis we do not explicitly model or cost activities aimed at verification of EoT, however active and passive detection efforts could form both control and measurement activities. Additional costs not included here likely include person-time to compile an elimination dossier, although at present it is unclear whether and how much additional screening or quality assurance would also be needed to meet the evidence.

This analysis features several novel properties over the only other past gHAT cost-effectiveness analysis. A study for Chad by Kohagne et al. [22] examined the most effective way to screen individuals in active screening, but it didn’t examine possible missed cases, nor modifications in other parts of the diagnostic or preventive cascade, like the addition of VC. Second, instead of using theoretical incidence levels [45], we employed a sophisticated, previously published model with parameters fitted to historical incidence data in Chad [20, 21]. This is the third time that this model – fit to local data – has been used in an economic analysis in a specific setting [37, 46], and it is the first time it has been used outside of the DRC – the country with the largest burden. One of the particular strengths of our analysis is that we used as much data as is available on the local transmission trends, economic costs, and resource use of gHAT interventions in Chad.

It should be noted that the addition of VC, in having such a quick and precipitous impact on transmission, also reduces the amount of uncertainty on burden and costs. For instance, the number of DALYs incurred by the *Improve PS* strategy and by the emph*{*Addition of VC strategy have very similar lower bounds 4902 vs 4361, respectively, but the upper bounds were 70,546 vs 13,367, respectively. The uncertainty in costs take longer to be reduced, but it is expected that the costs from 2021 on will be far less uncertain than those of less ambitious counterfactual strategies (see Figure 3).

We believe that the resources devoted to prevention and treatment of gHAT in Mandoul could now be diverted to address the existing burden in Moissala and Maro. Therefore, our study of the cost implications of the activities in Mandoul can serve as an illustrative guide for similar scale-ups of activities where appropriate in the effort to eliminate transmission in other persistently low-incidence locations of Sub-Saharan Africa. The most important insight from our prospective analysis is the implicit costs of false positives (S+ but P-cases); if false positives compel the program to continue vertical interventions, then the costs of delayed cessation are much higher than just the explicit cost of unnecessary treatment for people who might not truly be positive for the parasite. Now that fexinidazole is available, a place similar to Mandoul would still find the deployment of vector control economically efficient (see Supplementary Information S4 Text, Table B).

### Limitations

There are a few limitations to our analysis but we believe that they do not obscure the quality of the conclusions. First, cost-effectiveness by itself does not address operational feasibility; for instance, having sufficient stock of RDTs throughout the world is not conveyed here unless there is an impact on prices as a reflection of pressures on supply. However, supply chain issues are beyond the scope of any cost-effectiveness analysis. Moreover, the criteria to continue monitoring in case of disease resurgence or to verify the country as having achieving EoT might require resources that are beyond our calculations in this paper. The paper is meant to inform activities, but ultimately, national ownership of the strategies going forward are paramount for the success of the program.

Other foci in Chad contain important features different to those in Mandoul including different ecological and epidemiological settings so therefore we cannot conclude that the same strategies that were cost-effective in Mandoul would be cost-effective in Maro and Moissala. For instance, the tsetse population in those regions are not an ‘ecological island’, and Maro is contiguous with the Maitikoulou focus in the Central African Republic, thereby being at higher risk of disease re-introduction. Maro and Moissala likely still need sustained vertical interventions to reach local EoT, but that analysis is beyond the scope of the present study.

Second, there were a few simplifying assumptions on the structure of the model. We assumed no population growth. For the prospective analysis it should make no difference given the extreme low prevalence from 2019. For the retrospective analysis there would be small quantitative differences but not qualitative. This is a deterministic model, rather than a stochastic model which is more common to examine the time of EoT, but based on this groups other work we would expect to see very similar dynamics between deterministic and stochastic structures [36]. Furthermore, we do not consider spatial connectivity and re-importations in this paper, but we point the interested reader to the related publication by Rock et al. [21] which indicates that resurgence is unlikely even if VC stops and tsetse populations recover quickly. We do not scale PS back up after scale-down to the two clinics even if cases are detected. However, resurgence after the scale-down of PS happened in only 5 out of 5,000 iterations, so we believe it would have no impact on the conclusions that scale-back after five years would be appropriate. It should also be noted that the loss of expertise among staff members who rarely screen for gHAT cannot be sensibly accounted for in a cost-effectiveness analysis. Moreover, this analysis does not include self-curing asymptomatic infections (see [47, 48] and Model Y in [49]).

## Conclusion

Our analysis shows that the expansion of screening activities and the introduction of vector control against gHAT after 2014 was not only helpful in eliminating transmission of the parasite in Mandoul, but that it was a cost-effective use of resources. This indicates that similar expansions in small populations like those of Mandoul could be cost-effective, even in the presence of fexinidazole or an easier and cheaper drug for cases of either stage. Prospectively, we found that scale-back of activities in Mandoul would be economically sensible, as long as national public health and governance stakeholders are satisfied that transmission has been halted.

## Supporting information

S2 Text. Glossary of Technical Terms

S3 Text. Supplemental Results.

S4 Text. Parameter glossary.

S5 Text. PRIME-NTD Checklist.

S6 Text. CHEERS Checklist.

## Data Availability

All data produced are available online at https://osf.io/bjxwn/.

https://hatmepp.warwick.ac.uk/MandoulCEA/v1

## Supplementary Information

**S1 Text** An expansion of the materials and methods. **Figure A: Remaining gHAT foci in Chad**. All remaining gHAT foci in Chad are located in the Southern region of the country. The exact extent of the area of transmission for Mandoul is hard to precisely define. The Mandoul focus was determined by geolocating all the gHAT cases indicated as living in Mandoul in the WHO HAT Atlas. Reprinted from Rock et al. [21] under a CC-BY license. **Figure C: The gHAT transmission model and probability tree representing treatment outcomes**.A) The transmission model, depicting a Susceptible-Exposed-Infected-Recovered-Susceptible (SEIRS) model, which represents the progression of disease among low-risk humans (blue compartments), tsetse (purple compartment) and high-risk humans (red compartments), and the transmission of disease between the three groups. B) The probability tree representing treatment outcomes. Before 2020, a smaller tree was used constituting only the NECT and pentamidine branches for stage 1 and 2 disease, respectively, as fexinidazole was unavailable. **Figure B: Summary of active screening activities by mini-mobile teams in motorcycles. Table A: Summary of active screening activities by traditional teams in trucks**. Note: some of these cases might not be parasitologically confirmed. **Section 1.2** Additional method details covering the transmission model, strategy components, treatment model and health outcomes denominated as disability-adjusted life-years (DALYs). **S1.2.3 Text Additional method details covering the treatment model**.

**S2 Text** Glossary of technical terms.

**S3 Text** Additional results including figures and tables. **Figure C:** Disability adjusted life-years (DALYs) accrued through time for different counterfactual strategies in the prospective analysis. **Table E:** Treatment cost parameters. **Figure A:** Disability adjusted life-years (DALYs) accrued through time for different counterfactual strategies in the retrospective analysis. **Figure B: Close-up of costs spent vs to spend in retrospective analysis**. A) Costs spent by intervention for each strategy in 2014-2020. B) Costs spent vs those to be spent by activity. See table A for total estimates and uncertainty in each period. **Table A:** Costs spent vs total costs (in millions USD) for retrospective analysis. Means are given along with 95% prediction intervels (PIs). **Figure C:** Disability adjusted life-years (DALYs) accrued through time for different counterfactual strategies in the prospective analysis. **Figure E:** Cost-effectiveness acceptability curves (CEACs) for retrospective strategies, sensitivity analysis with alternative horizons with the cost-effectiveness acceptability frontier (CEAFs) marked in bold. **Table B: Sensitivity analysis if fexinidazole had been available since 2014**. Summary of effects, costs, elimination of transmission (EoT) by 2030, and cost-effectiveness with and without uncertainty. Highlighted lines are the results that changed after simulating treatment with fexinidazole (compare to Table 3). Means are given along with 95% prediction intervals (PIs). YLL: years of life lost (to fatal disease), YLD: years of life lost to disability, DALYs: disability-adjusted life-years, PS: passive screening, AS: active screening, VC: vector control, ICER: incremental cost-effectiveness ratio, WTP: willingness to pay (USD per DALY averted), EoT: elimination of transmission. **Figure F:** Expected value of perfect partial information (EVPPI) for retrospective analysis.

**S4 Text** Model parameter glossary.

**S5 Text** NTD-PRIME checklist.

**S6 Text** CHEERS checklist.

## Data Availability

Information about the data used for fitting is described in Rock *et al* [21]. The data used for clinical outcomes and costs were estimates from the literature. Screening data in this study were used to inform future potential screening coverage and were obtained through the WHO HAT Atlas, which cannot be shared publicly but it is under the stewardship of the WHO and available from the WHO (contact or visit https://www.who.int/teams/control-of-neglected-tropical-diseases/human-african-trypanosomiasis/atlas-of-hat) for researchers who meet the criteria for access to confidential data. Assumptions and estimates were parameterized according to conventions in the economic evaluation literature [50]. For assumptions around intervention, treatment effects and costs, see Supplementary Information S1 Text section S1.2.2 and S4 Text. Output data derived through the present study are available from OSF https://osf.io/bjxwn/“

## Code Availability

Full access to the code is available via Open Science Framework: https://osf.io/bjxwn/.

## Acknowledgements

The authors thank PNLTHA for original data collection, and the WHO for data access (in the framework of the WHO HAT Atlas [12]).

## Author Contributions

CRediT checklist:

- Conceptualisation: KSR, FT
- Methodology: MA, CH, AS
- Software: MA, CH, REC, SAS, KSR, PEB
- Validation: SAS, MA
- Formal analysis: CH, REC, MA
- Investigation: SB, AP, AS, IT
- Data curation: MP, SM, JD, PRB, XWS, MA, IT, PS, AS
- Writing - original draft: MA, KSR, EHC
- Writing - review and editing: MA, CH, SAS, REC, PRB, APS, PS, IT, AP, SB, PEB, SM, JD, XWS, EHC, MP, FT, KSR
- Visualisation: MA, PEB
- Supervision: KSR, FT
- Project administration: EHC, KSR
- Funding acquisition: KSR, FT, SB

