## Supplementary material for "Health economic evaluation of strategies to eliminate *gambiense* human African trypanosomiasis in the Mandoul disease focus of Chad": S1 Text. Supplemental Methods.

### S1. Supplementary Methods

#### Contents


S1.1 Locations . . . S1.2
S1.2 Models . . . S1.3
  S1.2.1 Transmission model . . . S1.3
  S1.2.2 Strategy components . . . S1.3
  S1.2.3 Treatment model . . . S1.7
  S1.2.4 Health outcomes denominated as disability-adjusted life-years (DALYs) . . . S1.7
S1.3 Cost functions . . . S1.9
  S1.3.1 Cost functions: active screening . . . S1.9
  S1.3.2 Cost functions: passive screening (screening at fixed health posts) . . . S1.14
  S1.3.3 Cost functions: vector control . . . S1.19
  S1.3.4 Cost functions: treatment . . . S1.19
References for SI Text 1 . . . S1.21


#### List of Figures


A Remaining gHAT foci in Chad . . . S1.2
B Decision trees . . . S1.4
C Transmission and treatment models . . . S1.5


#### List of Tables


A Summary of active screening activities by traditional teams . . . S1.6
B Summary of active screening activities by mini-mobile teams . . . S1.6
C Summary of passive screening in fixed health facilities . . . S1.7
D Parameters for treatment eligibility . . . S1.7
E Eligibility for treatment . . . S1.8
F Treatments and outcomes distributions for stage 1 and 2 patients . . . S1.8
G Active screening: cost function . . . S1.10
H Components of active screening costs. Full citations and explanations for the parameters will be given in Supplementary Information SI Text 4. . . . S1.10
I Cost breakdown for active screening activities . . . S1.10
J Passive screening: cost function . . . S1.15
K Components of passive screening costs. Full citations and explanations for the parameters will be given in Supplementary Information SI Text 4. . . . S1.15
L Cost breakdown for passive screening activities . . . S1.16
M Treatment: cost function . . . S1.19
N Parameters for treatment costs. Full citations and explanations for the parameters will be given in Supplementary Information SI Text 4. . . . S1.20
O Cost per person for gHAT treatment . . . S1.20

#### S1.1 Locations

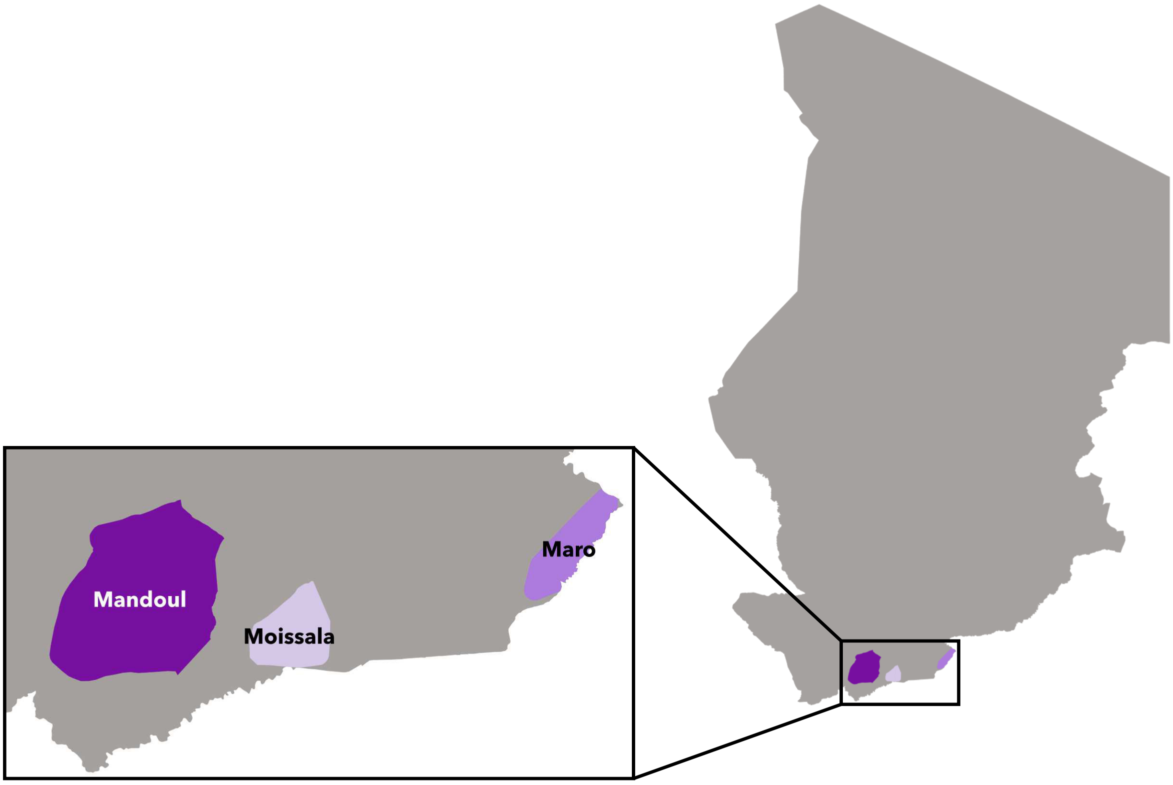


Figure A: Remaining gHAT foci in Chad. All remaining gHAT foci in Chad are located in the Southern region of the country. The exact extent of the area of transmission for Mandoul is hard to precisely define. The Mandoul focus was determined by geolocating all the gHAT cases indicated as living in Mandoul in the WHO HAT Atlas. Reprinted from Rock et al. [1] under a CC-BY license.

#### S1.2 Models

##### S1.2.1 Transmission model

We took epidemiological outputs from a set of deterministic transmission models by Rock et al. [1], fit to epidemiological data from the gHAT focus centered around Mandoul, Chad. These models are a derivative of the 'Warwick' gHAT models developed by Rock et al. [2] for gHAT transmission in Democratic Republic of Congo, and adapted for the Chadian context by Mahamat et al. [3] before the latest update [1]. The models used here were been described in detail in Rock et al. [1] and include heterogeneity in people's exposure to tsetse bites, some systematic non-participation in active screening (AS), and the possibility of non-human animal transmission (see Figure C). Fitting utilised a Markov chain Monte Carlo (MCMC) approach and staged, active and passive case data (2000–2019) from the WHO HAT Atlas and PNLTHA-Chad. The model variants used in the present study were selected based on statistical support for each based on Mandoul's human case data and weighted accordingly – this created an "ensemble" model which captures some structural uncertainty in gHAT epidemiology in Mandoul.

The ensemble model was used to project the impact of four strategies for our retrospective analysis: the status quo practice before 2014 of average AS, passive screening (PS), and no vector control (VC), which will be our referent case, and three other strategies made up of combinations of these interventions (and described in the main text). Likewise, prospective strategies described in the main text were simulated using this transmission model. While our primary outcome in the present study is the disability-adjusted life-year (DALY), we will also consider each strategy's capacity to reach the WHO's 2030 goal for gHAT: to stop transmission by 2030. In the deterministic model we use a proxy threshold of $< 1$ new infection per year to approximate when EoT occurs.

The transmission model calculates a range of important outputs to be used in the health economic evaluation: this includes the number of detected cases (active and passive) each year, and undetected deaths. It does this by triangulating various pieces of information which are included in simulations: 1) The model accounts for variable screening and corresponding case reporting – years with less screening typically result in fewer case detections and more people dying undetected 2) the model takes advantage of the ratio of stage 1 versus stage 2 cases identified – more stage 1 cases indicate more ongoing new transmission and more stage 2 cases indicate cases infected longer before. We also note that the stage ratio is expected to be skewed to early detection with AS which enables even people with mild or no symptoms to be identified and treated, whereas passive detection typically finds more people in stage 2 disease. From these ratios we can use the model fitting procedure to estimate the number of people who stop having stage 2 infection due to unreported deaths rather than those reported in the case counts. The parameter determining the proportion of cases that will go on to be reported rather than die ($u$) is one of the model parameters which is estimated through fitting to the case data.

Sensitivity analysis will look at the economic case of these interventions if fexinidazole had been available from 2014, as well as if the strategies remain good value for money with longer and shorter time horizons and with no discount rate. Structural uncertainty in the epidemiological model was taken into account via the ensemble model, and parameter uncertainty was incorporated by both the transmission model parameter estimation using the MCMC approach and using the net benefits framework with uncertainty in costs. Finally, value-of-information analysis will be performed to assess the most important sources of uncertainty.

##### S1.2.2 Strategy components

###### Active screening with traditional vehicle teams and with motorcycle teams (mini teams)

The examination of individuals in their village by mobile teams who screen and confirm cases. Vehicle teams go in two vehicles. Confirmation of suspects is done immediately via parasitology of the blood and cerebro-spinal fluid (for those who had trypanosomes in the blood). mAECT (minibar) is very rare, and it has only been used since 2017. See Kohagne *et. al* [4] and Mallaye *et. al* [5] for further explanations of the algorithm for traditional active surveillance.

PNLTHA manages the vehicle teams and they operate on a federal basis, so teams can move across cantonal or health district borders. Multiple villages are screened on the same day. Motorcycle ("mini-mobile") teams are confined to one health district. More recently, motorcycle teams have begun to perform follow-up surveillance (either carrying the parasitology equipment or transporting the suspected case to a hospital with capacity for parasitology). See the informational brochure by FIND for further explanations of the motorcycle teams [6, 7].

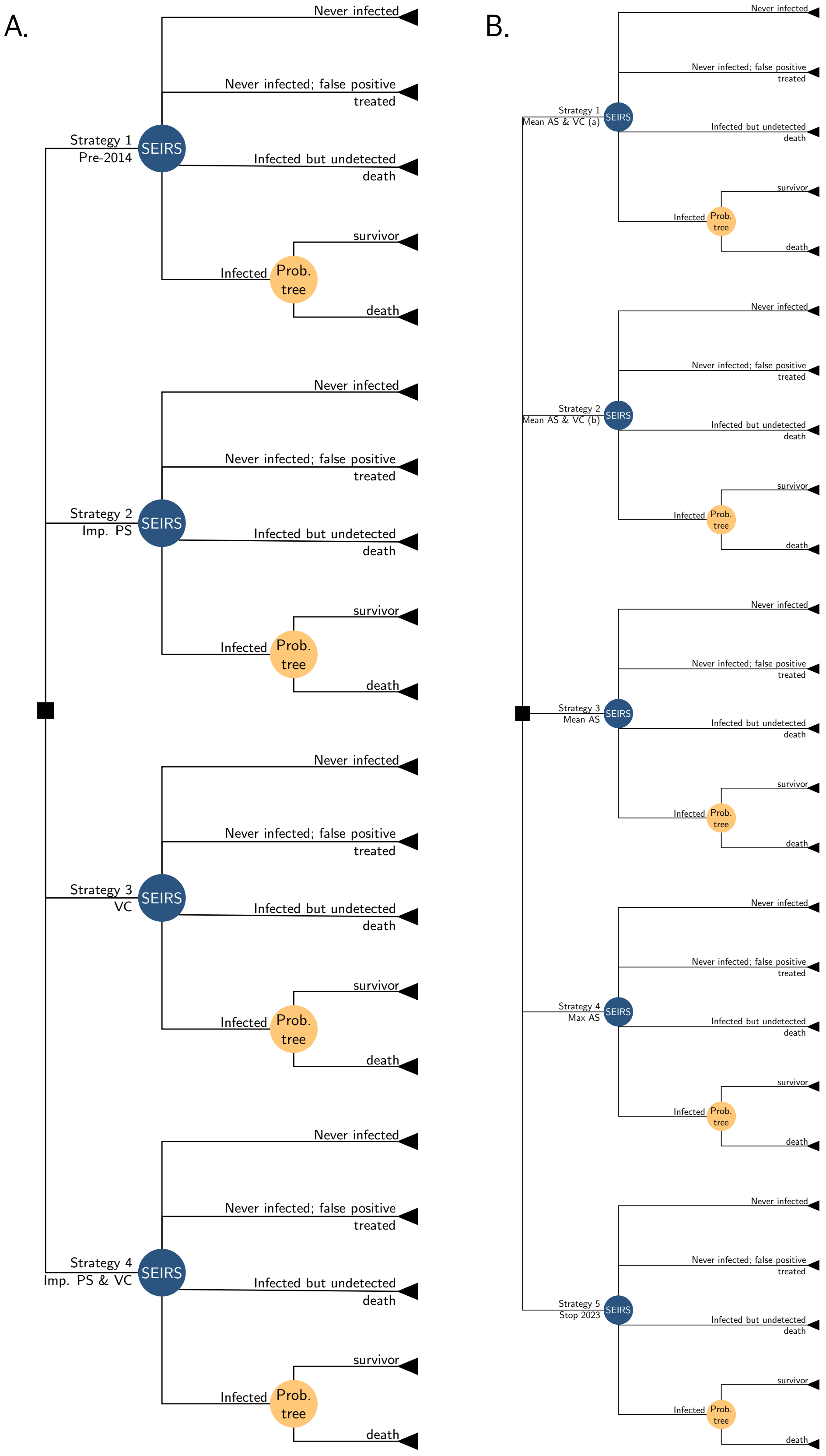


Figure B: A) Decision tree among four strategies for the retrospective analysis. B) A fifth branch is added for the prospective analysis, as there were 5 strategies to compare. For each strategy, the top two resulting branches of the model come from the transmission model only, and the bottom two resulting branches come from the treatment tree, which is fed outputs of detected cases from the transmission tree.

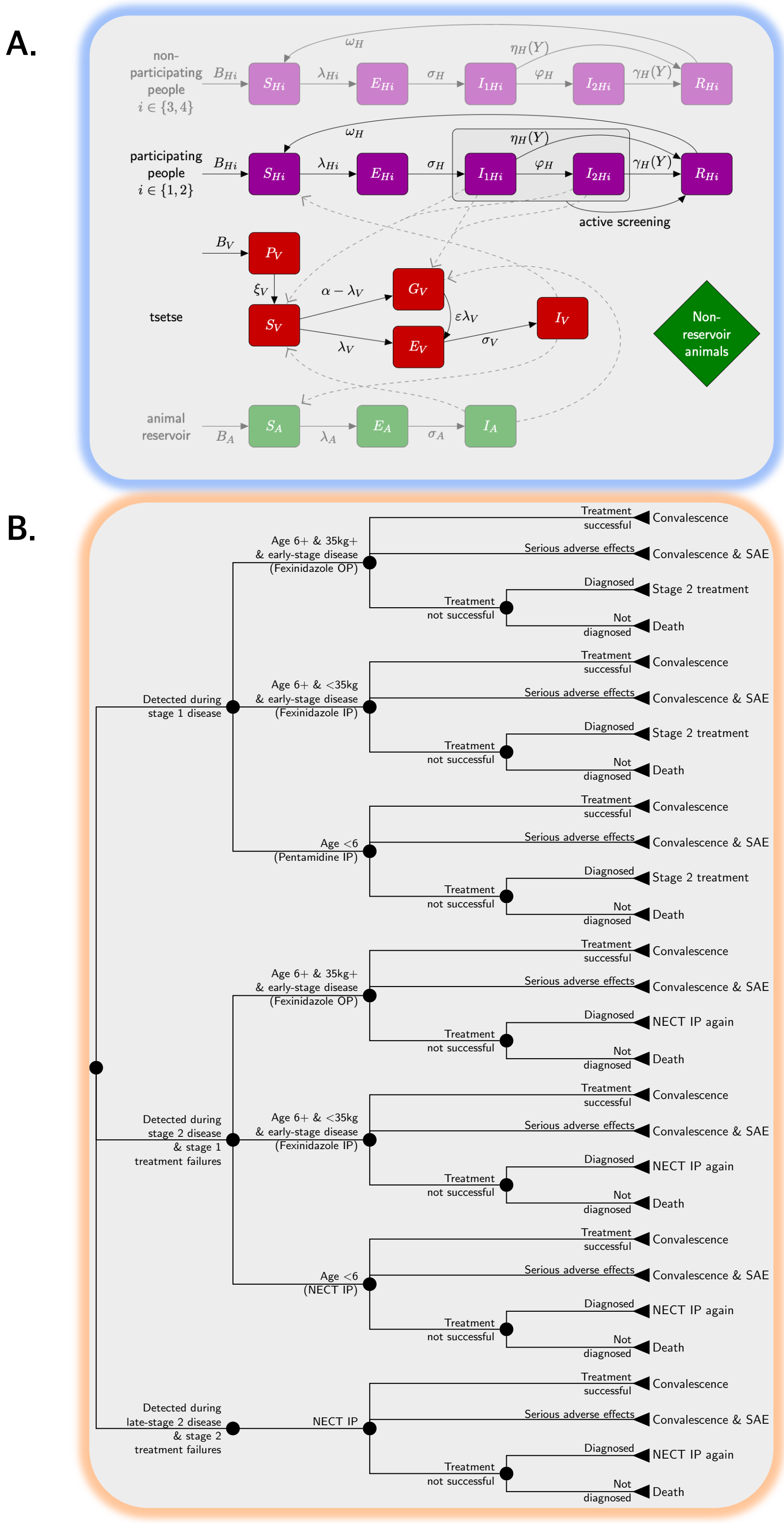


Figure C: A) The transmission model, depicting a Susceptible-Exposed-Infected-Recovered-Susceptible (SEIRS) model, which represents the progression of disease among low-risk humans (blue compartments), tsetse (purple compartment) and high-risk humans (red compartments), and the transmission of disease between the three groups. B) The probability tree representing treatment outcomes. Before 2020, a smaller tree was used constituting only the NECT and pentamidine branches for stage 1 and 2 disease, respectively, as fexinidazole was unavailable.

Traditionally, the stage of disease is determined by microscopy examination of the cerebro-spinal fluid, which is extracted via lumbar puncture for cases in which trypanosomes are present in the blood. Depending on whether trypanosomes are present in the cerebro-spinal fluid (stage 2) or not (stage 1) patients are referred to the appropriate health center or health district hospital for treatment. In the context of fexinidazole treatment, we do not expect that lumbar punctures will be performed by the active screening team, but rather that patients are referred to a health center that will determine eligibility for fexinidazole treatment. However, fexinidazole has only recently been approved for use and how eligibility takes place and lumbar punctures are administered is to be seen.

*Coverage.*

The WHO Atlas shows how many activities occurred and how many people were screened, but not how many people were targeted. From programmatic data we know that in 2017, there were 58 trips to target villages of between 178 to 2520 people. The coverage was an average of 40%, overall reaching as few as 8% or as many as 79% (IQR: 29%-49%). In the prospective analysis, "Mean AS" is equal to the average number of people screened for the period 2014–2019 (19,628) whereas under a "Max AS" strategy, the number of people screened is the maximum screened in any single year between 2000–2019 (22,146). In the prospective analysis, the actual screening numbers are applied in 2014–2019, then for 2020–2022 "Mean AS" is assumed for all strategies, and for 2023 and beyond "Mean AS", "Max AS", or "Stop AS" is applied depending on the strategy.

**Reactive screening**

Reactive screening (RS) is equivalent to AS, but it occurs after a case has been identified in the focus where AS had ceased after a three-year period of no cases.

| Year | Villages | Pop. Scr. | Cases | Scr per Village |
|---|---|---|---|---|
| 2014 | - | 22146 | - | - |
| 2015 | - | 17932 | 6 | - |
| 2016 | - | 11302 | 5 | - |
| 2017 | 58 | 9284 | 0 | 278 |
| 2018 | 43 | 10646 | 0 | 421 |
| 2019 | 62 | 11411 | 10 | 204 |
| 2020 | 96 | 18445 | 11 | 204 |

Table A: Summary of active screening activities by traditional teams in trucks. Note: some of these cases might not be parasitologically confirmed.

| Year | Villages | Pop. Scr. | Sero+ | LTFU | Cases | Scr per Village |
|---|---|---|---|---|---|---|
| 2014 | 0 | - | - | - | - | |
| 2015 | 114 | 9333 | 98 | ? | 6 | 82 |
| 2016 | 99 | 10705 | 176 | 10 | 5 | 108 |
| 2017 | 93 | 6860 | 165 | 95 | 0 | 74 |
| 2018 | 101 | 7437 | 213 | 75 | 0 | 74 |
| 2019 | 25 | 1229 | 53 | 12 | 0 | 49 |
| 2020 | 14 | 1182 | 22 | 1 | 0 | 85 |

Table B: Summary of active screening activities by mini-mobile teams in motorcycles.

**Passive screening**

gHAT screening that occurs in local health posts of patients who present themselves with specific gHAT symptoms. PS, or detection in fixed health facilities is assumed to take place with RDT diagnostic tests used for initial serological screening. For suspects that are RDT-positive, the health worker examines lymph node glands and performs microscopy exams, consisting of a blood sample taken and examined via microscopy, and since 2015, via LAMP. For the scale-back, the network of clinics capable of screening patients is scaled back rather than complete cessation; Bodo Hospital hospital, which performs both screening and confirmation capacities would be left.

*Coverage.* Coverage of PS was assumed to depend on the number of health centres that can perform a serological confirmation of gHAT. See Table C for the data on the coverage throughout 2014–2020.

| Year | Facilities - old | Facilities - new | Pop. Scr. Total | Sero+ | LTFU | Cases | Scr per Fac. |
|---|---|---|---|---|---|---|---|
| 2014 | 1 | | - | - | - | - | |
| 2015 | 1 | 9 | 892 | 31 | 5 | 22 | 97 |
| 2016 | 1 | 9 | 1382 | 30 | 0 | 8 | 138 |
| 2017 | 1 | 30 | 2289 | 29 | 0 | 6 | 74 |
| 2018 | 1 | 30 | 2277 | 21 | 7 | 2 | 73 |
| 2019 | 1 | 22 | 2651 | 7 | 3 | 1 | 115 |
| 2020 | 1 | 22 | 1354 | 11 | 6 | 0 | 58 |

Table C: Summary of passive screening in fixed health facilities

**Vector control**

consists of an annual deployment of Tiny Targets to control the population of tsetse. The impact of vector control in Mandoul is fit by the model; we estimate a 99% decrease in the population of tsetse in the first 4 months.

##### S1.2.3 Treatment model

Detected cases (either active or passive) are referred to the district hospital for treatment according to WHO guidelines. The specificity of the screening algorithm as part of the strategy *Mean AS & VC (a)* is 99.93% and for the strategies *Mean AS & VC (b)*, *Mean AS* and *Max AS* it is 100%. *Stop 2023 (No AS or VC)* signifies that AS and VC stop immediately in 2023.

For the retrospective analysis, we built the treatment algorithm according to the practice documented in the literature regarding the use of pentamidine (for S1 cases) and the use of NECT (for stage 2 cases) (see Fig C). Some documented cases were assumed to be false positives, since there was evidence of treatment of cases that were confirmed by a CATT 1:16 test. All S1 cases are assumed to have been treated on an outpatient basis. No use of older treatment was assumed (i.e. melarsoprol) in the period of 2014-2020.

For the prospective analysis (2021-2040), we built the treatment algorithm based on the WHO interim recommendations of 2019 [8] regarding the use of fexinidazole, and it is therefore a more elaborate model (see Fig C). For more details, see the supplementary information in Antillon *et. al*, Supplementary Note A, section 2.3 [9].

| Patient Characteristic | Parameterization | Summary |
|---|---|---|
| Under 6 years old | Beta(152.5, 2427.9) | 0.06 (0.05, 0.07) |
| Under 35 kg of weight | Beta(8.3, 359.6) | 0.02 (¡0.01, 0.04) |
| Late stage-2 disease | Beta(76.9, 44.9) | 0.63 (0.54, 0.72) |

Table D: Parameters for treatment eligibility.

##### S1.2.4 Health outcomes denominated as disability-adjusted life-years (DALYs)

As per recommendations of the Bill and Melinda Gates Foundation's reference case and WHO's guidelines for the conduct of cost-effectiveness analyses, we defined the utility of gHAT interventions in terms of disability-adjusted life-years (DALYs) [10–12]. DALYs were discounted at a rate of 3% per year [10, 12]. We follow established conventions to calculate DALYs and evaluate the estimates in present-day terms (after applying discounting) [10, 12, 13]. For the retrospective analysis, costs and DALYs are discounted to present-year values in 2014, the first year of the analysis, and for the prospective analysis present year values are expressed for the year 2021. For more details, see the supplementary information in Antillon *et. al*, Supplementary Note A, section 3 [9].

| Eligibility | Rationale | Summary |
|---|---|---|
| **Stage 1** | | |
| Pentamidine | Under 6 years old (1) | 0.06 (0.05, 0.07) |
| Fexinidazole-inpatient | Over 6 years old but under 35 kg of weight | 0.02 (¡0.01, 0.04) |
| Fexinidazole-outpatient | Over 6 years old and over 35 kg of weight | 0.92 (0.90, 0.93) |
| **Stage 2** | | |
| NECT | Under 6 years old or late-stage disease | 0.65 (0.57, 0.73) |
| Fexinidazole-inpatient | Over 6 years old but under 35 kg of weight and early stage-2 disease | ¡0.01 (¡0.01, 0.01) |
| Fexinidazole-outpatient | Over 6 years old, over 35 kg of weight, and early stage-2 disease | 0.34 (0.26, 0.42) |

[1] For simplicity, all patients over 6 years old were assumed to be over 20 kg in weight.

Table E: Eligibility for treatment

| Treatment | Outcomes | Estimate |
|---|---|---|
| **Stage 1** | | |
| Pentamidine | Cured | 0.05 (0.05, 0.06) |
| | Cured with SAEs | <0.01 (<0.01, <0.01) |
| | Rescue treatment | <0.01 (<0.01, <0.01) |
| | Death | <0.01 (<0.01, <0.01) |
| Fexinidazole - inpatient | Cured | 0.02 (<0.01, 0.04) |
| | Cured with SAEs | <0.01 (<0.01, <0.01) |
| | Rescue treatment | <0.01 (<0.01, <0.01) |
| | Death | <0.01 (<0.01, <0.01) |
| Fexinidazole - outpatient | Cured | 0.89 (0.87, 0.91) |
| | Cured with SAEs | 0.01 (<0.01, 0.02) |
| | Rescue treatment | 0.02 (<0.01, 0.03) |
| | Death | <0.01 (<0.01, <0.01) |
| **All treatments** | **Cured** | **0.97 (0.95, 0.98)** |
| | **Cured with SAEs** | **0.01 (<0.01, 0.03)** |
| | **Rescue treatment** | **0.02 (0.01, 0.03)** |
| | **Death** | **<0.01 (<0.01, <0.01)** |
| **Stage 2** | | |
| NECT | Cured | 0.56 (0.49, 0.64) |
| | Cured with SAEs | 0.06 (0.04, 0.08) |
| | Rescue treatment | 0.03 (0.01, 0.04) |
| | Death | <0.01 (<0.01, <0.01) |
| Fexinidazole - inpatient | Cured | <0.01 (<0.01, 0.01) |
| | Cured with SAEs | <0.01 (<0.01, <0.01) |
| | Rescue treatment | <0.01 (<0.01, <0.01) |
| | Death | <0.01 (<0.01, <0.01) |
| Fexinidazole - outpatient | Cured | 0.33 (0.25, 0.41) |
| | Cured with SAEs | <0.01 (<0.01, <0.01) |
| | Rescue treatment | <0.01 (<0.01, 0.01) |
| | Death | <0.01 (<0.01, <0.01) |
| **All treatments** | **Cured** | **0.90 (0.88, 0.92)** |
| | **Cured with SAEs** | **0.07 (0.05, 0.09)** |
| | **Rescue treatment** | **0.03 (0.02, 0.05)** |
| | **Death** | **<0.01 (<0.01, <0.01)** |

Table F: Treatments and outcomes distributions for stage 1 and 2 patients, calculated according to the probability tree in B. SAE: severe adverse events.

#### S1.3 Cost functions

The total costs are given by following expression:

$$\text{Total costs} = \sum_{i \in \text{all sub-categories}} \left( U_i \times C_i \right)$$

where
$i$ is the cost sub-category of cost (AS, PS, VC, or treatment)
$U$ is the unit of cost, which varies depending on the activity, such as people screened, teams deployed, fixed health centres outfitted with tests, etc.
$C$ is the cost per unit. All costs were denominated in 2020 US$.
$U_i$ or $C_i$, however, are in turn also functions, as described in the tables for AS (described in section S1.3.1), PS (described in section S1.3.2), VC (described in section S1.3.3), and treatment (described in section S1.3.4).

##### S1.3.1 Cost functions: active screening

The yearly costs of AS were calculated as a function of two groups of expenses: 1) overhead costs, and 2) the number of screening tests and confirmation tests that are used across all teams within the health zone. Because no synergies with other disease programs are believed to exist, we have employed a full costing method.

- Overhead costs: overhead costs are split between capital costs and recurrent costs to run an active screening team.
  - Capital costs consist of vehicles, medical equipment, energy (solar panels) and training (which occurs once every few years).
  - Recurrent costs consist of management and consumables that are spent on the team: fuel, staff time, etc.
- Costs that scale by population screened include:
  - CATT tests are scaled up according to the number of people that are screened per year, and a slight mark-up is included to account for wastage of CATT tests.
  - Confirmation tests are counted for all of those who are positive according to the CATT test, both the false positives (which are modelled according to the specificity of the test) and the true positives, which are the outputs of the dynamic model.
  - Lumbar punctures are not depicted as part of the AS costs, but are included as part of the treatment costs. Because many patients are eligible for fexinidazole treatment, which does not require lumbar punctures, we include lumbar puncture costs in the treatment portion of the analysis for those patients that are not eligible for fexinidazole. See section S1.3.4.

Briefly, we describe these parameters here, but they are displayed in more detail in tables A and D.

| Item | Units ($U$) | Cost ($C$) |
|---|---|---|
| Capital (annualized, traditional) | Proportion of patients handled by traditional team | AS capital (traditional team) |
| Capital (annualized, mini-team) | Proportion of patients handled by mini-team | AS capital (mini-team) |
| Management/recurrent expenses (traditional) | Proportion of patients handled by traditional team | AS recurrent (traditional) |
| Management/recurrent expenses (traditional | Proportion of patients handled by mini-team | AS recurrent (mini-teeam) |
| CATT testing (See Note 1) | AS coverage per year (traditional) × (1+wastage factors for CATT in AS context) | CATT × (1+delivery mark-up) |
| RDT testing (See Note 1) | AS coverage per year (traditional) × (1+wastage factors for RDT) | RDT × (1+delivery mark-up) |
| Microscopy/confirmation (traditional) | (1-CATT specificity) × (AS coverage per year × Population) | Microscopy |
| Microscopy/confirmation (mini-team) | (1-RDT specificity) × AS coverage per year (mini-team) × (1-LTFU mini-teams) | Microscopy |

[1] Ideally, CATT tests would be used for active screening and RDT tests would be used for passive screening because of the high wastage of CATT tests in the context of passive screening settings. In this context, RDT is also used for screening by mini-teams (motorcycle teams) as it lightens the load of supplies necessary for motorcycle teams to take.

Table G: Active screening: cost function

| Variable Name | Parameterization | Summary |
|---|---|---|
| AS coverage per year (traditional team) | Fixed | See Table A |
| AS coverage per year (motorcycle team) | Fixed | See Table B |
| Wastage factor for CATT administration in AS context | Beta(8, 92) | 0.08 (0.04, 0.14) |
| Wastage factor for RDT | Beta(1, 99) | 0.01 (¡0.01, 0.04) |
| CATT specificity | Beta(31, 2) | 1.00 (1.00, 1.00) |
| RDT specificity | Beta(3886, 24) | 0.99 (0.99, 1.00) |
| AS capital costs (annualized, traditional) | Gamma(20000, 0.02) | 4,000 (3,982, 4,018) |
| AS recurrent costs (annualized, traditional) | Gamma(8.475, 2167) | 18,503 (8,234, 32,348) |
| AS capital costs (annualized, motorcycle) | Gamma(8.475, 1000.81) | 8,527 (3,828, 15,216) |
| AS recurrent costs (annualized, motorcycle) | Gamma(70.05, 71.56) | 6,424 (5,031, 8,016) |
| Cost of CATT test | Gamma(23, 0.02) | 0.46 (0.29, 0.66) |
| Cost of RDT test | Gamma(8.475, 0.19) | 1.60 (0.70, 2.85) |
| Pr. lost-to-follow-up, RDT+ suspect | Beta, alpha = LTFU, beta = Sero+ | See Table B |
| Cost confirmation (microscopy) | Gamma(8.475, 1.27) | 10.69 (4.92, 18.89) |
| Cost of delivery (markup) | Beta(45, 55) | 0.20 (0.15, 0.25) |

Table H: Components of active screening costs. Full citations and explanations for the parameters will be given in Supplementary Information SI Text 4.

The cost per year, given the number of people screened, is therefore:

Table I: Cost breakdown for active screening activities

| Item | Units ($U$) | Cost per unit ($C$) | Cost per category |
|---|---|---|---|
| **Retrospective AS, 2014** | | | |
| Capital (annualized, traditional) | 1 | 4,000 (3,982, 4,018) | 4,000 (3,982, 4,018) |

Table I: Cost breakdown for active screening activities *(continued)*

| Item | Units ($U$) | Cost per unit ($C$) | Cost per category |
|---|---|---|---|
| Capital (annualized, mini-team) | 0 | 8,527 (3,828, 15,216) | 0 |
| Recurrent expenses (traditional) | 1 | 18,503 (8,234, 32,348) | 18,503 (8,234, 32,348) |
| Recurrent expenses (mini-team) | 0 | 6,424 (5,031, 8,016) | 0 |
| Microscopy (traditional) | 77.46 (45.89, 108.99) | 10.69 (4.92, 18.89) | 827 (311, 1,700) |
| Microscopy (mini-team) | 0 | 10.69 (4.92, 18.89) | 0 |
| CATT testing | 24,094 (22,976, 25,694) | 0.55 (0.35, 0.80) | 13,243 (8,351, 19,226) |
| RDT testing | 0 | 1.93 (0.83, 3.43) | 0 |
| **Subtotal - Traditional** | | | **36,574 (24,905, 51,666)** |
| **Subtotal - Mini-team** | | | **0** |
| **Total** | | | **36,574 (24,905, 51,666)** |
| **Retrospective AS, 2015** | | | |
| Capital (annualized, traditional) | 0.66 | 4,000 (3,982, 4,018) | 2,640 (2,628, 2,652) |
| Capital (annualized, mini-team) | 0.92701 | 8,527 (3,828, 15,216) | 7,904 (3,549, 14,105) |
| Recurrent expenses (traditional) | 0.66 | 18,503 (8,234, 32,348) | 12,212 (5,435, 21,350) |
| Recurrent expenses (mini-team) | 0.92701 | 6,424 (5,031, 8,016) | 5,955 (4,664, 7,431) |
| Microscopy (traditional) | 62.94 (37.29, 88.56) | 10.69 (4.92, 18.89) | 672 (253, 1,382) |
| Microscopy (mini-team) | 53.75 (34.56, 76.66) | 10.69 (4.92, 18.89) | 575 (233, 1,126) |
| CATT testing | 19,578 (18,669, 20,878) | 0.55 (0.35, 0.80) | 10,760 (6,785, 15,622) |
| RDT testing | 9,366 (9,272, 9,629) | 1.93 (0.83, 3.43) | 18,033 (7,779, 32,103) |
| **Subtotal - Traditional** | | | **26,285 (18,136, 36,684)** |
| **Subtotal - Mini-team** | | | **32,468 (20,758, 47,996)** |
| **Total** | | | **58,753 (43,894, 77,049)** |
| **Retrospective AS, 2016** | | | |
| Capital (annualized, traditional) | 0.51 | 4,000 (3,982, 4,018) | 2,040 (2,031, 2,049) |
| Capital (annualized, mini-team) | 1.078343 | 8,527 (3,828, 15,216) | 9,195 (4,128, 16,408) |
| Recurrent expenses (traditional) | 0.51 | 18,503 (8,234, 32,348) | 9,437 (4,200, 16,498) |
| Recurrent expenses (mini-team) | 1.078343 | 6,424 (5,031, 8,016) | 6,928 (5,425, 8,644) |
| Microscopy (traditional) | 39.26 (23.26, 55.24) | 10.69 (4.92, 18.89) | 419 (158, 862) |
| Microscopy (mini-team) | 62.48 (40.32, 88.96) | 10.69 (4.92, 18.89) | 669 (273, 1,317) |
| CATT testing | 12,211 (11,644, 13,022) | 0.55 (0.35, 0.80) | 6,711 (4,232, 9,744) |
| RDT testing | 10,895 (10,786, 11,201) | 1.93 (0.83, 3.43) | 20,977 (9,049, 37,344) |
| **Subtotal - Traditional** | | | **18,607 (12,669, 26,295)** |
| **Subtotal - Mini-team** | | | **37,768 (24,158, 55,847)** |
| **Total** | | | **56,375 (41,263, 76,024)** |
| **Retrospective AS, 2017** | | | |
| Capital (annualized, traditional) | 0.62 | 4,000 (3,982, 4,018) | 2,480 (2,469, 2,491) |
| Capital (annualized, mini-team) | 0.689472 | 8,527 (3,828, 15,216) | 5,879 (2,639, 10,491) |

Table I: Cost breakdown for active screening activities *(continued)*

| Item | Units ($U$) | Cost per unit ($C$) | Cost per category |
|---|---|---|---|
| Recurrent expenses (traditional) | 0.62 | 18,503 (8,234, 32,348) | 11,472 (5,105, 20,056) |
| Recurrent expenses (mini-team) | 0.689472 | 6,424 (5,031, 8,016) | 4,429 (3,469, 5,527) |
| Microscopy (traditional) | 39.35 (23.31, 55.36) | 10.69 (4.92, 18.89) | 420 (158, 864) |
| Microscopy (mini-team) | 17.94 (11.11, 26.85) | 10.69 (4.92, 18.89) | 192.04 (76.01, 384.65) |
| CATT testing | 12,239 (11,671, 13,051) | 0.55 (0.35, 0.80) | 6,727 (4,242, 9,766) |
| RDT testing | 6,966 (6,896, 7,161) | 1.93 (0.83, 3.43) | 13,412 (5,786, 23,877) |
| **Subtotal - Traditional** | | | **21,099 (14,086, 30,302)** |
| **Subtotal - Mini-team** | | | **23,913 (15,234, 35,263)** |
| **Total** | | | **45,012 (33,362, 59,454)** |
| **Retrospective AS, 2018** | | | |
| Capital (annualized, traditional) | 0.59 | 4,000 (3,982, 4,018) | 2,360 (2,350, 2,370) |
| Capital (annualized, mini-team) | 0.741403 | 8,527 (3,828, 15,216) | 6,322 (2,838, 11,281) |
| Recurrent expenses (traditional) | 0.59 | 18,503 (8,234, 32,348) | 10,917 (4,858, 19,085) |
| Recurrent expenses (mini-team) | 0.741403 | 6,424 (5,031, 8,016) | 4,763 (3,730, 5,943) |
| Microscopy (traditional) | 37.32 (22.11, 52.51) | 10.69 (4.92, 18.89) | 399 (150, 819) |
| Microscopy (mini-team) | 29.51 (18.67, 42.79) | 10.69 (4.92, 18.89) | 316 (126, 624) |
| CATT testing | 11,608 (11,069, 12,378) | 0.55 (0.35, 0.80) | 6,380 (4,023, 9,262) |
| RDT testing | 7,491 (7,416, 7,701) | 1.93 (0.83, 3.43) | 14,422 (6,221, 25,675) |
| **Subtotal - Traditional** | | | **20,055 (13,387, 28,798)** |
| **Subtotal - Mini-team** | | | **25,823 (16,471, 38,176)** |
| **Total** | | | **45,878 (33,933, 60,790)** |
| **Retrospective AS, 2019** | | | |
| Capital (annualized, traditional) | 0.91 | 4,000 (3,982, 4,018) | 3,640 (3,624, 3,656) |
| Capital (annualized, mini-team) | 0.11376 | 8,527 (3,828, 15,216) | 970 (435, 1,731) |
| Recurrent expenses (traditional) | 0.91 | 18,503 (8,234, 32,348) | 16,838 (7,493, 29,437) |
| Recurrent expenses (mini-team) | 0.11376 | 6,424 (5,031, 8,016) | 731 (572, 912) |
| Microscopy (traditional) | 40.23 (23.84, 56.61) | 10.69 (4.92, 18.89) | 430 (162, 883) |
| Microscopy (mini-team) | 5.41 (3.40, 7.94) | 10.69 (4.92, 18.89) | 57.85 (23.30, 115.16) |
| CATT testing | 12,514 (11,933, 13,345) | 0.55 (0.35, 0.80) | 6,878 (4,337, 9,986) |
| RDT testing | 1,149 (1,138, 1,182) | 1.93 (0.83, 3.43) | 2,213 (955, 3,940) |
| **Subtotal - Traditional** | | | **27,786 (17,959, 40,727)** |
| **Subtotal - Mini-team** | | | **3,972 (2,539, 5,873)** |
| **Total** | | | **31,758 (21,950, 44,789)** |
| **Retrospective AS, 2020 and later, prospective mean AS, and prospective max AS 2020-2022** | | | |
| Capital (annualized, traditional) | 0.69 | 4,000 (3,982, 4,018) | 2,760 (2,748, 2,772) |
| Capital (annualized, mini-team) | 0.608468 | 8,527 (3,828, 15,216) | 5,188 (2,329, 9,258) |
| Recurrent expenses (traditional) | 0.69 | 18,503 (8,234, 32,348) | 12,767 (5,682, 22,320) |

Table I: Cost breakdown for active screening activities *(continued)*

| Item | Units ($U$) | Cost per unit ($C$) | Cost per category |
|---|---|---|---|
| Recurrent expenses (mini-team) | 0.608468 | 6,424 (5,031, 8,016) | 3,909 (3,061, 4,878) |
| Microscopy (traditional) | 47.37 (28.07, 66.65) | 10.69 (4.92, 18.89) | 506 (190, 1,040) |
| Microscopy (mini-team) | 35.64 (22.65, 51.00) | 10.69 (4.92, 18.89) | 382 (154, 759) |
| CATT testing | 14,735 (14,051, 15,713) | 0.55 (0.35, 0.80) | 8,099 (5,107, 11,758) |
| RDT testing | 6,148 (6,086, 6,320) | 1.93 (0.83, 3.43) | 11,836 (5,106, 21,072) |
| **Subtotal - Traditional** | | | **24,132 (16,252, 34,374)** |
| **Subtotal - Mini-team** | | | **21,315 (13,643, 31,474)** |
| **Total** | | | **45,447 (33,967, 59,499)** |
| **Prospective max AS, 2023 and later** | | | |
| Capital (annualized, traditional) | 0.69 | 4,000 (3,982, 4,018) | 2,760 (2,748, 2,772) |
| Capital (annualized, mini-team) | 0.845215 | 8,527 (3,828, 15,216) | 7,207 (3,236, 12,861) |
| Recurrent expenses (traditional) | 0.69 | 18,503 (8,234, 32,348) | 12,767 (5,682, 22,320) |
| Recurrent expenses (mini-team) | 0.845215 | 6,424 (5,031, 8,016) | 5,430 (4,252, 6,776) |
| Microscopy (traditional) | 65.80 (38.99, 92.59) | 10.69 (4.92, 18.89) | 703 (264, 1,444) |
| Microscopy (mini-team) | 39.93 (23.42, 60.26) | 10.69 (4.92, 18.89) | 427 (162, 853) |
| CATT testing | 20,468 (19,518, 21,827) | 0.55 (0.35, 0.80) | 11,250 (7,094, 16,332) |
| RDT testing | 8,540 (8,454, 8,779) | 1.93 (0.83, 3.43) | 16,442 (7,092, 29,270) |
| **Subtotal - Traditional** | | | **27,480 (18,960, 38,351)** |
| **Subtotal - Mini-team** | | | **29,506 (18,828, 43,523)** |
| **Total** | | | **56,986 (42,745, 74,299)** |

##### S1.3.2 Cost functions: passive screening (screening at fixed health posts)

The yearly costs of PS were calculated as a function of two groups of expenses: 1) overhead costs, and 2) the number of consultations and screening and confirmation tests that are done in the focus.

- The previously-operating screening & diagnostic center (the Catholic Mission Hospital) is part of the default activities. One screening & confirmation center (Bodo District hospital) is part of the enhanced activities. All "screening-only" centers are part of the *Improved* or *enhanced* PS activities.
- Overhead costs: overhead costs are split between capital costs and recurrent costs to equip a health center to perform serological confirmation for HAT.
  - Capital costs consist of medical equipment, energy (e.g. solar panels) and training (which occurs periodically every few years). These costs are scaled by the number of facilities that can perform serological screening.
  - Recurrent costs consist of management at the national level. As of 2020, since only 23 out of the national 54 clinics are in Mandoul, we attributed 23/54 $\times$ \$6000 (the cost of management nationally) to the focus.
- Costs that scale by population screened include:
  - RDT tests are scaled up according to the number of people that are screened per year, and a slight mark-up is included to account for wastage of tests.
  - Confirmation tests are counted for all of those who are RDT+: both the false positives (which are modelled as a factor equal to the specificity of the test) and the true positives, which are the outputs of the dynamic model.
  - Lumbar punctures are not depicted as part of the passive surveillance diagnosis costs, but are included as part of the treatment costs. Because many patients are eligible for fexinidazole treatment, which does not require lumbar punctures, we include lumbar puncture costs in the treatment portion of the analysis for those patients that are not eligible for fexinidazole. See section S1.3.4
- Loop-mediated Isothermal Amplification (LAMP) was only available at the Bodo hospital and was performed on very patients per year, and due to an absence of cost estimates, no additional costs were imputed for this.
- Full citations and explanations for the parameters will be given in Supplementary Information S4 Text.

| Item | Units ($U$) | Cost ($C$) |
|---|---|---|
| Capital (annualized) - screening and confirmation sites | Number of facilities capable of screening and confirmation within the focus | Capital costs (clinic) |
| Capital (annualized) - screening only sites | Number of facilities capable of screening only within the focus | Capital costs (RDT clinic) |
| District management | Per district | District management costs |
| District management - expanded network | Per district | District management costs for expanded network |
| Consultation - screening and confirmation sites | PS coverage per year per clinic × Clinics in the focus | Consultation cost |
| Consultation - screening only sites | PS coverage per year per clinic × Clinics in the focus | Consultation cost |
| CATT testing - screening and confirmation sites | PS coverage per year per clinic × Clinics in the focus × (1+wastage for CATT in PS context) | CATT × (1+delivery mark-up) |
| RDT testing - screening only sites | PS coverage per year per clinic × Clinics in the focus × (1+wastage for RDT) | RDT × (1+delivery mark-up) |
| Microscopy/confirmation (suspects first identified in screening and confirmation sites) | (1-CATT specificity) × (PS coverage per year per clinic × Clinics in the focus) | Microscopy |
| Microscopy/confirmation (suspects first identified in screening-only sites) | (1-RDT specificity) × (1-Pr. LTFU) × (PS coverage per year per clinic × Clinics in the focus) | Microscopy |

[1] NA

Table J: Passive screening: cost function

| Variable Name | Parameterization | Summary |
|---|---|---|
| AS coverage per year (traditional team) | Fixed | See Table A |
| AS coverage per year (motorcycle team) | Fixed | See Table B |
| Wastage factor for CATT administration in AS context | Beta(8, 92) | 0.08 (0.03, 0.14) |
| Wastage factor for RDT | Beta(1, 99) | 0.01 (¡0.01, 0.04) |
| CATT specificity | Beta(31, 2) | 1.00 (1.00, 1.00) |
| RDT specificity | Beta(3886, 24) | 0.99 (0.99, 1.00) |
| AS capital costs (annualized, traditional) | Gamma(20000, 0.02) | 4,000 (3,982, 4,018) |
| AS recurrent costs (annualized, traditional) | Gamma(8.475, 2167) | 18,200 (8,048, 32,654) |
| AS capital costs (annualized, motorcycle) | Gamma(8.475, 1000.81) | 8,539 (3,864, 15,185) |
| AS recurrent costs (annualized, motorcycle) | Gamma(70.05, 71.56) | 6,409 (4,975, 7,989) |
| Cost of CATT test | Gamma(23, 0.02) | 0.46 (0.29, 0.67) |
| Cost of RDT test | Gamma(8.475, 0.19) | 1.61 (0.73, 2.88) |
| Pr. lost-to-follow-up, RDT+ suspect | Year-specific Beta distribution, alpha = LTFU, beta = Sero+ in Table B | See Table B |
| Cost confirmation (microscopy) | Gamma(8.475, 1.27) | 10.64 (4.78, 18.68) |
| Cost of delivery (markup) | Beta(45, 55) | 0.20 (0.15, 0.25) |

Table K: Components of passive screening costs. Full citations and explanations for the parameters will be given in Supplementary Information SI Text 4.

The cost per year, given the number of people screened and the number of health centres available for PS, is shown in table L.

Table L: Cost breakdown for passive screening activities

| Item | Units ($U$) | Cost per unit ($C$) | Cost per category |
|---|---|---|---|
| **Retrospective PS, 2014** | | | |
| Capital - full clinic | 1 | 1,822 (811, 3,208) | 1,822 (811, 3,208) |
| Capital - RDT clinic | 0 | 271 (120, 477) | 0 |
| Management | 1 | 396 (272, 545) | 396 (272, 545) |
| OP visit - default | 100 | 2 (0, 5) | 196.81 (32.21, 505.83) |
| OP visit - enhanced | 0 | 1.97 (0.32, 5.06) | 0 |
| CATT | 125 (117, 134) | 0.55 (0.35, 0.80) | 68.96 (42.90, 100.84) |
| RDT | 0 | 1.94 (0.87, 3.46) | 0 |
| Microscopy for false positives - default | 0.35 (0.21, 0.49) | 10.64 (4.78, 18.68) | 3.74 (1.37, 7.54) |
| Microscopy for false positives - enhanced | 0 | 10.64 (4.78, 18.68) | 0 |
| **Subtotal - default** | | | **1,379 (817, 2,118)** |
| **Subtotal - enhanced** | | | **1,109 (600, 1,809)** |
| **Total** | | | **2,488 (1,433, 3,935)** |
| **Retrospective PS, 2015** | | | |
| Capital - full clinic | 2 | 1,822 (811, 3,208) | 3,644 (1,622, 6,417) |
| Capital - RDT clinic | 7 | 271 (120, 477) | 1,895 (837, 3,341) |
| Management | 9 | 396 (272, 545) | 3,567 (2,445, 4,908) |
| OP visit - default | 119 | 2 (0, 5) | 234.20 (38.33, 601.94) |
| OP visit - enhanced | 774 | 1.97 (0.32, 5.06) | 1,523 (249, 3,915) |
| CATT | 149 (139, 159) | 0.55 (0.35, 0.80) | 82.07 (51.05, 120.00) |
| RDT | 782 (774, 803) | 1.94 (0.87, 3.46) | 1,514 (678, 2,711) |
| Microscopy for false positives - default | 0.42 (0.25, 0.59) | 10.64 (4.78, 18.68) | 4.45 (1.63, 8.97) |
| Microscopy for false positives - enhanced | 657 (553, 734) | 10.64 (4.78, 18.68) | 6,987 (3,050, 12,345) |
| **Subtotal - default** | | | **2,539 (1,479, 3,983)** |
| **Subtotal - enhanced** | | | **16,912 (11,928, 23,058)** |
| **Total** | | | **19,451 (14,006, 26,105)** |
| **Retrospective PS, 2016** | | | |
| Capital - full clinic | 2 | 1,822 (811, 3,208) | 3,644 (1,622, 6,417) |
| Capital - RDT clinic | 7 | 271 (120, 477) | 1,895 (837, 3,341) |
| Management | 9 | 396 (272, 545) | 3,567 (2,445, 4,908) |
| OP visit - default | 128 | 2 (0, 5) | 251.92 (41.23, 647.46) |
| OP visit - enhanced | 1251 | 1.97 (0.32, 5.06) | 2,462 (403, 6,328) |
| CATT | 160 (150, 171) | 0.55 (0.35, 0.80) | 88.27 (54.91, 129.07) |
| RDT | 1,264 (1,251, 1,297) | 1.94 (0.87, 3.46) | 2,446 (1,095, 4,381) |
| Microscopy for false positives - default | 0.45 (0.27, 0.63) | 10.64 (4.78, 18.68) | 4.79 (1.75, 9.65) |
| Microscopy for false positives - enhanced | 1,211 (1,107, 1,250) | 10.64 (4.78, 18.68) | 12,888 (5,658, 22,843) |
| **Subtotal - default** | | | **2,563 (1,493, 4,008)** |
| **Subtotal - enhanced** | | | **24,684 (16,438, 35,280)** |
| **Total** | | | **27,247 (18,708, 38,150)** |
| **Retrospective PS, 2017** | | | |
| Capital - full clinic | 2 | 1,822 (811, 3,208) | 3,644 (1,622, 6,417) |
| Capital - RDT clinic | 28 | 271 (120, 477) | 7,579 (3,346, 13,364) |
| Management | 30 | 396 (272, 545) | 11,890 (8,148, 16,360) |
| OP visit - default | 101 | 2 (0, 5) | 198.78 (32.53, 510.89) |
| OP visit - enhanced | 2100 | 1.97 (0.32, 5.06) | 4,133 (676, 10,622) |
| CATT | 126 (118, 135) | 0.55 (0.35, 0.80) | 69.65 (43.33, 101.85) |

Table L: Cost breakdown for passive screening activities *(continued)*

| Item | Units ($U$) | Cost per unit ($C$) | Cost per category |
|---|---|---|---|
| RDT | 2,121 (2,101, 2,178) | 1.94 (0.87, 3.46) | 4,107 (1,839, 7,355) |
| Microscopy for false positives - default | 0.35 (0.21, 0.50) | 10.64 (4.78, 18.68) | 3.78 (1.38, 7.62) |
| Microscopy for false positives - enhanced | 2,030 (1,847, 2,098) | 10.64 (4.78, 18.68) | 21,598 (9,529, 38,157) |
| **Subtotal - default** | | | **2,491 (1,436, 3,936)** |
| **Subtotal - enhanced** | | | **50,732 (35,695, 69,613)** |
| **Total** | | | **53,223 (37,957, 72,207)** |
| **Retrospective PS, 2018** | | | |
| Capital - full clinic | 2 | 1,822 (811, 3,208) | 3,644 (1,622, 6,417) |
| Capital - RDT clinic | 28 | 271 (120, 477) | 7,579 (3,346, 13,364) |
| Management | 30 | 396 (272, 545) | 11,890 (8,148, 16,360) |
| OP visit - default | 55 | 2 (0, 5) | 108.25 (17.72, 278.21) |
| OP visit - enhanced | 2160 | 1.97 (0.32, 5.06) | 4,251 (696, 10,926) |
| CATT | 68.76 (64.46, 73.63) | 0.55 (0.35, 0.80) | 37.93 (23.59, 55.46) |
| RDT | 2,182 (2,161, 2,240) | 1.94 (0.87, 3.46) | 4,224 (1,891, 7,565) |
| Microscopy for false positives - default | 0.19 (0.11, 0.27) | 10.64 (4.78, 18.68) | 2.06 (0.75, 4.15) |
| Microscopy for false positives - enhanced | 1,445 (994, 1,828) | 10.64 (4.78, 18.68) | 15,377 (6,362, 28,363) |
| **Subtotal - default** | | | **2,367 (1,347, 3,791)** |
| **Subtotal - enhanced** | | | **44,746 (32,269, 60,615)** |
| **Total** | | | **47,113 (34,469, 63,145)** |
| **Retrospective PS, 2019** | | | |
| Capital - full clinic | 2 | 1,822 (811, 3,208) | 3,644 (1,622, 6,417) |
| Capital - RDT clinic | 20 | 271 (120, 477) | 5,413 (2,390, 9,546) |
| Management | 22 | 396 (272, 545) | 8,719 (5,976, 11,997) |
| OP visit - default | 82 | 2 (0, 5) | 161.38 (26.41, 414.78) |
| OP visit - enhanced | 2464 | 1.97 (0.32, 5.06) | 4,849 (794, 12,464) |
| CATT | 102.52 (96.11, 109.78) | 0.55 (0.35, 0.80) | 56.55 (35.18, 82.69) |
| RDT | 2,489 (2,465, 2,555) | 1.94 (0.87, 3.46) | 4,818 (2,158, 8,629) |
| Microscopy for false positives - default | 0.29 (0.17, 0.40) | 10.64 (4.78, 18.68) | 3.07 (1.12, 6.18) |
| Microscopy for false positives - enhanced | 1,421 (556, 2,174) | 10.64 (4.78, 18.68) | 15,104 (4,628, 31,654) |
| **Subtotal - default** | | | **2,439 (1,401, 3,884)** |
| **Subtotal - enhanced** | | | **40,330 (26,899, 58,814)** |
| **Total** | | | **42,770 (29,030, 61,594)** |
| **Retrospective PS, 2020 and later** | | | |
| Capital - full clinic | 2 | 1,822 (811, 3,208) | 3,644 (1,622, 6,417) |
| Capital - RDT clinic | 20 | 271 (120, 477) | 5,413 (2,390, 9,546) |
| Management | 22 | 396 (272, 545) | 8,719 (5,976, 11,997) |
| OP visit - default | 39 | 2 (0, 5) | 76.76 (12.56, 197.27) |
| OP visit - enhanced | 1320 | 1.97 (0.32, 5.06) | 2,598 (425, 6,677) |
| CATT | 48.76 (45.71, 52.21) | 0.55 (0.35, 0.80) | 26.90 (16.73, 39.33) |
| RDT | 1,333 (1,320, 1,369) | 1.94 (0.87, 3.46) | 2,581 (1,156, 4,623) |
| Microscopy for false positives - default | 0.14 (0.08, 0.19) | 10.64 (4.78, 18.68) | 1.46 (0.53, 2.94) |
| Microscopy for false positives - enhanced | 853 (549, 1,121) | 10.64 (4.78, 18.68) | 9,077 (3,692, 17,178) |
| **Subtotal - default** | | | **2,324 (1,308, 3,732)** |
| **Subtotal - enhanced** | | | **29,814 (21,620, 40,291)** |

Table L: Cost breakdown for passive screening activities *(continued)*

| Item | Units ($U$) | Cost per unit ($C$) | Cost per category |
|---|---|---|---|
| **Total** | | | **32,138 (23,594, 42,762)** |

##### S1.3.3 Cost functions: vector control

The costs of vector control activities for a year cost $56,000 according to a micro-costing effort published in [14] in 2016 USD. When we account for inflation and changes in the exchange of the Central African Franc and the US dollar, we estimate that the equivalent in 2020 USD is 61,238. We applied a gamma distribution in which the 95% CI would span half and double the estimate: Gamma(8.475, rate: 8132). The resulting mean is 68,717 and the 95% CI is 29,881-123,615.

##### S1.3.4 Cost functions: treatment

| Item | Units ($U$) | Cost ($C$) |
|---|---|---|
| Doctor's consult | All patients | Outpatient consult |
| Staging cases (supplies and time); patients ineligible for fexinidazole treatment (see Notes 1-2). | Patients in both stages of disease ineligible for fexinidazole treatment. | Lumbar puncture cost |
| Pentamidine (see Notes 1-3). | Cases of stage 1 disease detected with AS or PS × proportion of patients ineligible for fexinidazole. | Pentamidine × (1+delivery mark-up) |
| Outpatient care for stage 1 with pentamidine (see Notes 1-3). | Cases of stage 1 disease detected with AS or PS × proportion of patients ineligible for fexinidazole × length of treatment for pentamidine. | Outpatient consult |
| NECT (see Notes 1-3). | Cases of stage 2 disease detected with AS or PS × proportion of patients ineligible for fexinidazole. | NECT × (1+delivery mark-up) |
| Inpatient care for stage 2 with NECT (see Notes 1-3). | (Cases stage 2 detected with AS or PS) × proportion of patients ineligible for fexinidazole × length of treatment for NECT. | Inpatient cost per day |
| Fexinidazole (see Notes 1-2). | Patients in both stages of disease eligible for fexinidazole treatment. | Fexinidazole × (1+delivery mark-up) |
| Inpatient care for either stage 1 or 2 with fexinidazole (see Notes 1-3) | Patients eligible for fexinidazole on an inpatient basis × length of treatment for fexinidazole. | Inpatient cost per day |
| Outpatient care for either stage 1 or 2 with fexinidazole (see Notes 1-4). | Patients eligible for outpatient treatment. | Outpatient consult |
| Treatment for severe adverse events | Patients under each treatment who experience severe adverse events | (Outpatient consult + Inpatient cost per day × length of treatment for severe adverse events) |
| Microscopy confirmation | All S1, S2 confirmed patients | Microscopy |

[1] Fexinidazole was only available in Chad after 2020. See WHO treatment recommendations for eligibility for fexinidazole. While some patients are eligible for fexinidazole (over 6 years of age and below the severe threshold of disease) the recommendations stipulate inpatient care for some patients due to low weight.
[2] The proportion of patients eligible for fexinidazole was determined as follows: (1-proportion of patients under 6 years of age) × (1-proportion of patients with signs of late stage 2 disease).
[3] The proportion of patients who had fexinidazole treatment on an inpatient basis was determined by multiplying the equation in note 2 with the proportion of patients who were over 35 kg of weight.
[4] Fexinidazole is currently recommended on an outpatient basis only for some patients and only as a directly-observed therapy, so we have imputed a cost for the daily administration by a village health worker. For simplicity, we have given this the same value as a regular outpatient visit since it constitutes a small portion of all costs.

Table M: Treatment: cost function

We show here the components of the costs per case treated depending on the stage and the treatment. The parameters for the above table are available in section S4.9 and eligibility distributions are described in table E.

| Variable.Name | Parameterization | Summary |
|---|---|---|
| Lumbar puncture and laboratory exam - cost | Gamma(2.42, 3.66) | 8.81 (1.33, 22.51) |
| Duration of hospital stay for NECT treatment in days | Fixed | 10 |
| Duration of hospital stay fexinidazole for stage 1 or 2 disease in days | Fixed | 10 |
| Duration of severe adverse events in days | Gamma (1.219, 2.377) | 2.89 (0.13, 9.94) |
| Probability of serious adverse events - pentamidine | Beta(1,499) | 0.0026 (0.0002, 0.0081) |
| Probability of serious adverse events - NECT | Beta(11.6,226.4) | 0.10 (0.07, 0.13) |
| Probability of serious adverse events - fexinidazole | Beta(3,261) | 0.01 (¡0.01, 0.03) |
| Outpatient consultation - cost | Gamma(2.48,0.79) | 1.97 (0.32, 5.06) |
| Hospital day - cost | Gamma(5.45,1.76) | 9.52 (3.28, 19.34) |
| Course of pentamidine - cost | Fixed | 54 |
| Course of NECT - cost | Fixed | 360 |
| Course of fexinidazole - cost | Fixed | 50 |
| Delivery mark-up | Beta(45,55) | 0.20 (0.15, 0.25) |

Table N: Parameters for treatment costs. Full citations and explanations for the parameters will be given in Supplementary Information SI Text 4.

| | Pentamidine | NECT | Fexinidazole - inpatient | Fexinidazole - outpatient |
|---|---|---|---|---|
| Staging | 8.81 (1.33, 22.51) | 8.81 (1.33, 22.51) | 0 | 0 |
| Doctor's consult | 19.68 (3.22, 50.58) | 1.97 (0.32, 5.06) | 1.97 (0.32, 5.06) | 19.68 (3.22, 50.58) |
| Inpatient care | 0 | 95.17 (32.75, 193.45) | 95.17 (32.75, 193.45) | 0 |
| Medicine | 64.80 (62.26, 67.35) | 431.99 (415.07, 448.98) | 60.00 (57.65, 62.36) | 60.00 (57.65, 62.36) |
| Treatment for SAE | <0.01 (<0.01, 0.02) | 2.95 (0.24, 11.39) | 0.34 (0.02, 1.58) | 0.34 (0.02, 1.58) |
| **Total** | **93.29 (72.85, 126.87)** | **540.88 (471.90, 644.90)** | **157.47 (94.95, 256.41)** | **79.70 (62.77, 110.32)** |

Table O: Cost per person for different gHAT treatments. Because these are costs averaged over all patients and SAEs are rare, the average cost per patient for SAE is low.
