## Supplementary material for "Health economic evaluation of strategies to eliminate *gambiense* human African trypanosomiasis in the Mandoul disease focus of Chad": S2 Text. Glossary of Technical Terms

### S2. Glossary of Technical Terms

Box 1: Glossary (adapted from Antillon et. al 2022 [1] under a CC-BY 4.0 license.)

#### Epidemiology Terms

**Intervention** Interventions are separate activities to address a health need (e.g. active screening (AS) or vector control (VC)).

**Strategy** A strategy is a combination of interventions, carried out with a specific coverage, and in parallel. In this paper, we simulate strategies with and without an improvement in PS and with and without VC.

**Elimination of transmission (EoT)** Globally this is the 2030 goal for gHAT; here we also consider local EoT for health zones. The feasibility of EoT is expressed as a probability equal to the proportion of our simulations in which new infections is zero before a given year (usually 2030).

**Disability-adjusted life-year (DALY)** In order to present the burden of disease in one common metric across diseases, DALYs are calculated in cost-effectiveness analyses. This is the sum of the years lived with disability due to the disease and the years of life lost by fatal cases. See section **??**.

#### Health Economics Terms

**Parameter uncertainty** Uncertainty in the level of transmission or in the costs of interventions and treatment due to unknown underlying parameters (see supplementary section **??** for an explanation of our parameterization of the health outcomes and cost model).

**Willingness-to-pay (WTP) or cost-effectiveness threshold** The amount of money that payers would pay to avert one DALY arising from the disease in the analysis (gHAT). No specific threshold is recommended, but a recent analysis shows that the WTP in Chad is between $30–518 in 2020 USD per DALY averted [2–4].

**Incremental cost-effectiveness ratio** A ratio of marginal cost for a marginal benefit, calculated as follows:

$$\text{ICER} = \frac{\Delta\text{Costs}}{\Delta\text{DALYs}} = \frac{\text{Costs}_{\text{strategy}} - \text{Costs}_{\text{next best}}}{\text{Effects}_{\text{strategy}} - \text{Effects}_{\text{next best}}}$$

Not that the ICER is not a strategy against the comparator, but a strategy against the *next best* strategy in terms of costs incurred and DALYs averted.

**Cost-effective strategy** The strategy where the ICER is less than the WTP (or cost-effectiveness threshold). We say that the cost-effective strategy is "conditional" on the WTP.

**Dominated strategy** A strategy that costs more than another while yielding worse health outcomes as determined by the DALY calculations. This strategy ought not be implemented.

**Weakly dominated strategy (or strategies under extended dominance)** A strategy in which the ICER is higher that the next more expensive strategy. Unlike a strongly dominated strategy it costs less than the most expensive strategy, but the cost-per-DALY averted is higher. This strategy ought not be implemented.

**Net monetary benefit** The net benefits (NMB) framework is derived from ICERs, but also takes uncertainty into account.

$$\text{NMB} \mid \text{WTP} : \text{WTP} \times \Delta\text{DALYs} - \Delta\text{Costs}$$

The optimal strategy at a given WTP is the strategy with the highest mean NMB at that value of WTP.

**Optimal strategy** Analogous to the cost-effective strategy when no uncertainty is assumed, this is the strategy that is recommended by the NMB framework.
