## Supplementary material for "Health economic evaluation of strategies to eliminate *gambiense* human African trypanosomiasis in the Mandoul disease focus of Chad": S3 Text. Supplemental Results.

### S3. Supplementary Results

#### Contents


S3.1 Supplemental results for the retrospective analysis . . . . . . . . . . . . . . . . . . . . . . . . S3.2
S3.2 Supplemental results for the prospective analysis . . . . . . . . . . . . . . . . . . . . . . . . . S3.4
S3.3 Sensitivity analysis of time horizon and discounting rate . . . . . . . . . . . . . . . . . . . . . S3.6
S3.4 Sensitivity analysis: results if fexinidazole had been available . . . . . . . . . . . . . . . . . . . S3.7
S3.5 Parameter-specific uncertainty analysis (retrospective analysis) . . . . . . . . . . . . . . . . . . S3.8


#### List of Figures


A Disability adjusted life-years (DALYs) accrued through time for different counterfactual strategies in the retrospective analysis. . . . . . . . . . . . . . . . . . . . . . . . . . . . . . . . . . . S3.2
B Close-up of costs spent vs to spend in retrospective analysis . . . . . . . . . . . . . . . . . . . S3.3
C Disability adjusted life-years (DALYs) accrued through time for different counterfactual strategies in the prospective analysis. . . . . . . . . . . . . . . . . . . . . . . . . . . . . . . . . . . . S3.4
D Uncertainty in cost-effectiveness for prospective strategies. . . . . . . . . . . . . . . . . . . . . S3.5
E Cost-effectiveness acceptability curves (CEACs) for retrospective strategies . . . . . . . . . . . S3.6
F Expected value of perfect partial information (EVPPI) for retrospective analysis . . . . . . . . S3.8


#### List of Tables


A Costs spent vs total costs for retrospective analysis. . . . . . . . . . . . . . . . . . . . . . . . . S3.2
B Sensitivity analysis if fexinidazole had been available since 2014. . . . . . . . . . . . . . . . . . S3.7

#### S3.1 Supplemental results for the retrospective analysis

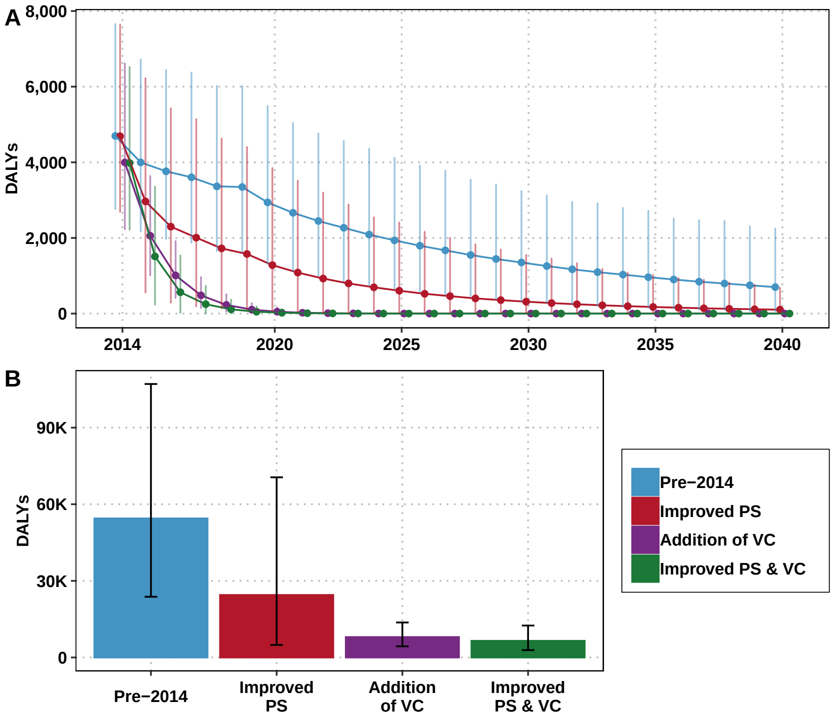


Figure A: Disability adjusted life-years (DALYs) accrued through time for different counterfactual strategies in the retrospective analysis.

| Strategies | Costs 2014-2020 | Costs 2014-2040 | Proportion spent |
|---|---|---|---|
| Pre-2014 | 0.57 (0.43, 0.74) | 1.67 (1.24, 2.20) | 0.35 (0.31, 0.39) |
| Improved PS | 0.84 (0.67, 1.04) | 2.51 (1.67, 3.28) | 0.34 (0.28, 0.45) |
| Addition of VC | 0.89 (0.59, 1.27) | 1.28 (0.86, 1.84) | 0.70 (0.60, 0.72) |
| Improved PS & VC | 1.13 (0.83, 1.51) | 1.78 (1.30, 2.39) | 0.63 (0.54, 0.67) |

Table A: Costs spent vs total costs (in millions USD) for retrospective analysis. Means are given along with 95% prediction intervels (PIs).

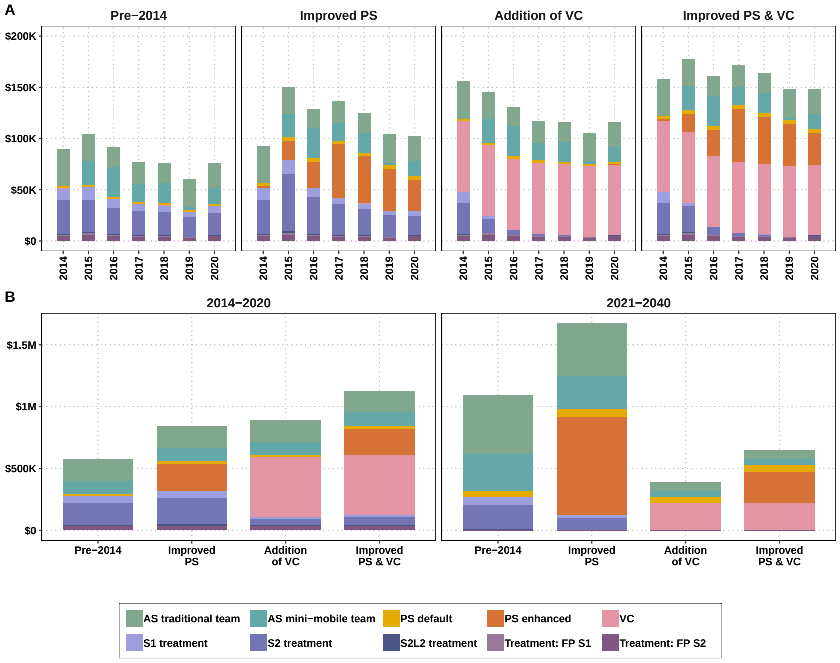


Figure B: Close-up of costs spent vs to spend in retrospective analysis. A) Costs spent by intervention for each strategy in 2014-2020. B) Costs spent vs those to be spent by activity. See table A for total estimates and uncertainty in each period.

#### S3.2 Supplemental results for the prospective analysis

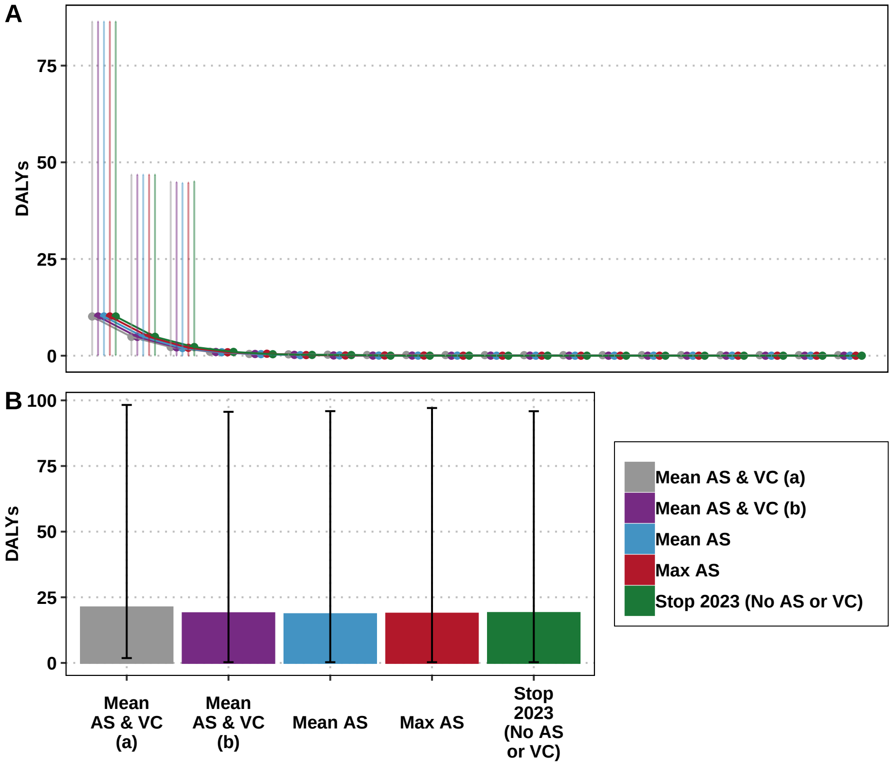


Figure C: Disability adjusted life-years (DALYs) accrued through time for different counterfactual strategies in the prospective analysis.

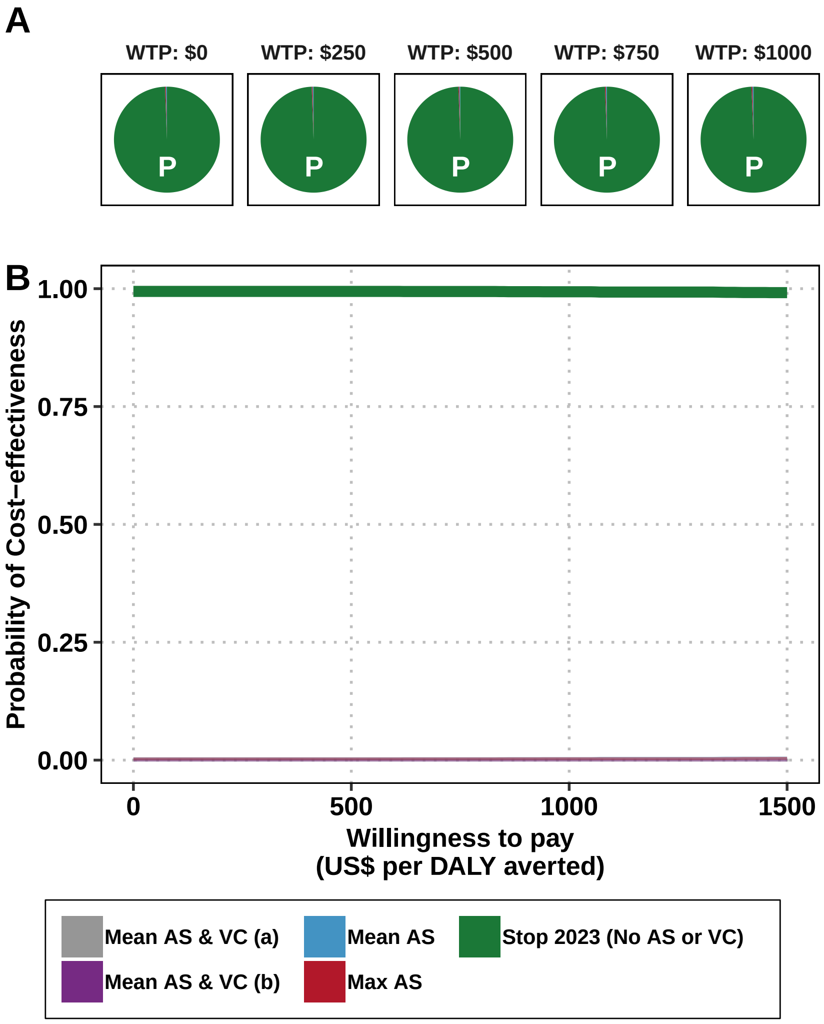


Figure D: Uncertainty in cost-effectiveness for prospective strategies. A) Pie graphs depicting the probability that each strategy is optimal. Strategies with the highest mean net monetary benefit are marked with a "P". B) Cost-effectiveness acceptability curves (CEACs) with the cost-effectiveness acceptability frontier (CEAFs) marked in bold.

#### S3.3 Sensitivity analysis of time horizon and discounting rate

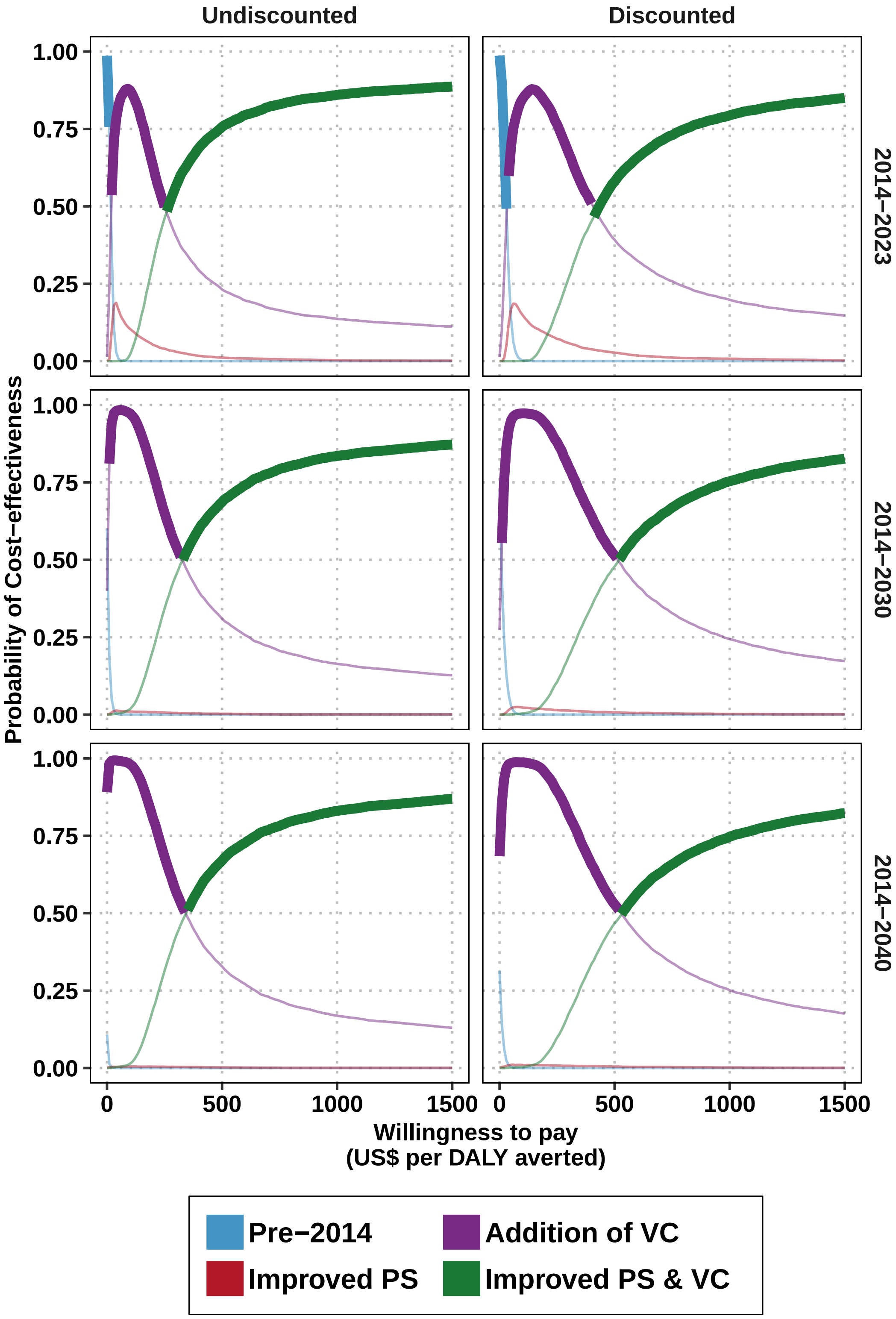


Figure E: Cost-effectiveness acceptability curves (CEACs) for retrospective strategies, sensitivity analysis with alternative horizons with the cost-effectiveness acceptability frontier (CEAFs) marked in bold.

#### S3.4 Sensitivity analysis: results if fexinidazole had been available

| | Pre-2014 | Improved PS | Addition of VC | Improved PS & VC |
|---|---|---|---|---|
| **Health effects** | | | | |
| Reported cases | 1432 (694, 2538) | 1066 (543, 1896) | 185 (93, 315) | 216 (115, 341) |
| Deaths undetected | 1189 (520, 2365) | 533 (102, 1525) | 174 (95, 302) | 142 (62, 272) |
| Cases total | 2620 (1256, 4880) | 1598 (690, 3361) | 359 (215, 556) | 358 (208, 549) |
| Deaths detected[a] | 1 (0, 2) | 1 (0, 2) | 0 (0, 0) | 0 (0, 0) |
| YLD | 862 (458, 1551) | 430 (168, 908) | 111 (63, 187) | 96 (49, 175) |
| YLL | 53,601 (23,117, 105,716) | 24,025 (4,616, 68,057) | 7,865 (4,273, 13,621) | 6,411 (2,763, 12,273) |
| DALYs | 54,463 (23,669, 107,167) | 24,455 (4,843, 68,885) | 7,976 (4,383, 13,724) | 6,507 (2,867, 12,377) |
| **Costs, in thousands US$** | | | | |
| AS costs | 1062 (771, 1425) | 973 (579, 1378) | 405 (293, 548) | 406 (293, 561) |
| PS costs | 67 (38, 107) | 1102 (710, 1585) | 67 (38, 107) | 547 (391, 753) |
| VC costs | 0 (0, 0) | 0 (0, 0) | 695 (312, 1246) | 698 (312, 1268) |
| Treatment costs | 301 (147, 534) | 256 (135, 447) | 61 (35, 97) | 72 (42, 112) |
| Costs total | 1429 (1081, 1852) | 2330 (1542, 3061) | 1228 (809, 1791) | 1723 (1253, 2356) |
| **EoT** | | | | |
| Year of EoT | After 2050 | 2043 (2029, After 2050) | 2015 (2015, 2015) | 2015 (2015, 2015) |
| Prob EoT 2030 | <0.01 | 0.06 | >0.99 | >0.99 |
| **Cost-effectiveness without uncertainty (discounted)[b]** | | | | |
| DALYs averted | 0 | 12,384 | 19,942 | 20,720 |
| Costs averted | 0 | 647,567 | 38,408 | 441,938 |
| ICER | Min Cost | Dominated | 2 | 519 |
| **Cost-effectiveness with uncertainty, conditional on WTP[c].** | | | | |
| WTP: $0 | 0.53(p) | 0 | 0.47 | 0 |
| WTP: $250 | 0 | 0.01 | 0.89(p) | 0.1 |
| WTP: $500 | 0 | 0 | 0.52(p) | 0.47 |
| WTP: $750 | 0 | 0 | 0.33 | 0.66(p) |
| WTP: $1000 | 0 | 0 | 0.25 | 0.75(p) |

[a] Detected deaths are those that occur due to treatment failure or loss-to-follow-up.
[b] Cost-effectiveness results are given for discounted DALYs and costs as per convention
[c] (p) is the preferred strategy; the strategy with the highest mean net monetary benefits

Table B: Sensitivity analysis if fexinidazole had been available since 2014. Summary of effects, costs, elimination of transmission (EoT) by 2030, and cost-effectiveness with and without uncertainty. Highlighted lines are the results that changed after simulating treatment with fexinidazole. Means are given along with 95% prediction intervals (PIs). YLL: years of life lost (to fatal disease), YLD: years of life lost to disability, DALYs: disability-adjusted life-years, PS: passive screening, AS: active screening, VC: vector control, ICER: incremental cost-effectiveness ratio, WTP: willingness to pay (USD per DALY averted), EoT: elimination of transmission.

#### S3.5 Parameter-specific uncertainty analysis (retrospective analysis)

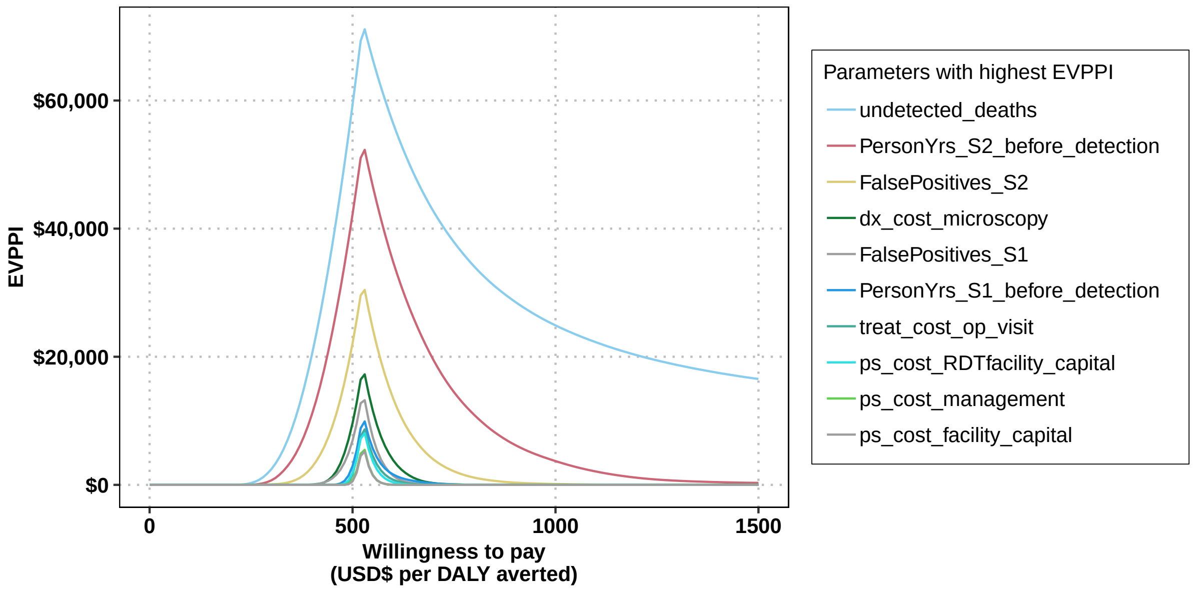


Figure F: Expected value of perfect partial information (EVPPI) for retrospective analysis.
