## Supplementary material for "Health economic evaluation of strategies to eliminate *gambiense* human African trypanosomiasis in the Mandoul disease focus of Chad": S4 Text. Parameter glossary.

### S4. Parameter Glossary

**Contents**


S4.1 Organization of parameters . . . S4.3
S4.2 Principles for parameterization . . . S4.3
S4.3 Summary of health outcome parameters . . . S4.4
S4.4 Summary of cost parameters . . . S4.8
S4.5 Notes on Screening Parameters . . . S4.11
S4.5.1 pop . . . S4.11
S4.5.2 as_num_screened_trad . . . S4.11
S4.5.3 as_num_screened_moto . . . S4.11
S4.5.4 as_prop_screened_moto . . . S4.12
S4.5.5 as_ltfu_moto . . . S4.12
S4.5.6 as_factor_num_screened_enhanced . . . S4.12
S4.5.7 dx_sens_rdt . . . S4.13
S4.5.8 dx_spec_catt . . . S4.13
S4.5.9 dx_spec_rdt . . . S4.13
S4.5.10 ps_coverage . . . S4.13
S4.5.11 ps_ltfu_enhanced . . . S4.14
S4.5.12 ps_num_facilities . . . S4.14
S4.5.13 ps_num_facilities_enhanced . . . S4.14
S4.5.14 dx_wastage_catt_as . . . S4.15
S4.5.15 dx_wastage_catt_ps . . . S4.15
S4.5.16 dx_wastage_rdt . . . S4.15
S4.6 Notes on Treatment Parameters . . . S4.16
S4.6.1 prob_late_stage2 . . . S4.16
S4.6.2 treat_duration_fexi . . . S4.16
S4.6.3 treat_duration_nect . . . S4.17
S4.6.4 treat_duration_sae . . . S4.17
S4.6.5 treat_prob_failure_fexi . . . S4.17
S4.6.6 treat_prob_failure_fexi_late_s2 . . . S4.18
S4.6.7 treat_prob_failure_nect_s2 . . . S4.18
S4.6.8 treat_prob_failure_pent_s1 . . . S4.19
S4.6.9 treat_prob_sae_fexi . . . S4.19
S4.6.10 treat_prob_sae_nect_s2 . . . S4.20
S4.6.11 treat_prob_sae_pent_s1 . . . S4.20
S4.6.12 treat_prob_under35kg . . . S4.21
S4.6.13 treat_prob_under6yo . . . S4.23
S4.7 Notes on Life-years lost (DALY) Parameters . . . S4.24
S4.7.1 disability_weighting_s1 . . . S4.24
S4.7.2 disability_weighting_s2 . . . S4.24
S4.7.3 disability_weighting_sae . . . S4.25
S4.7.4 age_of_death . . . S4.25

S4.8 Notes on Screening and Vector Control Cost Parameters . . . S4.25
S4.8.1 as_cost_moto_capital . . . S4.25
S4.8.2 as_cost_moto_followup . . . S4.27
S4.8.3 as_cost_moto_management . . . S4.27
S4.8.4 as_cost_team_capital . . . S4.27
S4.8.5 as_cost_team_management . . . S4.28
S4.8.6 dx_cost_catt . . . S4.28
S4.8.7 dx_cost_lumbar_exam . . . S4.29
S4.8.8 dx_cost_mAECT . . . S4.29
S4.8.9 dx_cost_LAMP . . . S4.29
S4.8.10 dx_cost_microscopy . . . S4.29
S4.8.11 dx_cost_rdt . . . S4.30
S4.8.12 ps_cost_facility_capital . . . S4.30
S4.8.13 ps_cost_management . . . S4.31
S4.8.14 ps_cost_RDTfacility_capital . . . S4.31
S4.8.15 vc_cost . . . S4.31
S4.9 Notes on Treatment Cost Parameters . . . S4.32
S4.9.1 rx_cost_fexinidazole . . . S4.32
S4.9.2 rx_cost_nect . . . S4.32
S4.9.3 rx_cost_pentamidine . . . S4.32
S4.9.4 rx_delivery_markup . . . S4.32
S4.9.5 treat_cost_ip_day . . . S4.33
S4.9.6 treat_cost_op_visit . . . S4.33

#### List of Tables

A Screening parameters . . . S4.5
B Treatment parameters . . . S4.6
C Life-years lost (DALY) parameters . . . S4.7
D Screening and VC cost parameters . . . S4.9
E Treatment cost parameters . . . S4.10

#### S4.1 Organization of parameters

The code makes use of five r lists: `surv_effect`, `surv_cost`, `treat_effect`, and `treat_cost`. Within those lists sit a set of variables described in the tables later in this supplement.

- `surv_effect`: these are the parameters related to the consumption (units) and effectiveness of screening and vector control activities.
- `surv_cost`: these are the costs of active screening and passive surveillance. The units are either general demographic units (cases or population) or parameters found in surv_effect.
- `treat_effect`: these are the parameters related to the consumption (units) and effectiveness of treatment.
- `treat_cost`: these are the costs of treatment. The units are either general demographic units (cases or population) or parameters found in treat_effect.

Additionally, a list named `epi_output` holds the output from the dynamic model: stage 1 and stage 2 cases detected by passive and active surveillance.

#### S4.2 Principles for parameterization

These are the rules we follow to parameterize models. The rationale behind some of the items in the tables are easier to understand with these rules in mind.

- **Transferability of costs across time** Costs have been updated to 2020 USD values. All costs are converted to local currency units in the year of the study, inflated to 2020 values using the consumer price index (CPI) of the country, and then converted to USD using the exchange rate in 2020. It should be noted that the 2003 WHO Guide to Cost-effectiveness recommends that the GDP inflator be used (see 3.2.6 Transferability of costs across time, page 43) but we found that the data on this measure (from the World Bank) were sometimes sparse so we relied on the consumer price index instead [1].

- **Transferability of costs across settings** To 'borrow' data from other countries, we follow the 2003 WHO Guide to Cost-effectiveness recommendations in section 3.2.7 Transferability of costs across settings) [1]. For non-traded items (nurse and doctor time) we convert USD or LCU prices into PPP (international dollars) values in the year of the cost study and then turn the value in international dollars to local currency (still in the year of the study) of the country where a cost estimate is needed. Then, we use the CPI to inflate costs to 2018 levels and then use the exchange rate with USD to get 2020 USD values.

- **Combining multiple sources of information** Values from different publications are combined using meta-analytic methods, as described, in sections S4.5-S4.9.

- **Choice of probability distributions** Costs and ratios were modeled via gamma distributions and proportions or probability were modeled with beta distributions. These distributions were parameterized using the method of moments (see Briggs 2006 [2], Chapter 4).

- **Missing information on uncertainty: Gamma distributions.**

1. Option A: Whenever uncertainty was missing for a cost or ratio, we assigned the parameter a gamma distribution that would yield credible intervals between half and double the estimate available in the literature.

2. Option B: If at least 2 studies listed a cost, then we take the range of the costs to parameterize a gamma distribution for which the the 95 percent confidence interval (the 2.5th and 97.5th percentile) matches the range of estimates from the literature. For these occasions, we use a method to parameterize gamma distributions using quantiles rather than using the mean and standard error of a sample (method of moments) [3].

- **Missing information on uncertainty: Beta distributions.**

1. Option A: Usually modeled assuming that 100 trials were observed with the **proportion_estimate x 100** as the alpha parameter and **(1-proportion_estimate) x 100** as beta parameter.

2. Option B: If at least 2 studies listed a probability or a proportion, then we take the range of the costs to parameterize a beta distribution by assuming the range matches the 95 percent confidence interval (the 2.5th and 97.5th percentile). We use a method to parameterize Beta distributions using quantiles rather than the mean and standard error of a sample (method of moments) [4].

#### S4.3 Summary of health outcome parameters

Below are all the parameters that model health outcomes, as well as a summary of their characteristics. An extended discussion of our choices is featured in the sections annotated in the table.

| Variable description | Variable name | Statistical Distribution | Descriptive Summary | Notes |
|---|---|---|---|---|
| Population | pop | Fixed value | 41000 | See section S4.5.1 |
| Active surveillance, population seen by traditional teams_x000D_ | as_num_screened_trad | Fixed value | Varies by year | See section S4.5.2 |
| Active surveillance, population seen by motorcycle teams | as_num_screened_moto | Fixed value | Varies by year | See section S4.5.3 |
| Proportion of people screened actively who are screened by motorbike teams | as_prop_screened_moto | Fixed value | Varies by year | See section S4.5.4 |
| RDT loss-to-follow-up (active screening) | as_ltfu_moto | Fixed value | Varies by year | See section S4.5.5 |
| Active surveillance, population seen max vs mean AS | as_factor_num_screened_enhanced | Fixed value | 0.3891 | See section S4.5.6 |
| RDT algorithm: diagnostic sensitivity | dx_sens_rdt | Beta(230, 1) | 1.00 (0.98, 1.00) | See section S4.5.7 |
| CATT Algorithm: Diagnostic specificity | dx_spec_catt | Beta(2, 31) | 0.06 (<0.01, 0.16) | See section S4.5.8 |
| RDT Algorithm: diagnostic specificity | dx_spec_rdt | Beta(3886, 24) | 0.99 (0.99, 1.00) | See section S4.5.9 |
| Passive surveillance: coverage of the population per facility as a proportion of the population | ps_coverage | Fixed value | Varies by year | See section S4.5.10 |
| Loss-to-follow-up for RDT-positives in clinics without lab for confirmation | ps_ltfu_enhanced | Fixed value | Varies by year | See section S4.5.11 |
| Passive surveillance: number of facilities | ps_num_facilities | Fixed value | 1 | See section S4.5.12 |
| Passive surveillance: Number of facilties with enhanced screening | ps_num_facilities_enhanced | Fixed value | Varies by year | See section S4.5.13 |
| CATT algorithm: wastage in the context of active screening | dx_wastage_catt_as | Beta(8, 92) | 0.08 (0.04, 0.14) | See section S4.5.14 |
| CATT algorithm: wastage in the context of active screening | dx_wastage_catt_ps | Beta(25, 75) | 0.25 (0.17, 0.34) | See section S4.5.15 |
| Diagnostic, RDT wastage | dx_wastage_rdt | Beta(1, 99) | 0.01 (<0.01, 0.04) | See section S4.5.16 |

Table A: Screening parameters

| Variable description | Variable name | Statistical Distribution | Descriptive Summary | Notes |
|---|---|---|---|---|
| Proportion cases of severe stage 2 (S2) as a proportion of cases in stage 2 (S2) | prob_late_stage2 | Beta(76.93, 44.87) | 0.63 (0.55, 0.71) | See section S4.6.1 |
| Duration of a course of treatment with fexinidazole | treat_duration_fexi | Fixed value | 10 | See section S4.6.2 |
| Length of hospital stay: NECT treatment | treat_duration_nect | Fixed value | 10 | See section S4.6.3 |
| Duration of hospital stay for SAE | treat_duration_sae | Gamma(1.22, 2.38) | 2.88 (0.14, 9.65) | See section S4.6.4 |
| Probability of relapse (treatment failure): fexinidazole for S1/Early S2 disease | treat_prob_failure_fexi | Beta(9.49, 496.54) | 0.02 (<0.01, 0.03) | See section S4.6.5 |
| Probability of relapse (treatment failure) - fexinidazole for late S2 disease | treat_prob_failure_fexi_late_s2 | Beta(43.56, 300.11) | 0.13 (0.09, 0.17) | See section S4.6.6 |
| Probability of relapse (treatment failure): NECT for S2 disease | treat_prob_failure_nect_s2 | Beta(15.87, 378.55) | 0.04 (0.02, 0.06) | See section S4.6.7 |
| Probability of relapse (treatment failure): pentamidine for S1 disease | treat_prob_failure_pent_s1 | Beta(50.3, 665.48) | 0.07 (0.05, 0.09) | See section S4.6.8 |
| Serious adverse events (SAE): fexinidazole treatment | treat_prob_sae_fexi | Beta(3, 261) | 0.01 (<0.01, 0.03) | See section S4.6.9 |
| Serious adverse events (SAE): NECT treatment | treat_prob_sae_nect_s2 | Beta(40.88, 367.8) | 0.10 (0.07, 0.13) | See section S4.6.10 |
| Serious adverse events (SAE): pentamidine treatment | treat_prob_sae_pent_s1 | Beta(1.43, 551.42) | 0.003 (<0.001, 0.008) | See section S4.6.11 |
| Percent cases under 35 kg as a proportion of all cases over the age of 6 | treat_prob_under35kg | Beta(8.3, 359.6) | 0.02 (<0.01, 0.04) | See section S4.6.12 |
| Percent cases under 6 as a proportion of all cases | treat_prob_under6yo | Beta(152.53, 2427.9) | 0.06 (0.05, 0.07) | See section S4.6.13 |

Table B: Treatment parameters

| Variable description | Variable name | Statistical Distribution | Descriptive Summary | Notes |
|---|---|---|---|---|
| Disability weights: stage 1 (S1) disease | disability_weighting_s1 | Beta(22.96, 147.21) | 0.14 (0.09, 0.19) | See section S4.7.1 |
| Disability weights: stage 2 (S2) disease | disability_weighting_s2 | Beta(18.37, 15.63) | 0.54 (0.37, 0.70) | See section S4.7.2 |
| Disability weights: serious adverse events (SAE) | disability_weighting_sae | Uniform(0.04, 0.11) | 0.08 (0.04, 0.11) | See section S4.7.3 |
| Age of death from infection (for years of life lost per fatal case) | age_of_death | Gamma(148, 0.18) | 26.65 (22.56, 31.07) | See section S4.7.4 |

Table C: Life-years lost (DALY) parameters

#### S4.4 Summary of cost parameters

Below are all the cost parameters and a summary of their characteristics in three tables for surveillance, treatment, and vector control costs. An extended discussion of our choices is featured in the sections annotated in the tables.

| Variable description | Variable name | Statistical Distribution | Descriptive Summary | Notes |
|---|---|---|---|---|
| Active screening: capital costs of a motorcycle team | as_cost_moto_capital | Gamma(8.47, 1000.81) | 8,489 (3,742, 14,809) | See section S4.8.1 |
| Active screening: motorcycle-based follow-up of patients | as_cost_moto_followup | Gamma(70.05, 54.93) | 3,843 (2,992, 4,782) | See section S4.8.2 |
| Active screening: fixed management costs of a motorcycle team | as_cost_moto_management | Gamma(70.05, 91.56) | 6,411 (5,017, 7,965) | See section S4.8.3 |
| Active screening: capital costs of a traditional team | as_cost_team_capital | Gamma(200000, 0.02) | 4,000 (3,983, 4,017) | See section S4.8.4 |
| Active screening: fixed management costs of a traditional team | as_cost_team_management | Gamma(8.47, 2167) | 18,361 (8,308, 32,750) | See section S4.8.5 |
| CATT algorithm: cost per test used | dx_cost_catt | Gamma(22.87, 0.02) | 0.46 (0.29, 0.66) | See section S4.8.6 |
| Lumbar puncture and laboratory exam: cost per person that needs to be staged | dx_cost_lumbar_exam | Gamma(3.26, 2.87) | 9.38 (2.11, 21.85) | See section S4.8.7 |
| mAECT diagnosis: cost | dx_cost_mAECT | Gamma(8.47, 0.23) | 1.95 (0.87, 3.51) | See section S4.8.8 |
| LAMP diagnosis: cost | dx_cost_LAMP | Gamma(8.47, 0.62) | 5.27 (2.32, 9.41) | See section S4.8.9 |
| Microscopy (Bloodsample, LNA, mAECT): cost per person that needs to be confirmed | dx_cost_microscopy | Gamma(8.47, 1.27) | 10.70 (4.75, 18.98) | See section S4.8.10 |
| RDT algorithm: costs per test used | dx_cost_rdt | Gamma(8.47, 0.19) | 1.60 (0.72, 2.84) | See section S4.8.11 |
| Passive surveillance: capital costs of a facility | ps_cost_facility_capital | Gamma(8.47, 216.24) | 1,826 (798, 3,241) | See section S4.8.12 |
| Passive surveillance: fixed recurrent management costs | ps_cost_management | Gamma(32.47, 12.18) | 396.09 (271.96, 542.85) | See section S4.8.13 |
| Capital costs of RDT facilities (training only) | ps_cost_RDTfacility_capital | Gamma(8.47, 31.97) | 270.21 (119.69, 477.03) | See section S4.8.14 |
| The total cost of vector control | vc_cost | Gamma(8.47, 8132) | 68,917 (30,975, 121,361) | See section S4.8.15 |

Table D: Screening and VC cost parameters

| Variable description | Variable name | Statistical Distribution | Descriptive Summary | Notes |
|---|---|---|---|---|
| Course of fexinidazole: cost | rx_cost_fexinidazole | Gamma(100, 0.5) | 50.00 (40.76, 60.34) | See section S4.9.1 |
| Course of nifurtimox eflornithine combination therapy (NECT): cost | rx_cost_nect | Gamma(100, 3.6) | 360.47 (291.82, 433.21) | See section S4.9.2 |
| Course of pentamidine: cost | rx_cost_pentamidine | Gamma(100, 0.54) | 54.11 (44.12, 65.18) | See section S4.9.3 |
| Drug delivery mark-up | rx_delivery_markup | Uniform(0.15, 0.25) | 0.20 (0.15, 0.25) | See section S4.9.4 |
| Hospital stay: cost per day | treat_cost_ip_day | Gamma(5.45, 1.76) | 9.60 (3.40, 19.44) | See section S4.9.5 |
| Outpatient consultation: cost | treat_cost_op_visit | Gamma(2.48, 0.79) | 1.96 (0.31, 5.05) | See section S4.9.6 |

Table E: Treatment cost parameters

### S4.5 Notes on Screening Parameters

#### S4.5.1 pop

- Name in the code: pop
- Description of variable: Population
- Source: [5, 6]]
- Country of estimate: NA
- Statistical distribution and parameters: Fixed value
- Summary statistics (mean and 95% CI or fixed value): 41000

**Notes**

The population of Mandoul focus was estimated by a census in 2013 [5]. In that year, the focus was dotted by 114 settlements of the Sara ethnic group: 1029 people lived in 22 encampments of fewer than 100 people, 17,626 lived in hamlets of between 100-500 people, and 20,016 people lived in 27 villages of more than 500 people [5].

#### S4.5.2 as_num_screened_trad

- Name in the code: as_num_screened_trad
- Description of variable: Active surveillance, population seen by traditional teams_x000D_
- Source: HAT Atlas data, see SI Text 1, Table A and B
- Country of estimate: NA
- Statistical distribution and parameters: Fixed value
- Summary statistics (mean and 95% CI or fixed value): Varies by year

**Notes**

The capacity of Chad teams has been reported at up to 1500 per day [7]. Although villages are mostly of fewer than 500 people, teams cover multiple teams per day. However, most of the data has shown that the practical capacity lies closed to 700-1000 cases.

See SI Text 1, Table A for the capacity from year to year.

| Year | Estimate |
|---|---|
| 2014 | 22146 |
| 2015 | 17932 |
| 2016 | 11302 |
| 2017 | 9284 |
| 2018 | 10646 |
| 2019 | 11411 |
| 2020 | 18445 |
| 2021 | 13543 |

#### S4.5.3 as_num_screened_moto

- Name in the code: as_num_screened_moto
- Description of variable: Active surveillance, population seen by motorcycle teams
- Source: HAT Atlas data, see SI Text 1, Table A and B
- Country of estimate: TCD_mandoul
- Statistical distribution and parameters: Fixed value
- Summary statistics (mean and 95% CI or fixed value): Varies by year

**Notes**

For the capacity of motorcycle teams, see SI Text 1, Table B. The mean capacity was 79.51 and the standard error was 7.82. Therefore, we parameterised the model with Normal(79.51, 7.82). According to information from the teams, they visit about one village per day and teams work around the year. The values of the parameter therefore have a distribution of 79.51(62.88, 94.14)

The lower number of capacity by motorcycle teams compared to traditional teams is due to the fact that the settlements and hamlets that motorcycle teams go to are less populated, and therefore teams spend more time for travel and set up of activities.

| Year | Estimate |
|---|---|
| 2014 | 0 |
| 2015 | 9333 |
| 2016 | 10705 |
| 2017 | 6860 |
| 2018 | 7437 |
| 2019 | 1229 |
| 2020 | 1183 |
| 2021 | 6085 |

##### S4.5.4 as_prop_screened_moto

- Name in the code: `as_prop_screened_moto`
- Description of variable: Proportion of people screened actively who are screened by motorbike teams
- Source: HAT Atlas data, see SI Text 1, Table A and B
- Country of estimate: TCD
- Statistical distribution and parameters: Fixed value
- Summary statistics (mean and 95% CI or fixed value): Varies by year

**Notes**

The split in the number of people screened between traditional and motorcycle teams was available from program data shown in SI Text 1, Tables A-B.

| Year | Estimate |
|---|---|
| 2014 | 0.00 |
| 2015 | 0.34 |
| 2016 | 0.49 |
| 2017 | 0.38 |
| 2018 | 0.41 |
| 2019 | 0.09 |
| 2020 | 0.31 |
| 2021 | 0.31 |

##### S4.5.5 as_ltfu_moto

- Name in the code: `as_ltfu_moto`
- Description of variable: RDT loss-to-follow-up (active screening)
- Source: HAT Atlas data, see SI Text 1, Table A and B
- Country of estimate: NA
- Statistical distribution and parameters: Fixed value
- Summary statistics (mean and 95% CI or fixed value): Varies by year

**Notes**

Because the data showed significant heterogeneity according to the tau-squared test for heterogeneity, we have chosen to use the random-effects estimate, to which we assigned a distribution of Beta(4.16, 14.03).

##### S4.5.6 as_factor_num_screened_enhanced

- Name in the code: `as_factor_num_screened_enhanced`
- Description of variable: Active surveillance, population seen max vs mean AS
- Source: HAT Atlas data, see SI Text 1, Table A and B

- Country of estimate: NA
- Statistical distribution and parameters: Fixed value
- Summary statistics (mean and 95% CI or fixed value): 0.3891

**Notes**

For the years 2021 and after, we simulate active screening of 19,628 people for *Mean AS* and 22,146 for Max AS. Therefore, the *Max AS* has 1.12 times as many screenings as *Mean AS*.

##### S4.5.7 dx_sens_rdt

- Name in the code: `dx_sens_rdt`
- Description of variable: RDT algorithm: diagnostic sensitivity
- Source: [8]
- Country of estimate: NA
- Statistical distribution and parameters: Beta(230, 1)
- Summary statistics (mean and 95% CI or fixed value): 1.00 (0.98, 1.00)

**Notes**

Based on a study in Guinea and Cote d'Ivoire [8], there was 1 sample from a gHAT patient that tested negative out of 231. Therefore, the parameter distribution for specifity is Beta(230, 1).

##### S4.5.8 dx_spec_catt

- Name in the code: `dx_spec_catt`
- Description of variable: CATT Algorithm: Diagnostic specificity
- Source: [9]
- Country of estimate: NA
- Statistical distribution and parameters: Beta(2, 31)
- Summary statistics (mean and 95% CI or fixed value): 0.06 (<0.01, 0.16)

**Notes**

Based on a systematic review by Mitashi and colleagues [9], who reported ranges of 83.5-99.3%. The beta distribution that corresponds with that range as 95% confidence intervals is Beta(31, 2).

##### S4.5.9 dx_spec_rdt

- Name in the code: `dx_spec_rdt`
- Description of variable: RDT Algorithm: diagnostic specificity
- Source: [8]
- Country of estimate: TCD
- Statistical distribution and parameters: Beta(3886, 24)
- Summary statistics (mean and 95% CI or fixed value): 0.99 (0.99, 1.00)

**Notes**

Specificity of RDT has been documented by a study in Guinea and Cote d'Ivoire [8], where there were 31 samples from non-HAT patients that tested positive out of 257. However, we consider the completement of the positivity rate (1-positivity rate) in the passive screening system, so the distribution is Beta(3886, 24) which is summarized by: 0.9939 (0.9912, 0.9961).

##### S4.5.10 ps_coverage

- Name in the code: `ps_coverage`
- Description of variable: Passive surveillance: coverage of the population per facility as a proportion of the population
- Source: HAT Atlas data, see SI Text 1, Table
- Country of estimate: NA
- Statistical distribution and parameters: Fixed value
- Summary statistics (mean and 95% CI or fixed value): Varies by year

**Notes**

For the number of patients screened in passive surveillance, see table C for the years 2014-2020. For those years, the parameter is a fixed value.

For the years 2021 and later we considered the mean number of RDTs administered from 2014-2020, which was 92.72 and had a standard error of 12.20. Therefore, we parameterised the model with Normal(92.72, 12.20). The distribution of the parameter is therefore 92.72(68.32, 117.12).

| Year | Estimate |
|---|---|
| 2014 | 100 |
| 2015 | 119 |
| 2016 | 128 |
| 2017 | 101 |
| 2018 | 55 |
| 2019 | 82 |
| 2020 | 39 |
| 2021 | 93 (69, 117) |

##### S4.5.11 ps_ltfu_enhanced

- Name in the code: `ps_ltfu_enhanced`
- Description of variable: Loss-to-follow-up for RDT-positives in clinics without lab for confirmation
- Source: HAT Atlas data, see SI Text 1, Table
- Country of estimate: TCD
- Statistical distribution and parameters: Fixed value
- Summary statistics (mean and 95% CI or fixed value): Varies by year

**Notes**

Because the data showed significant heterogeneity according to the tau-squared test for heterogeneity, we have chosen to use the random-effects estimate, to which we assigned a distribution of Beta(4.16, 14.03).

##### S4.5.12 ps_num_facilities

- Name in the code: `ps_num_facilities`
- Description of variable: Passive surveillance: number of facilities
- Source: HAT Atlas data, see SI Text 1, Table
- Country of estimate: NA
- Statistical distribution and parameters: Fixed value
- Summary statistics (mean and 95% CI or fixed value): 1

**Notes**

Before the work by FIND, there was only one facility that could screen, diagnose, or treat any gHAT cases: the Catholic Mission Hospital in Bodo. The hospital began screening cases in 1993, and has seized operations related to HAT in 2019, when the specialist for diagnosis and treatment, Sister Cecile was no longer available.

##### S4.5.13 ps_num_facilities_enhanced

- Name in the code: `ps_num_facilities_enhanced`
- Description of variable: Passive surveillance: Number of facilties with enhanced screening
- Source: HAT Atlas data, see SI Text 1, Table
- Country of estimate: NA
- Statistical distribution and parameters: Fixed value
- Summary statistics (mean and 95% CI or fixed value): Varies by year

**Notes**

See SI Text 1, Table A for the capacity from year to year. One new facility was prepared for diagnosis and treatment of gHAT cases: Bodo district hospital. The other facilities were only stocked for screening with RDT tests. In 2019, a set of clinics was closed as these were in locations with no cases detected and very few individuals screened.

| Year | Estimate |
|---|---|
| 2014 | 0 |
| 2015 | 8 |
| 2016 | 8 |
| 2017 | 29 |
| 2018 | 29 |
| 2019 | 21 |
| 2020 | 21 |
| 2021 | 21 |

##### S4.5.14 dx_wastage_catt_as

- Name in the code: `dx_wastage_catt_as`
- Description of variable: CATT algorithm: wastage in the context of active screening
- Source: [10]
- Country of estimate: NA
- Statistical distribution and parameters: Beta(8, 92)
- Summary statistics (mean and 95% CI or fixed value): 0.08 (0.04, 0.14)

**Notes**

CATT tests come in packs of 50, and the list cost is assumed to consider that a pack is used on 50 patients. Once a pack is opened, one test is used as a positive control and one test is used as a negative control, so wastage is at least 4 percent. Shelf life of the test is one week in refrigeration and wastage in active screening activities is relatively low; generally, wastage of CATT tests in the context of active screening occurs at the end of the day when there are tests remaining in an open pack. To be conservative, we doubled the 4-percent lower bound for wastage and assigned the parameter a distribution of Beta(8, 92).

##### S4.5.15 dx_wastage_catt_ps

- Name in the code: `dx_wastage_catt_ps`
- Description of variable: CATT algorithm: wastage in the context of active screening
- Source: [10]
- Country of estimate: COD
- Statistical distribution and parameters: Beta(25, 75)
- Summary statistics (mean and 95% CI or fixed value): 0.25 (0.17, 0.34)

**Notes**

CATT tests come in packs of 50, and the list cost is assumed to consider that a pack is used on 50 patients. Once a pack is opened, one test is used as a positive control, one test is used as a negative control, so wastage is at least of 4 percent. Shelf life of the test is one week in refrigeration, but wastage in passive surveillance is relatively high because the frequency of patients per week might be well under 50 patients. Our colleague working in DRC suggested that a 25-percent wastage factor would be appropriate (personal communication, Rian Snijders, ITM).

##### S4.5.16 dx_wastage_rdt

- Name in the code: `dx_wastage_rdt`
- Description of variable: Diagnostic, RDT wastage
- Source: [11]
- Country of estimate: NA
- Statistical distribution and parameters: Beta(1, 99)
- Summary statistics (mean and 95% CI or fixed value): 0.01 (<0.01, 0.04)

**Notes**

We followed the same assumption as Snijders and colleagues that less than 1 percent of RDT tests would not be used [11]. Because there was no sense of uncertainty in this parameter, we assumed a Beta (1,99) distribution.

#### S4.6 Notes on Treatment Parameters

##### S4.6.1 prob_late_stage2

- Name in the code: `prob_late_stage2`
- Description of variable: Proportion cases of severe stage 2 (S2) as a proportion of cases in stage 2 (S2)
- Source: [12–19]
- Country of estimate: NA
- Statistical distribution and parameters: Beta(76.93, 44.87)
- Summary statistics (mean and 95% CI or fixed value): 0.63 (0.55, 0.71)

**Notes**

The definition of severe stage 2 gHAT disease by the WHO is when there are more than 100 white blood cells (WBC, leukocytes) per micro-liter in the cerebro-spinal fluid. We have searched the clinical trials for the proportion of stage 2 patients that have high concentrations of leukocytes upon admission to treatment.

| **Study** | **Arm** | **Events** | **Total** | **Est.** | **95% CI** |
|---|---|---|---|---|---|
| *Priotto 2006* | Melarsoprol and Nifurtimox | 9 | 18 | 0.50 | (0.26−0.74) |
| *Priotto 2006* | Melarsoprol and Eflornithine | 12 | 19 | 0.63 | (0.38−0.84) |
| *Priotto 2006* | NECT | 6 | 17 | 0.35 | (0.14−0.62) |
| *Eperon 2006* | Melarsoprol | 407 | 1108 | 0.37 | (0.34−0.40) |
| *Priotto 2007* | NECT | 31 | 52 | 0.60 | (0.45−0.73) |
| *Priotto 2007* | Eflornithine | 39 | 51 | 0.76 | (0.63−0.87) |
| *Checchi 2007* | NECT | 18 | 31 | 0.58 | (0.39−0.75) |
| *Priotto 2009* | NECT | 107 | 143 | 0.75 | (0.67−0.82) |
| *Priotto 2009* | Eflornithine | 115 | 143 | 0.80 | (0.73−0.87) |
| *Ngoyi 2010* | Pentamidine and Melarsoprol | 209 | 272 | 0.77 | (0.71−0.82) |
| *Priotto 2012* | Single arm | 1373 | 2190 | 0.63 | (0.61−0.65) |
| *Schmid 2012* | Single arm | 397 | 629 | 0.63 | (0.59−0.67) |
| **Common effect model** | | | **4673** | **0.58** | **(0.57−0.60)** |
| **Random effects model** | | | | **0.63** | **(0.55−0.71)** |

Heterogeneity: $I^2$ = 96%, $\tau^2$ = 0.3, $p$ < 0.01

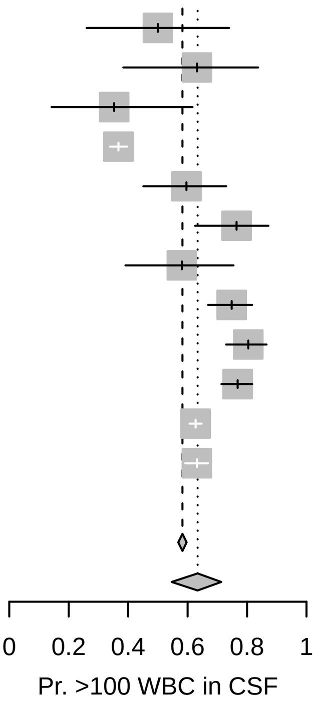


Because the data showed significant heterogeneity according to the tau-squared test for heterogeneity, we have chosen to use the random-effects combined estimate: 0.634 (0.546, 0.713), represented by the probability distribution Beta(76.93, 44.87).

##### S4.6.2 treat_duration_fexi

- Name in the code: `treat_duration_fexi`
- Description of variable: Duration of a course of treatment with fexinidazole
- Source: [20]
- Country of estimate: NA
- Statistical distribution and parameters: Fixed value
- Summary statistics (mean and 95% CI or fixed value): 10

**Notes**

For the only trial that is published [20], patients were released on days 13-18 after the initiation of treatment, although the treatment only took 10 days, so we have assumed that in routine care that the average patient will be in inpatient care for 10 days.

##### S4.6.3 treat_duration_nect

- Name in the code: `treat_duration_nect`
- Description of variable: Length of hospital stay: NECT treatment
- Source: [20]
- Country of estimate: NA
- Statistical distribution and parameters: Fixed value
- Summary statistics (mean and 95% CI or fixed value): 10

**Notes**

For NECT treatment, patients must stay in inpatient care for a minimum of 7 days for the eflornithine infusions, and whether they stay for a total of 10 days for nifurtimox administration is unclear. For the most recent clinical trial [20], NECT patients were released on days 13-18 after admission, but we have assumed that for the most recent trial the average patient can be released from care after 10 days in the hospital.

##### S4.6.4 treat_duration_sae

- Name in the code: `treat_duration_sae`
- Description of variable: Duration of hospital stay for SAE
- Source: [21]
- Country of estimate: NA
- Statistical distribution and parameters: Gamma(1.22, 2.38)
- Summary statistics (mean and 95% CI or fixed value): 2.88 (0.14, 9.65)

**Notes**

Our only source of information for the duration of severe adverse events (SAEs) is Alirol 2013 [21], which lists the most common adverse events and the median duration of these events. Most envents last a median of 1-2 days (with interquartile ranges reaching up to 4 days).

For simplicity, we have fit a gamma distribution with interquartile range of 1-4 days. Our distribution is therefore Gamma(1.22, 2.38) with a mean and 95% confidence interval of 2.93 (0.13, 10.06), which provides a sufficiently large range of values in light of the scarce information we have.

##### S4.6.5 treat_prob_failure_fexi

- Name in the code: `treat_prob_failure_fexi`
- Description of variable: Probability of relapse (treatment failure): fexinidazole for S1/Early S2 disease
- Source: [19]
- Country of estimate: NA
- Statistical distribution and parameters: Beta(9.49, 496.54)
- Summary statistics (mean and 95% CI or fixed value): 0.02 (<0.01, 0.03)

**Notes**

Mesu and colleagues [20] have published the only study on fexinidazole treatment effectiveness on late-stage 2 cases. Moreover, the accompanying meta-analysis for the WHO treatment guidelines released in 2019 show the outcomes an additional extension study on stage 1, both early and late-stage 2 disease as well we for the data from Mesu et al stratified by the concentration of WBC in the CSF [19].

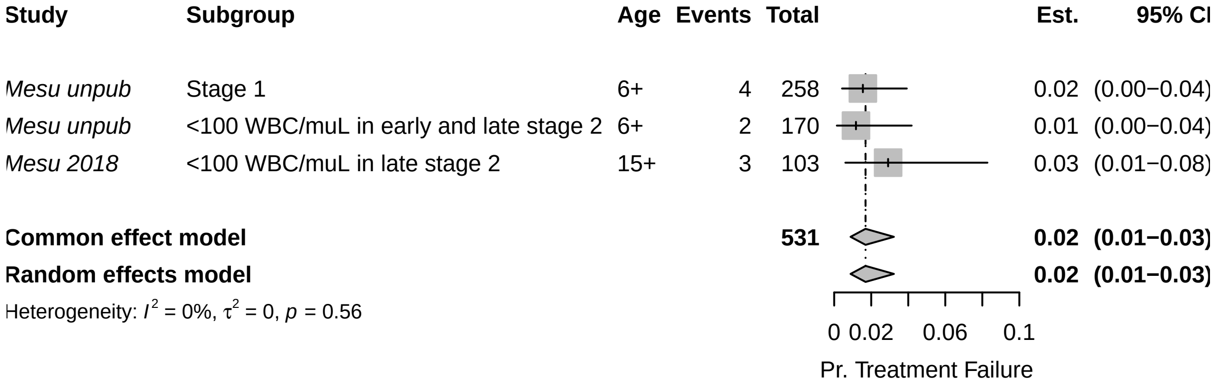

Because the data showed non-significant heterogeneity according to the tau-squared test for heterogeneity, we have chosen to use the fixed-effects combined estimate: 0.017 (0.009, 0.032), for which we assigned a distribution of Beta(9.49, 496.54).

##### S4.6.6 treat_prob_failure_fexi_late_s2

- Name in the code: `treat_prob_failure_fexi_late_s2`
- Description of variable: Probability of relapse (treatment failure) - fexinidazole for late S2 disease
- Source: [19]
- Country of estimate: NA
- Statistical distribution and parameters: Beta(43.56, 300.11)
- Summary statistics (mean and 95% CI or fixed value): 0.13 (0.09, 0.17)

**Notes**

Mesu et al have published the only study on fexinidazole treatment effectiveness on late-stage 2 cases. Moreover, the accompanying meta-analysis for the WHO treatment guidelines released in 2019 show the outcomes an additional extension study on stage 1, both early and late-stage 2 disease ([19]).

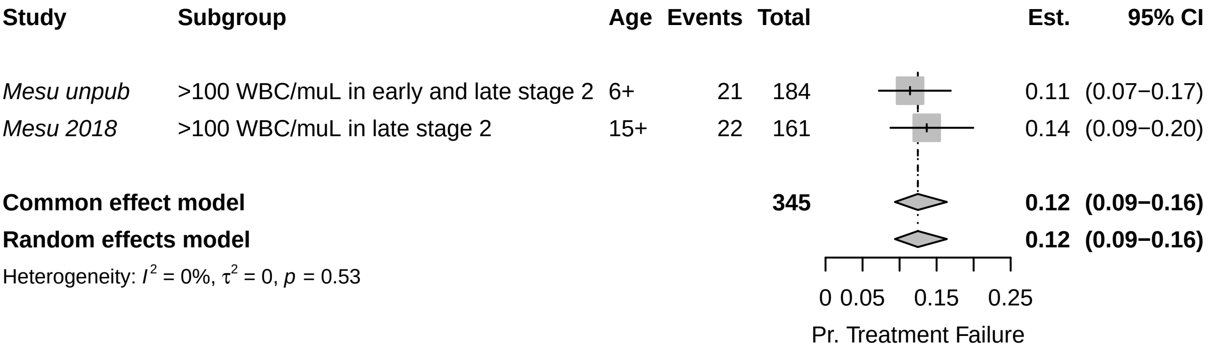


Because the data showed non-significant heterogeneity according to the tau-squared test for heterogeneity, we have chosen to use the fixed-effects combined estimate: 0.125 (0.094, 0.164). The beta parameters of the random-effects estimate Beta(43.56, 300.11) for the probability of severe adverse effects attributable to NECT administration.

##### S4.6.7 treat_prob_failure_nect_s2

- Name in the code: `treat_prob_failure_nect_s2`
- Description of variable: Probability of relapse (treatment failure): NECT for S2 disease
- Source: [14–17, 20, 22]
- Country of estimate: NA
- Statistical distribution and parameters: Beta(15.87, 378.55)
- Summary statistics (mean and 95% CI or fixed value): 0.04 (0.02, 0.06)

**Notes**

The WHO guidelines for treatment of HAT in 2019 [19] presented existing data on treatment failure of NECT. Kansiime and colleagues [22] also performed a systematic review of studies estimating the outcomes of NECT treatment. To produce one comprehensive estimate of treatment failure, we performed a meta-analysis on proportions within single groups.

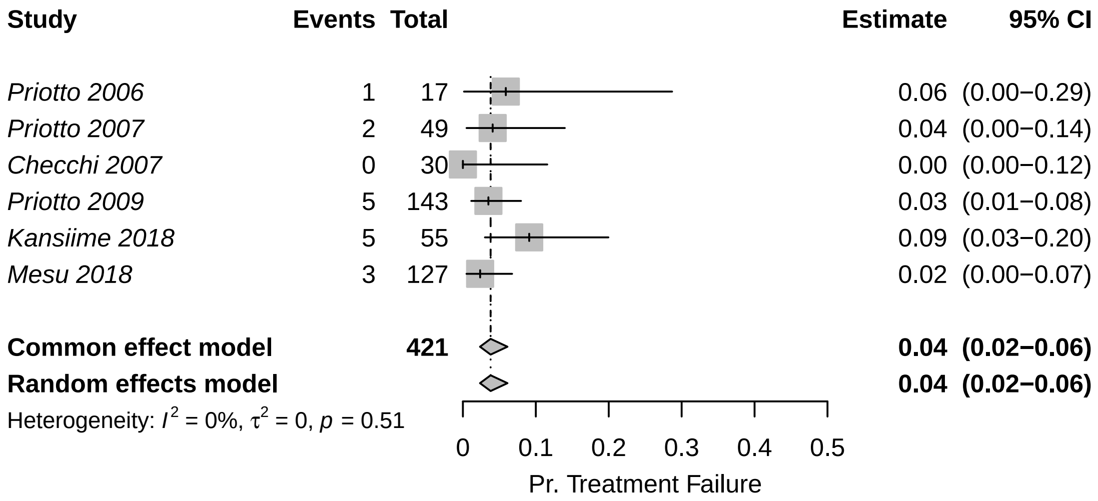


Because the data showed non-significant heterogeneity according to the tau-squared test for heterogeneity, we have chosen to use the fixed-effects estimate, to which we assigned a distribution of Beta(15.87, 378.55)

##### S4.6.8 treat_prob_failure_pent_s1

- Name in the code: `treat_prob_failure_pent_s1`
- Description of variable: Probability of relapse (treatment failure): pentamidine for S1 disease
- Source: [12, 18, 23–26]
- Country of estimate: NA
- Statistical distribution and parameters: Beta(50.3, 665.48)
- Summary statistics (mean and 95% CI or fixed value): 0.07 (0.05, 0.09)

**Notes**

The WHO guidelines for treatment of HAT in 2019 [19] presented existing data on treatment failure of pentamidine treatment. To produce one comprehensive estimate of treatment failure, we performed a meta-analysis on proportions within a single group.

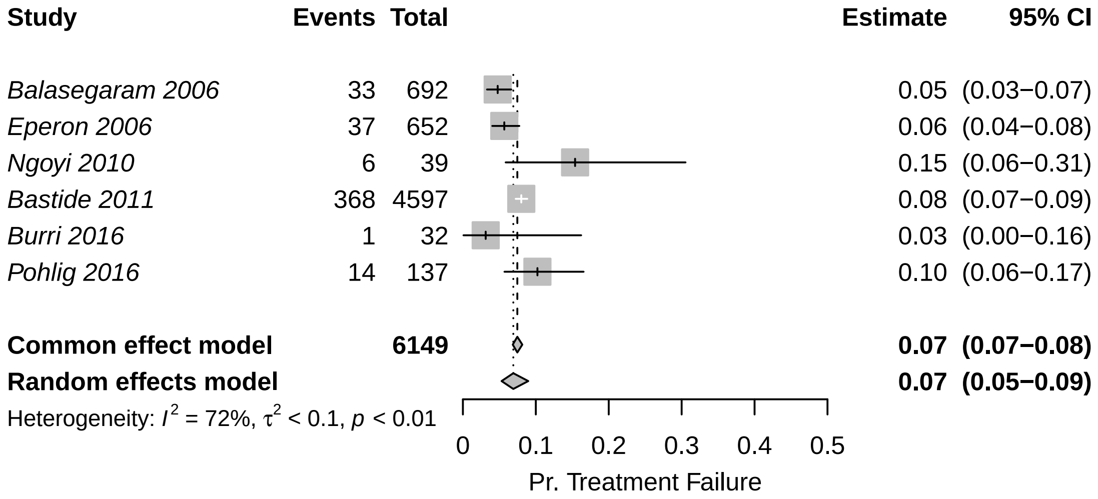


Because the data showed significant heterogeneity according to the tau-squared test for heterogeneity, we have chosen to use the random-effects estimate, to which we assigned a distribution of Beta(50.30, 665.47).

##### S4.6.9 treat_prob_sae_fexi

- Name in the code: `treat_prob_sae_fexi`
- Description of variable: Serious adverse events (SAE): fexinidazole treatment
- Source: [20]
- Country of estimate: NA

- Statistical distribution and parameters: Beta(3, 261)
- Summary statistics (mean and 95% CI or fixed value): 0.01 (<0.01, 0.03)

**Notes**

There is only one published study on fexinidazole, so the probability of serious adverse events will be parameterized with the observations from that study: 4 adverse events attributable to fexinidazole in 3 people among 264 people, so we have assigned a distribution of Beta(3, 261).

There were additional data reported in the appendix to the WHO's interim guidelines ([19]) related to studies that are ongoing. However, since we do not know details about whether those SAEs were attributable to treatment, we have chosen to omit those data.

##### S4.6.10 treat_prob_sae_nect_s2

- Name in the code: `treat_prob_sae_nect_s2`
- Description of variable: Serious adverse events (SAE): NECT treatment
- Source: [14–17, 20, 22]
- Country of estimate: NA
- Statistical distribution and parameters: Beta(40.88, 367.8)
- Summary statistics (mean and 95% CI or fixed value): 0.10 (0.07, 0.13)

**Notes**

The WHO guidelines for treatment of gHAT in 2019 [19], presented all NECT studies to date, as did Kansiime and colleagues [22]. We searched through these studies for evidence of the probability of severe adverse vents (SAEs),descrived as events of Grade 3 or higher according to the National Cancer Institute Common Toxicity Criteria for Adverse Events (CTCAE).

To produce one comprehensive estimate of the probability of SAE, we performed a meta-analysis on proportions within single groups.

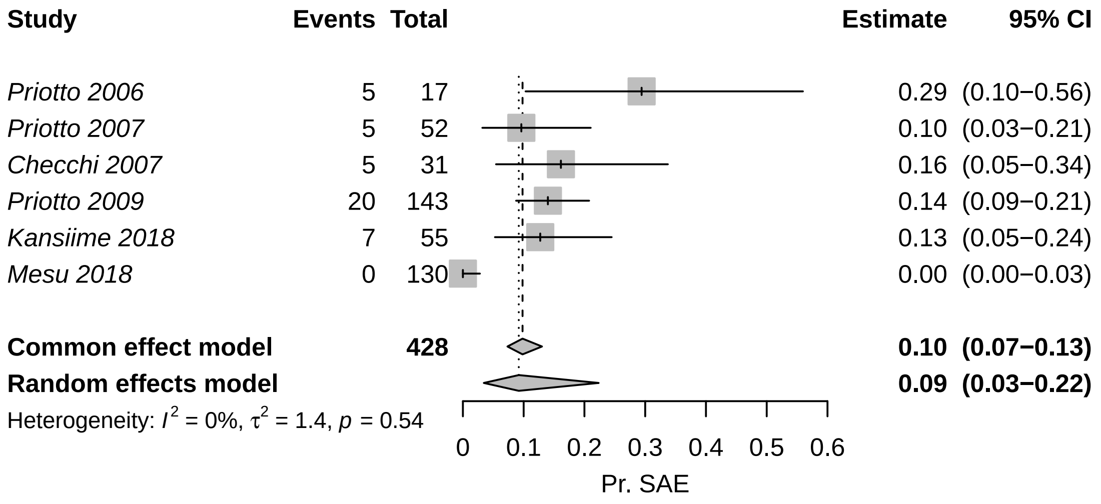


Because the data showed non-significant heterogeneity according to the tau-squared test for heterogeneity, we have chosen to use the fixed-effects estimate: 0.098 (0.073, 0.130), which would result from a beta distribution of Beta(40.88, 367.80).

##### S4.6.11 treat_prob_sae_pent_s1

- Name in the code: `treat_prob_sae_pent_s1`
- Description of variable: Serious adverse events (SAE): pentamidine treatment
- Source: [12, 23, 24]
- Country of estimate: NA
- Statistical distribution and parameters: Beta(1.43, 551.42)
- Summary statistics (mean and 95% CI or fixed value): 0.003 (<0.001, 0.008)

**Notes**

As part of the WHO guidelines for treatment of HAT in 2019 ([19]), Cochrane performed a systematic review of studies that evaluated the efficacy of NECT compared to fexinidazole studies and presented the probability of serious adverse events. Severe or serious adverse events in studies for S1 treatment were defined as “significant hazard, contra- indication, side effect, or precaution” [12, 23, 24].

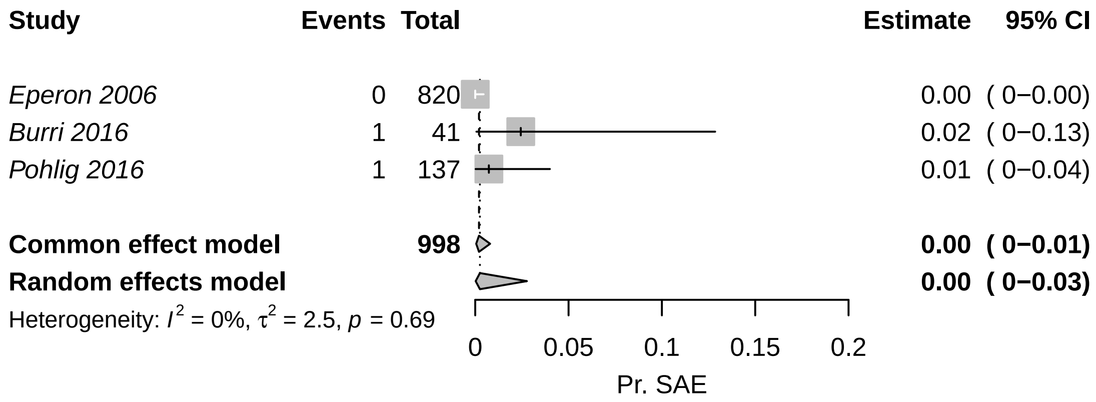


Because the results do not contain evidence of significant heterogeneity, we have chosen to use the fixed (pooled) estimate of 0 (0-0.01), which would result from a beta distribution of Beta(1.43, 551.42).

##### S4.6.12 treat_prob_under35kg

- Name in the code: `treat_prob_under35kg`
- Description of variable: Percent cases under 35 kg as a proportion of all cases over the age of 6
- Source: [13–18, 20, 22–24]
- Country of estimate: NA
- Statistical distribution and parameters: Beta(8.3, 359.6)
- Summary statistics (mean and 95% CI or fixed value): 0.02 (<0.01, 0.04)

**Notes**

To determine whether a patient is eligible for fexinidazole treatment, we could not find any studies that would tell us the number of HAT patients who weighed less than 35 kg, but we have estimated the number of people that might weigh less than 35 kg by examining the distribution of weight among patients in the trials in the literature. Furthermore, we have examined how this variable is related to potential selection by age if the study population. We are interested in the proportion of older children and adults that might weigh less than 35 kg, as age under 6 is a contraindication for fexinidazole.

We fit a gamma distribution by the method of moments to the reported mean and standard deviations of each of the studies. For Priotto 2012, no SD was reported, but an inter-quartile range was reported, so we fit a gamma distribution by the method in Cook [3].

We then took the expected number of people under 35 kg, and then performed a single-proportion meta-analysis with the expected number of people in each study under and over the 35 kg-threshold.

```
## Warning: Use argument 'subgroup' instead of 'byvar' (deprecated).
```

| Citation | Group | Age Group | Mean weight | Measure of spread | No. of observations | Gamma distr. alpha par. | Gamma distr. beta par. | Prop. <35kg | Simulated No. <35kg |
|---|---|---|---|---|---|---|---|---|---|
| Priotto 2006 | Melarsoprol and Nifurtimox | All ages | 49.20 | SD = 14.4 | 18 | 11.67 | 4.21 | 0.16 | 3 |
| Priotto 2006 | Melarsoprol and Eflornithine | All ages | 50.00 | SD = 10.3 | 19 | 23.56 | 2.12 | 0.06 | 1 |
| Priotto 2006 | NECT | All ages | 51.40 | SD = 8.4 | 17 | 37.44 | 1.37 | 0.02 | 0 |
| Priotto 2007 | NECT | Over 15 years old | 51.70 | SD = 7.4 | 52 | 48.81 | 1.06 | 0.01 | 0 |
| Priotto 2007 | Eflornithine | Over 15 years old | 53.10 | SD = 7.2 | 51 | 54.39 | 0.98 | 0.00 | 0 |
| Checchi 2007 | NECT | All ages | 44.80 | SD = 15.1 | 31 | 8.80 | 5.09 | 0.28 | 9 |
| Priotto 2009 | NECT | Over 15 years old | 53.00 | SD = 8.7 | 143 | 37.11 | 1.43 | 0.01 | 2 |
| Priotto 2009 | Eflornithine | Over 15 years old | 53.90 | SD = 8.3 | 143 | 42.17 | 1.28 | 0.01 | 1 |
| Ngoyi 2010 | Pentamidine and Melarsoprol | Over 12 years old | 56.00 | SD = 10.0 | 360 | 31.36 | 1.79 | 0.01 | 3 |
| Priotto 2012 | Single arm | All ages | 49.00 | IQR: 40-56 | 2190 | 16.37 | 2.96 | 0.12 | 265 |
| Schmid 2012 | Single arm | All ages | 45.00 | SD = 16.0 | 629 | 7.91 | 5.69 | 0.29 | 182 |
| Burri 2016 | Pentamidine | Over 15 years old and >35 kg | 48.50 | SD = 7.6 | 40 | 40.83 | 1.19 | 0.03 | 1 |
| Pohlig 2016 | Pentamidine | Over 12 years old and >30 kg | 45.70 | SD = 7.8 | 137 | 34.15 | 1.34 | 0.08 | 10 |
| Pohlig 2016 | Pafuramidine | Over 12 years old and >30 kg | 44.70 | SD = 7.9 | 136 | 32.02 | 1.40 | 0.10 | 14 |
| Kansiime 2018 | All | Over 15 years old | 51.69 | SD = 9.7 | 109 | 28.22 | 1.83 | 0.03 | 3 |
| Mesu 2018 | NECT | Over 15 years old | 50.70 | SD = 9.6 | 130 | 27.89 | 1.82 | 0.04 | 5 |
| Mesu 2018 | Fexinidazole | Over 15 years old | 50.50 | SD = 8.2 | 264 | 37.93 | 1.33 | 0.02 | 5 |

| Study | Arm | Events | Total | Est. | 95% CI |
|---|---|---|---|---|---|
| **all_ages_lbl = Adolescents and adults** | | | | | |
| *Priotto 2007* | NECT | 0 | 52 | 0.00 | (0.00−0.07) |
| *Priotto 2007* | Eflornithine | 0 | 51 | 0.00 | (0.00−0.07) |
| *Priotto 2009* | NECT | 2 | 143 | 0.01 | (0.00−0.05) |
| *Priotto 2009* | Eflornithine | 1 | 143 | 0.01 | (0.00−0.04) |
| *Ngoyi 2010* | Pentamidine and Melarsoprol | 3 | 360 | 0.01 | (0.00−0.02) |
| *Burri 2016* | Pentamidine | 1 | 40 | 0.03 | (0.00−0.13) |
| *Pohlig 2016* | Pentamidine | 10 | 137 | 0.07 | (0.04−0.13) |
| *Pohlig 2016* | Pafuramidine | 14 | 136 | 0.10 | (0.06−0.17) |
| *Kansiime 2018* | All | 3 | 109 | 0.03 | (0.01−0.08) |
| *Mesu 2018* | NECT | 5 | 130 | 0.04 | (0.01−0.09) |
| *Mesu 2018* | Fexinidazole | 5 | 264 | 0.02 | (0.01−0.04) |
| **Common effect model** | | | **1565** | **0.03** | **(0.02−0.04)** |
| **Random effects model** | | | | **0.02** | **(0.01−0.04)** |
| Heterogeneity: $I^2 = 70\%$, $\tau^2 = 0.8$, $p < 0.01$ | | | | | |
| **all_ages_lbl = All ages** | | | | | |
| *Priotto 2006* | Melarsoprol and Nifurtimox | 3 | 18 | 0.17 | (0.04−0.41) |
| *Priotto 2006* | Melarsoprol and Eflornithine | 1 | 19 | 0.05 | (0.00−0.26) |
| *Priotto 2006* | NECT | 0 | 17 | 0.00 | (0.00−0.20) |
| *Checchi 2007* | NECT | 9 | 31 | 0.29 | (0.14−0.48) |
| *Priotto 2012* | Single arm | 265 | 2190 | 0.12 | (0.11−0.14) |
| *Schmid 2012* | Single arm | 182 | 629 | 0.29 | (0.25−0.33) |
| **Common effect model** | | | **2904** | **0.16** | **(0.15−0.17)** |
| **Random effects model** | | | | **0.15** | **(0.08−0.27)** |
| Heterogeneity: $I^2 = 95\%$, $\tau^2 = 0.4$, $p < 0.01$ | | | | | |

Test for subgroup differences (fixed effect): $\chi^2_1 = 135.06$, df = 1 ($p < 0.01$)
Test for subgroup differences (random effects): $\chi^2_1 = 17.61$, df = 1 ($p < 0.01$)

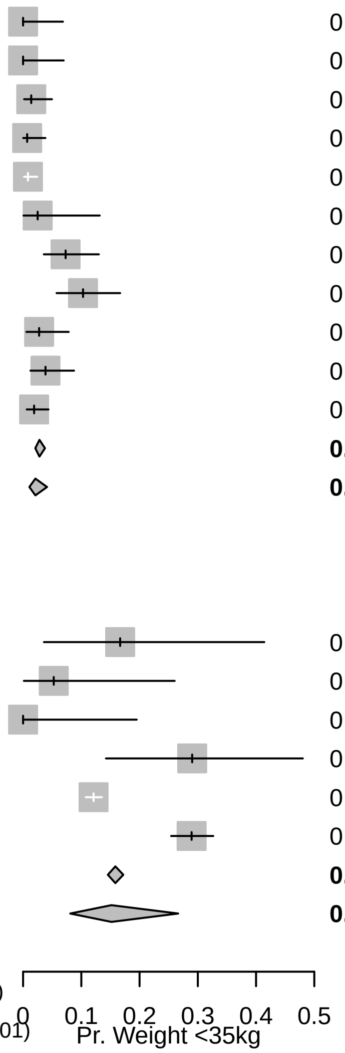


Because the data showed significant heterogeneity according to the tau-squared test for heterogeneity, we have chosen to use the random-effects estimate: 0.02 (0.01-0.04), represented by probability distribution: Beta(8.30, 359.61).

##### S4.6.13 treat_prob_under6yo

- Name in the code: `treat_prob_under6yo`
- Description of variable: Percent cases under 6 as a proportion of all cases
- Source: [12, 13]
- Country of estimate: NA
- Statistical distribution and parameters: Beta(152.53, 2427.9)
- Summary statistics (mean and 95% CI or fixed value): 0.06 (0.05, 0.07)

**Notes**

There were only two studies where the number of children under 5 or 6 years of age was stated explicitly [12, 13].

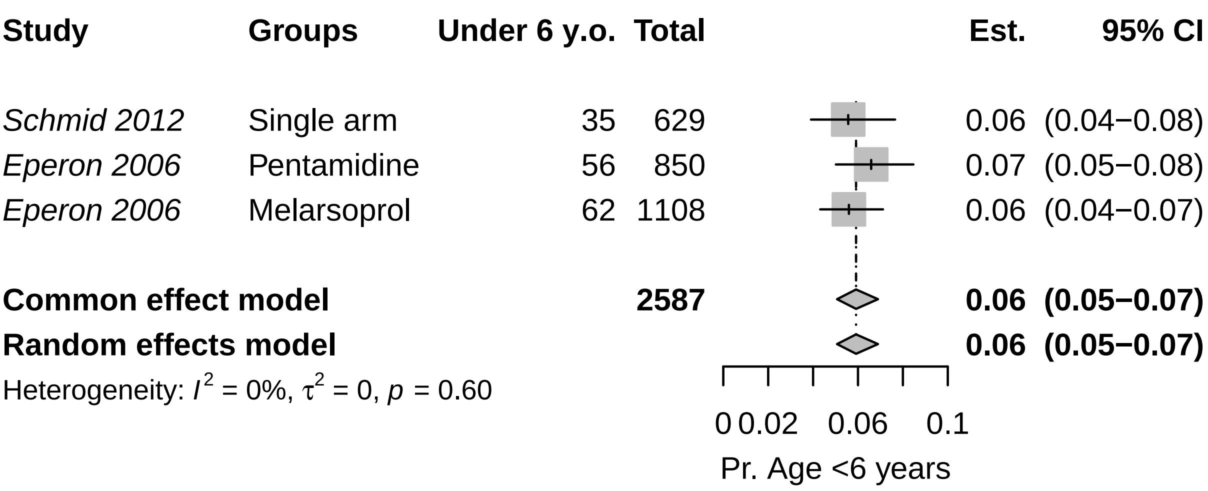


Because the data showed non-significant heterogeneity according to the tau-squared test for heterogeneity, we have chosen to use the fixed-effects combined estimate: 0.059 (0.051, 0.069). The beta parameters of the random-effects estimate Beta(153.53, 2427.90) for the probability of that a patient is under 6 years old.

#### S4.7 Notes on Life-years lost (DALY) Parameters

##### S4.7.1 disability_weighting_s1

- Name in the code: `disability_weighting_s1`
- Description of variable: Disability weights: stage 1 (S1) disease
- Source: [27]
- Country of estimate: NA
- Statistical distribution and parameters: Beta(22.96, 147.21)
- Summary statistics (mean and 95% CI or fixed value): 0.14 (0.09, 0.19)

**Notes**

The Global Burden of Disease listed the impact of sleeping sickness as equivalent to the health state labeled "Motor plus cognitive impairments, severe" and their estimate for a disability weight is 0.542 (0.374-0.702), using the 2013 weight values. No distinction was made between stage 1 and 2 of the disease.

While this seems appropriate for the second stage of sleeping sickness, for stage 1 disability we chose to use the disability weights for equivalent to "infectious disease, acute episode, severe", which is described as "has a high fever and pain, and feels very weak, which causes great difficulty with daily activities" and has a much lower disability weight equivalent to 0.133 (0.088-0.190). The distribution for this parameter is therefore Beta(22.96, 147.21).

It should be noted that other cost-effectiveness analyses have used different values for disability weights [28–30]. These values arise from the 1994 Global Burden of Disease Study but we prefer to consider updated values. Since most of the disability is due to deaths rather than illness during life, we do not believe that this difference is cause for concern.

##### S4.7.2 disability_weighting_s2

- Name in the code: `disability_weighting_s2`
- Description of variable: Disability weights: stage 2 (S2) disease
- Source: [27]
- Country of estimate: NA
- Statistical distribution and parameters: Beta(18.37, 15.63)
- Summary statistics (mean and 95% CI or fixed value): 0.54 (0.37, 0.70)

**Notes**

The Global Burden of Disease listed the impact of sleeping sickness as equivalent to the health state labeled "Motor plus cognitive impairments, severe" and their estimate for a disability weight is 0.542 (0.374-0.702), using the 2013 weight values. No distinction was made between stage 1 and 2 of the disease. The distribution for the parameter is Beta(18.37, 15.63).

It should be noted that other cost-effectiveness analyses have used different values for disability weights [28–30]. These values arise from the 1994 Global Burden of Disease Study but we prefer to consider updated values. Since most of the disability is due to deaths rather than illness during life, we do not believe that this difference is cause for concern.

##### S4.7.3 disability_weighting_sae

- Name in the code: `disability_weighting_sae`
- Description of variable: Disability weights: serious adverse events (SAE)
- Source: [27]
- Country of estimate: NA
- Statistical distribution and parameters: Uniform(0.04, 0.11)
- Summary statistics (mean and 95% CI or fixed value): 0.08 (0.04, 0.11)

**Notes**

As far as we are aware, no one has considered the disability due to severe adverse events attributable to gHAT treatment, but the most common adverse events are gastrointestinal problems and headaches.

We consulted the Global Burden of Disease for disability weights. The health state labeled "symptomatic tension-type headache" was described as "moderate headache that also affects the neck, which causes difficulty in daily activities" and was estimated to have a disability weight equal to 0.037 (0.022–0.057). The health state labeled "moderate symptomatic gastritis and duodenitis without anemia" was described as "abdominopelvic problem, moderate has pain in the belly and feels nauseous; the person has difficulties with daily activities" and and was estimated to have a disability weight equal to 0.114 (0.078–0.159).

Our distribution is therefore Uniform 0.037-0.114. Since most of the disability is due to deaths rather than illness during life, we do not believe that the uncertainty in this parameter is cause for concern for the purpose of the conclusions of this analysis.

##### S4.7.4 age_of_death

- Name in the code: `age_of_death`
- Description of variable: Age of death from infection (for years of life lost per fatal case)
- Source: [12–18, 20–25, 31]
- Country of estimate: NA
- Statistical distribution and parameters: Gamma(148, 0.18)
- Summary statistics (mean and 95% CI or fixed value): 26.65 (22.56, 31.07)

**Notes**

No good registry of age of infection exists, so we have searched through the literature that we have used to parameterize the model for the average age of HAT patients. Among the studies that we have used to inform other parameters, eight studies reported age information in a sample of patients of all ages, and nine studies have reported the age information in a sample of older children or adults (12-15 years and older).

However, the data exists in a state that is difficult to synthesize:

Therefore, we have fit a gamma distribution to the means and medians of the studies that included patients of all ages. We have omitted the median from Alirol 2013 as this median seems unusually high – even higher than the mean age of patients in studies were only adults (over the age of 15) were recruited. Our distribution is therefore Gamma(147.93, 0.18) with a mean and 95% confidence interval of 27 (23, 31), which provides a sufficiently large bound of uncertainty in lieu of the information we have.

#### S4.8 Notes on Screening and Vector Control Cost Parameters

##### S4.8.1 as_cost_moto_capital

- Name in the code: `as_cost_moto_capital`
- Description of variable: Active screening: capital costs of a motorcycle team
- Source: Program data
- Country of estimate: COD
- Statistical distribution and parameters: Gamma(8.47, 1000.81)
- Summary statistics (mean and 95% CI or fixed value): 8,489 (3,742, 14,809)

| Citation | Group | Age Group | Summary |
|---|---|---|---|
| Priotto 2006 | Melarsoprol and Nifurtimox | All ages | Mean: 29.1 range: 5-56 |
| Priotto 2006 | Melarsoprol and Eflornithine | All ages | Mean: 28.1 range: 11-61 |
| Priotto 2006 | NECT | All ages | Mean: 29.1 range: 9-62 |
| Balasegaram 2006 | Pentamidine | All ages | 148 under 15 and 504 over 15 |
| Eperon 2006 | Pentamidine | All ages | 56 patients 0-5, 226 patients 6-15, 568 patients 15+ |
| Eperon 2006 | Melarsoprol | All ages | 63 patients 0-5, 249 patients 6-15, 796 patients 15+ |
| Priotto 2007 | NECT | Over 15 years old | Mean: 33.1 range: 15-69 |
| Priotto 2007 | Eflornithine | Over 15 years old | Mean: 36.1 range: 15-70 |
| Checchi 2007 | NECT | All ages | Mean: 23.9 range: 4-45 |
| Priotto 2009 | NECT | Over 15 years old | Mean: 32.8 SD: 12.5 |
| Priotto 2009 | Eflornithine | Over 15 years old | Mean: 34.6 SD: 13.5 |
| Ngoyi 2010 | Pentamidine | Over 12 years old | Mean: 35 SD: 13 |
| Ngoyi 2010 | Pentamidine and Melarsoprol | Over 12 years old | Mean: 34 SD: 12 |
| Priotto 2012 | Single arm | All ages | Median: 24 IQR: 15-35 |
| Schmid 2012 | Single arm | All ages | 35 patients 0-4 yo, 65 patients 5-11 yo, and 529 patients 12 yo or more. |
| Hasker 2012 | All | All ages | Median: 27 IQR: 16-40 |
| Alirol 2013 | Single arm | All ages | Median: 36 IQR: 20-50 |
| Burri 2016 | Pentamidine | Over 15 years old and >35 kg | Median: 31 range: 15-50 |
| Pohlig 2016 | Pentamidine | Over 12 years old and >30 kg | Median: 31 range: 13-75 |
| Pohlig 2016 | Pafuramidine | Over 12 years old and >30 kg | Median: 30 range: 12-64 |
| Kansiime 2018 | NECT | Over 15 years old | Mean: 27.23 SD: 12.07 |
| Kansiime 2018 | Eflornithine | Over 15 years old | Mean: 27.33 SD: 8.59 |
| Mesu 2018 | NECT | Over 15 years old | Mean: 35.2 SD: 13.2 |
| Mesu 2018 | Fexinidazole | Over 15 years old | Mean: 34.5 SD: 12.6 |

**Notes**

According to Snijders et al [10], the capital cost of an active surveillance team was $11,406 in 2018 for a team that administer CATT.

Because in Mandoul active screening occurs for at most 1 months of the year, and in DRC the screening occurs for 11 months of the year, we will take the 1/11th of the cost that does not include training (as most capital for active screening is shared with other programs.) To account for training, which is program-specific, we will leave 20% of the cost intact, and take only 1/11th of the rest of the capital.

Because we had no sense of the uncertainty in capital costs, we assigned a gamma distribution that had a confidence interval that spanned half the estimate and twice the estimate, yielding a distribution of Gamma(8.47, 1152.75) with a sample of 9,823 (4,394, 17,514).

##### S4.8.2 as_cost_moto_followup

- Name in the code: `as_cost_moto_followup`
- Description of variable: Active screening: motorcycle-based follow-up of patients
- Source: Program data
- Country of estimate: TCD
- Statistical distribution and parameters: Gamma(70.05, 54.93)
- Summary statistics (mean and 95% CI or fixed value): 3,843 (2,992, 4,782)

**Notes**

Program documents showed that the motorcycle teams in Mandoul cost about $3000 per year. Because we had no sense of the uncertainty in capital costs, we assigned a gamma distribution that had a confidence interval that spanned half the estimate and twice the estimate, yielding a distribution of Gamma(8.47, 398.36) with a sample of 3,355 (1,489, 5,861).

##### S4.8.3 as_cost_moto_management

- Name in the code: `as_cost_moto_management`
- Description of variable: Active screening: fixed management costs of a motorcycle team
- Source: Program data
- Country of estimate: TCD
- Statistical distribution and parameters: Gamma(70.05, 91.56)
- Summary statistics (mean and 95% CI or fixed value): 6,411 (5,017, 7,965)

**Notes**

Program documents showed that the motorcycle teams in Mandoul cost about $6000 per year. Because we had no sense of the uncertainty in capital costs, we assigned a gamma distribution that had a confidence interval that spanned half the estimate and twice the estimate, yielding a distribution of Gamma(8.47, 796.72) with a sample of 6,763 (2,950, 12,038).

##### S4.8.4 as_cost_team_capital

- Name in the code: `as_cost_team_capital`
- Description of variable: Active screening: capital costs of a traditional team
- Source: Program data
- Country of estimate: COD
- Statistical distribution and parameters: Gamma(200000, 0.02)
- Summary statistics (mean and 95% CI or fixed value): 4,000 (3,983, 4,017)

**Notes**

To our knowledge, no active surveillance costs have been estimated via detailed costing study in Mandoul or Chad more broadly [32]. One study reported variable costs of active screening [7], but not the capital investments nor supervision.

Lutumba and colleagues [33] calculated that the total cost of screening 40,000 patients in 2003 was 46,734.29 Euros (1 Euro = 0.86 USD in 2003) (see table 3 of [33]). Of that value, 21 percent of the costs were capital costs, or a cost of 9258.7 in 2020 USD. The publication did not indicate whether that value was annualized or not.

According to Bessel and colleagues [30] the cost for a mobile team that screens 250 patients per day for 220 days a year has capital investments (annualized for five years) of $12,000 for a team that administers CATT and $12,781 for a team that administers RDT (in 2013 USD) (see [30] Table S1). Adjusting for inflation, that would equal 10923.56 in 2020 USD.

According to Snijders et al [10], the capital cost of an active surveillance team was $11,406 in 2018 for a team that administer CATT.

Because in Mandoul active screening occurs for at most 1 months of the year, and in DRC the screening occurs for 11 months of the year, we will take the 1/11th of the cost that does not include training (as most capital for active screening is shared with other programs.) To account for training, which is program-specific, we will leave 20% of the cost intact, and take only 1/11th of the rest of the capital.

Although no publication gave us a sense of the uncertainty in capital costs, but since we had three studies, we assumed that the range was equivalent to the 95% confidence interval. Therefore, our probability distribution is Gamma(199999.99, 0.02), which yields a distribution with mean and confidence intervals of 3,192 (3,178, 3,206).

##### S4.8.5 as_cost_team_management

- Name in the code: `as_cost_team_management`
- Description of variable: Active screening: fixed management costs of a traditional team
- Source: Program data
- Country of estimate: TCD
- Statistical distribution and parameters: Gamma(8.47, 2167)
- Summary statistics (mean and 95% CI or fixed value): 18,361 (8,308, 32,750)

**Notes**

To our knowledge, no active surveillance costs have been estimated via detailed costing study [32]. However, we know that the variable recurrent costs in 2009 were 5544000 [7]. We add to that the cost of supervision, which we will assume is the same as the cost of supervision of the passive screening activities (5.9M CFA in 2019 for the whole country, and we assume a third is due to activities in Mandoul). Therefore, variable recurrent costs are 12750.7509 in 2020 USD, and supervision is 3569.3389 in 2020 USD.

To parameterize the model, we assign a distribution with 95% confidence intervals equal to half and double the costs. The distribution is Gamma(8.47, 2167.10), which yields a distribution with median and confidence intervals of 18,339 (8,042, 32,743).

##### S4.8.6 dx_cost_catt

- Name in the code: `dx_cost_catt`
- Description of variable: CATT algorithm: cost per test used
- Source: [10, 30, 32, 33]
- Country of estimate: COD
- Statistical distribution and parameters: Gamma(22.87, 0.02)
- Summary statistics (mean and 95% CI or fixed value): 0.46 (0.29, 0.66)

**Notes**

To our knowledge, costs for CATT tests are only featured in three sources: 1) the WHO, cited by Keating et al 2015 [32], 2) by Lutumba et al [34], and 3) by Bessel et al [30].

1) Keating et al listed the cost of a CATT test at 0.73 in 1998 USD, equivalent to 0.80 in 2020 USD.

2) Lutumba et al listed the cost of a CATT test at 0.52 in 2003 USD, equivalent to 0.42 in 2020 USD.

3) Bessel et al listed the cost of CATT test at 0.70 for a mobile team and 0.89 for a fixed post in 2013 USD. The reason for the difference in price between screening at a mobile team vs screening at a fixed post was due to the differential wastage and was given in the supplement notes: "Costs at mobile teams and fixed units are different because once a bottle of CATT antigen is open repeat cases and controls must be performed. Materials are the cost of test materials plus the cost of the lancet."

In terms of 2020 USD in Chad, the costs are 0.62 for CATT tests in mobile teams 0.78 for CATT tests in fixed posts. We take into account wastage as separate parameter.

4) Snijders et al listed the cost of a CATT test at 0.74 in terms of 2018 USD, which is equivalent to 0.65 in Chad in 2020 [10].

Because we had more than two studies on this parameter, we chose gamma distributions where the 2.5th and the 97.5th percentiles would match the minimum and maximum values in the literature, so our parameter distribution is Gamma(38.71, 0.02), which has a mean and confidence interval of 0.59 (0.42, 0.79).

Considering delivery costs (described in S4.9.4), the cost of CATT tests are 0.84 (0.59, 1.13) in 2018 USD. Considering both delivery costs and wastage, the cost of CATT tests are 0.93 (0.65, 1.25) in active screening teams and 1.08 (0.75, 1.46) in passive surveillance (fixed) posts.

##### S4.8.7 dx_cost_lumbar_exam

- Name in the code: `dx_cost_lumbar_exam`
- Description of variable: Lumbar puncture and laboratory exam: cost per person that needs to be staged
- Source: [10, 30, 35]
- Country of estimate: COD
- Statistical distribution and parameters: Gamma(3.26, 2.87)
- Summary statistics (mean and 95% CI or fixed value): 9.38 (2.11, 21.85)

**Notes**

To our knowledge, costs for lumbar puncture tests were listed in detail only by Bessel and colleagues [30] and Snijders and colleagues [10]. The cost listed by Bessel et al was 2.38 in terms of 2013 USD, equivalent to 1.42 in 2018 USD. Snijders and colleagues report a cost of 23.02 in 2018 USD. Irurzun-Lopez [35] have reported a similar value, so we will assume a value equal to that of Snijders and colleages.

Because we had more than two studies to inform this parameter, we chose gamma distributions where the 2.5th and the 97.5th percentiles would match the minimum and maximum values in the literature, so our parameter distribution is Gamma(2.42, 3.66), which yields a distribution with mean and confidence intervals of 8.86 (1.44, 23.01).

##### S4.8.8 dx_cost_mAECT

- Name in the code: `dx_cost_mAECT`
- Description of variable: mAECT diagnosis: cost
- Source: [[33]; Bessel2018; [10]]
- Country of estimate: COD
- Statistical distribution and parameters: Gamma(8.47, 0.23)
- Summary statistics (mean and 95% CI or fixed value): 1.95 (0.87, 3.51)

**Notes**

UNDER CONSTRUCTION

##### S4.8.9 dx_cost_LAMP

- Name in the code: `dx_cost_LAMP`
- Description of variable: LAMP diagnosis: cost
- Source: [36]
- Country of estimate: UGA
- Statistical distribution and parameters: Gamma(8.47, 0.62)
- Summary statistics (mean and 95% CI or fixed value): 5.27 (2.32, 9.41)

**Notes**

To our knowledge, costs for LAMP tests were listed in Wastling et al (2010 PLoS NTD). The cost listed was 3.53 USD (2010) and in terms of 2020 USD costs the per-test cost is 4.70.

Because we have no sense of the uncertainty, we will assign a Gamma distribution where the credible intervals are equal to half and double the estimate. herefore, our probability distribution is Gamma(8.47, 0.62), which yields a distribution with median and confidence intervals of 5.30 (2.33, 9.35).

##### S4.8.10 dx_cost_microscopy

- Name in the code: `dx_cost_microscopy`
- Description of variable: Microscopy (Bloodsample, LNA, mAECT): cost per person that needs to be confirmed

- Source: [11]
- Country of estimate: COD
- Statistical distribution and parameters: Gamma(8.47, 1.27)
- Summary statistics (mean and 95% CI or fixed value): 10.70 (4.75, 18.98)

**Notes**

The per-patient price to confirm a patient with a full microscopy procedure is reported in an upcoming publication by Snijders et al [11]. They report that a microscopy procedure constituted of mAECT and LNA (lymph node aspiration) costs 9.53 in 2018 USD, or $8.13 in 2020 USD in Chad. Because we had no report of the standard error around that estimate, we assigned a gamma distribution that had a confidence interval that spanned half the estimate and twice the estimate, yielding a distribution of Gamma(8.47, 1.27) with a sample of 10.73 (4.73, 19.08).

##### S4.8.11 dx_cost_rdt

- Name in the code: `dx_cost_rdt`
- Description of variable: RDT algorithm: costs per test used
- Source: [11]
- Country of estimate: NA
- Statistical distribution and parameters: Gamma(8.47, 0.19)
- Summary statistics (mean and 95% CI or fixed value): 1.60 (0.72, 2.84)

**Notes**

To our knowledge, costs for RDT tests were listed in Sutherland et al [29], Bessel et al [30], and an upcoming publication by Snijders and colleagues [11].

1) Both Sutherland et. al and Bessel et. al coincided on a cost of 0.50 USD 2013 for the test in the international market (after a 25 cent subsidy paid for outside of DRC). Bessel also calculated staff costs, shipment, and wastage for 0.32 USD 2013. Because we can split the cost between tradable and non-tradable costs, we inflate and adjust the costs for the staff costs and shipment and then add the cost of the RDT. In terms of 2020 USD costs, the shipment and the staff costs are 0.17, and the total cost is 0.92.

2) Snijders et. al reported a cost between 0.85 and 1.97, depending on the company from which the test is purchased. It should be noted that no single company produces enough tests for any single intervention, and so a range of prices must be considered. Snijders' estimate does not include delivery or wastage, so we take that into account separately.

It appears difficult to compare the three estimates, so we will take Snijders' estimate from a recent micro-costing analysis. The mean was $1.59, and we assign a distribution with confidence intervals equal to half and double the costs. The distribution is Gamma(8.47, 0.19) with a sample of 1.59 (0.71, 2.79). No conversion to Chadian price levels was done because tests are bought and sold in the international market.

Considering delivery costs (described in S4.9.4), the cost of RDT tests are 2.30 (1.03, 4.09) in 2020 USD. Considering both delivery costs and wastage, the cost is 2.47 (1.08, 4.45) in passive surveillance teams, and in mini-mobile teams.

##### S4.8.12 ps_cost_facility_capital

- Name in the code: `ps_cost_facility_capital`
- Description of variable: Passive surveillance: capital costs of a facility
- Source: Program data
- Country of estimate: COD
- Statistical distribution and parameters: Gamma(8.47, 216.24)
- Summary statistics (mean and 95% CI or fixed value): 1,826 (798, 3,241)

**Notes**

Capital costs apply to each health center or hospital that is capable of HAT diagnosis.

To our knowledge, no passive screening costs have been estimated via detailed costing study [32]. We took into account the results of a micro-costing study in Yasa Bonga and Mosango [11], two health zones of Kwilu province. The publication reported a cost of 1580 in 2018, which would be equivalent to 1628 in 2020 USD.

To parameterize the model, we assign a distribution with 95% confidence intervals equal to half and double the costs. The distribution is Gamma(8.47, 216.24), which yields a distribution with median and confidence intervals of 1,749 (809, 3,224).

##### S4.8.13 ps_cost_management

- Name in the code: `ps_cost_management`
- Description of variable: Passive surveillance: fixed recurrent management costs
- Source: Program data
- Country of estimate: TCD
- Statistical distribution and parameters: Gamma(32.47, 12.18)
- Summary statistics (mean and 95% CI or fixed value): 396.09 (271.96, 542.85)

**Notes**

Recurrent management costs are fixed at the facility level.

To our knowledge, no passive screening costs have been estimated via detailed costing study [32]. But program estimates say that it costs about 5.9M CFA to manage the passive screening system and 2.9M CFA for sensitization of the personnel for the whole country (unpublished). In 2019, there were 54 clinics in the country, 23 of which were in Mandoul, so we will attribute a proportional cost for supervision and sensitization to the focus. In USD, this equals $325.54.

To parameterize the model, we assign a distribution with confidence intervals, since the estimate is from multiple years of the program costs (unpublished). The distribution is Gamma(32.47, 12.18), which yields a distribution with median and confidence intervals of 392 (271, 538).

##### S4.8.14 ps_cost_RDTfacility_capital

- Name in the code: `ps_cost_RDTfacility_capital`
- Description of variable: Capital costs of RDT facilities (training only)
- Source: Program data
- Country of estimate: COD
- Statistical distribution and parameters: Gamma(8.47, 31.97)
- Summary statistics (mean and 95% CI or fixed value): 270.21 (119.69, 477.03)

**Notes**

Capital costs apply to each health center or hospital that is capable of HAT diagnosis. For clinics that do both screening, confirmation and staging, see the section on the parameter 'ps_cost_facility_capital'.

For clinics that are not able to do diagnosis but only RDT screening, the only capital cost is training. In Snijders' et. al 2021, 12,029 2018 USD was spent to train 43 health centers in DRC to do RDT-based screening. In terms of 2020 USD in Chad, this amounts to 239.

To parameterize the model, we assign a distribution with 95% confidence intervals equal to half and double the costs. The distribution is Gamma(8.47, 31.69), which yields a distribution with median and confidence intervals of 259 (122, 480).

##### S4.8.15 vc_cost

- Name in the code: `vc_cost`
- Description of variable: The total cost of vector control
- Source: [37]
- Country of estimate: NA
- Statistical distribution and parameters: Gamma(8.47, 8132)
- Summary statistics (mean and 95% CI or fixed value): 68,917 (30,975, 121,361)

**Notes**

To our knowledge, only one vector control micro-costing study has been performed by Rayaisse and colleagues [37] in Mandoul, Chad, which reported a cost of 56,000 2016 USD. The equivalent cost in 2020 USD for Chad is 61,237.91 USD.

As there was no sense of the uncertainty in target deployment costs, we assign a distribution with confidence intervals equal to half and double the costs. The distribution is Gamma(8.47, 8131.61), which yields a distribution with mean and confidence intervals of 69,069.04 (31,216.23-123,746.33).

#### S4.9 Notes on Treatment Cost Parameters

##### S4.9.1 rx_cost_fexinidazole

- Name in the code: `rx_cost_fexinidazole`
- Description of variable: Course of fexinidazole: cost
- Source: [29]
- Country of estimate: Global
- Statistical distribution and parameters: Gamma(100, 0.5)
- Summary statistics (mean and 95% CI or fixed value): 50.00 (40.76, 60.34)

**Notes**

The cost of fexanidozole, for stage 1 and 2 disease. In the near future this will be the drug of choice for first-line treatment for both stages of disease. It may require hospitalization, but eventually it should be taken on an outpatient basis.

##### S4.9.2 rx_cost_nect

- Name in the code: `rx_cost_nect`
- Description of variable: Course of nifurtimox eflornithine combination therapy (NECT): cost
- Source: [38]
- Country of estimate: Global
- Statistical distribution and parameters: Gamma(100, 3.6)
- Summary statistics (mean and 95% CI or fixed value): 360.47 (291.82, 433.21)

**Notes**

This represents the cost of NECT to the capital for stage 2 disease. Simarro and colleagues listed a cost of 1440 USD for treatment of four patients.

Because it is available on the international market, where is it sold in USD, and not subject to the inflationary pressures of any particular country, we have not inflated the cost or converted them to any other currency.

In the future it may be replaced with fexinidozole and this would be the drug for treatment failures or very severe patients.

##### S4.9.3 rx_cost_pentamidine

- Name in the code: `rx_cost_pentamidine`
- Description of variable: Course of pentamidine: cost
- Source: [32]
- Country of estimate: Global
- Statistical distribution and parameters: Gamma(100, 0.54)
- Summary statistics (mean and 95% CI or fixed value): 54.11 (44.12, 65.18)

**Notes**

The cost of pentamidine, for stage 1 disease. Because it is available on the international market, where is it sold in USD, and not subject to the inflationary pressures of any particular country, we have not inflated the cost or converted them to any other currency.

In the future pentamidine treatment it may be replaced with fexinidozole treatment, which would circumvent the need for a lumbar puncture.

##### S4.9.4 rx_delivery_markup

- Name in the code: `rx_delivery_markup`
- Description of variable: Drug delivery mark-up
- Source: [39, 40]
- Country of estimate: TCD
- Statistical distribution and parameters: Uniform(0.15, 0.25)
- Summary statistics (mean and 95% CI or fixed value): 0.20 (0.15, 0.25)

**Notes**

Because we do not know the delivery price of drugs for each country, we have applied the standard value for the mark up of traded goods recommended by the WHO CHOICE program for AFRO E: https://www.who.int/choice/cost-effectiveness/inputs/price_multiplier/en/.

##### S4.9.5 treat_cost_ip_day

- Name in the code: treat_cost_ip_day
- Description of variable: Hospital stay: cost per day
- Source: [1, 39, 40]
- Country of estimate: TCD
- Statistical distribution and parameters: Gamma(5.45, 1.76)
- Summary statistics (mean and 95% CI or fixed value): 9.60 (3.40, 19.44)

**Notes**

We got the estimates of inpatient treatment costs from the 2010 WHO CHOICE cost estimates (recently updated by [39] and [40]). In 2010, a day at a primary hospital in Chad would be 11.90 (4.92, 25.90) I$. In 2010, a day at a secondary hospital would be 12.40 (4.73, 27.86) I$, and a day at a tertiary hospital would be 16.04 (6.37, 36.93) I$.

The equivalent estimates in 2010 USD are 5.84 (2.41, 12.70) per day at a primary hospital, 6.08 (2.32, 13.66) per day at a secondary hospital, and 7.87 (3.12, 18.11) per day at a tertiary hospital. After converting to local currency, applying the inflation index, and converting to 2020 USD, the estimates are 6.17 (2.55, 13.43) per day at a primary hospital, 6.43 (2.45, 14.44) per day at a secondary hospital, and 8.31 (3.30, 19.14) per day at a tertiary hospital.

At the moment, we do not know how many of each kind of hospital the population of HAT patients attend, nor do we understand how costs at district hospitals, referral hospitals, etc resemble those of the three kinds of hospitals under analysis by the WHO CHOICE program. Therefore, we take the estimate with a higher mean (hospital day in a tertiary hospital) in an effort not to under-state the costs of treatment and interventions.

To parameterize the model, we assign a gamma distribution with 95% confidence intervals equal to those reported by WHO CHOICE [1]. The distribution is Gamma(5.45, 1.76), which yields a distribution with median and confidence intervals of 9.63 (3.34, 19.37). Although this yields a higher median, the uncertainty is adequately characterized.

##### S4.9.6 treat_cost_op_visit

- Name in the code: treat_cost_op_visit
- Description of variable: Outpatient consultation: cost
- Source: [1, 39, 40]
- Country of estimate: TCD
- Statistical distribution and parameters: Gamma(2.48, 0.79)
- Summary statistics (mean and 95% CI or fixed value): 1.96 (0.31, 5.05)

**Notes**

We got the estimates of an outpatient consultation from the 2010 WHO CHOICE cost estimates (recently updated by [39] and [40]). In 2010, a consult at a health centre without beds in Chad would be 2.58 (0.53, 7.45) I$. In 2010, and a consult at a health centre with beds in Chad would be 3.19 (0.62, 9.72) I$.

The equivalent estimates in 2010 USD are 1.27 (0.26, 3.65) per day at a health centre without beds, 1.56 (0.30, 4.77) per day at a health centre with beds. After converting to local currency, applying the inflation index, and converting to 2020 USD, the estimates are 1.34 (0.27, 3.86) per consultation at a health centre without beds, and 1.65 (0.32, 5.04) per consultation at a health centre with beds.

At the moment, we do not know how many of each kind of health centre the population of HAT patients attend. Therefore, we take the estimate with a higher mean (health centre with beds) in an effort not to under-state the costs of treatment and interventions.

To parameterize the model, we assign a gamma distribution with 95% confidence intervals equal to those reported by WHO CHOICE [1]. The distribution is Gamma(2.48, 0.79), which yields a distribution with median and confidence intervals of 1.95 (0.33, 5.08). Although this yields a higher median, the uncertainty is adequately characterized.
