## Supplementary material for "Health economic evaluation of strategies to eliminate *gambiense* human African trypanosomiasis in the Mandoul disease focus of Chad": S5 Text. PRIME-NTD Checklist.

### S5. PRIME criteria

| Principle and what has been done to satisfy the principle? | Place in the manuscript |
|---|---|
| **1. Stakeholder engagement**<br>Strategy components were determined along with the country director of PNLTHA, Peka Mallaye and other PNLTHA members, Justin Darnas and Severin Mbainda (co-authors). Implementation of simulations of AS and PS costs were aided by information from Paul Bessell, who has helped maintain the data of surveillance activities by FIND (Foundation for Innovative & Novel Diagnostics) and collaborators at the Liverpool School of Tropical Medicine, who have run field operations in Chad since 2014. | Authorship list and acknowledgements. |
| **2. Complete model documentation**<br>Full model (including the fitting code) and documentation are available through OpenScienceFramework (OSF). The epidemiological model is fully described in the fitting study of Rock *et al* [2] and cost model in S1 Text. | Description in Supplementary Information S1 Text and access the code via `MandoulCEA`[1] and `MandoulReanalysis`[2]. |
| **3. Complete description of data used**<br>Information about the epidemiological case data used for fitting is described in Rock *et al* [2]. The data for the interventions was retrieved from administrative records and is described in Supplementary Information S1 Text, Section S1.2.2. The data used for clinical outcomes and costs were estimates from the literature. Assumptions and estimates were parameterized according to conventions in the economic evaluation literature [3] | For assumptions around intervention, treatment effects and costs, see Supplementary Information S1 Text. |
| **4. Communicating uncertainty**<br>*Structural uncertainty*:<br>Uncertainty arising from the choice of discounting, time horizon are shown by re-running the entire analysis with alternative assumptions. The transmission model outputs are from an ensemble model comprised of various competing model structures, weighted by statistical support for each variant (*e.g.* most outputs do not have any transmission from non-human animal hosts following weighting, although this is included in two model variants). | Main text results and discussion. Time horizons and discounting two-way sensitivity analysis is in Supplementary Information S3 Text, Section S3.3, Figure E. Sensitivity analysis of fexinidazole treatment for those who are eligible rather than only older treatments (pentamidine and NECT) is in Supplementary Information S3 Text, Table B). |
| *Parameter uncertainty*:<br>The epidemiological parameters are the posterior distributions of a model fitted to time-series data, and full details are available in another publication (Rock *et al* [2]). For the parameters to model health outcomes and costs, assumptions and estimates were parameterized according to conventions in the economic evaluation literature [3], taking care to sample from large distributions for aspects for which we knew very little. | Epidemiological parameters are available on OSF `MandoulReanalysis`[2]. Health outcome and cost-effectiveness parameters: see Table 2 and Supplementary information S4 Text |
| *Prediction uncertainty*:<br>Observational uncertainty in epidemiological model predictions. Tables 2 and 3 present both means and 95% prediction intervals. The `GUI`[3] includes box and whisker plots to show uncertainty in cases, deaths and DALYs. We include the probability of meeting EoT by 2030 as well as the expected year of EoT. For cost-effectiveness results, we present the optimal decisions (the probability of each of being cost-effective) at different willingness-to-pay thresholds rather than solely providing ICERs using the net benefits framework (see Tables 3 & 4). | Tables 2-4 of the manuscript and the `GUI`[3]. |
| **5. Testable model outcomes**<br>Epidemiological model outputs are routinely reported metrics of the disease course: detected active and passive case detections. Therefore, these predictions can be compared to future data as long as the data is put into context alongside measures of active screening coverage and the number of fixed health posts equipped for passive screening. Some components of costs predictions can be validated against expenditures, but it must be noted that these are economic costs, and so resource use for which there is no explicit invoice is taken into account as well. | Epidemiological outcomes for each year until 2050 can be viewed in the `GUI`[3]. All the ingredients to the economic costs were shown in detail in the Supplementary Information S1 Text, section S1.3. |

[1] `MandoulCEA` with full address: `https://osf.io/bjxwn/`.
[2] `MandoulReanalysis` with full address: `https://osf.io/rak9d/`.
[3] `GUI` with full address: `https://hatmepp.warwick.ac.uk/MandoulCEA/v1/`

Table 1: PRIME-NTD criteria fulfilment. We summarise how the NTD Modelling Consortium's "5 key principles of good modelling practice" [1] have been met in the present study.
