## Supplementary material for "Health economic evaluation of strategies to eliminate *gambiense* human African trypanosomiasis in the Mandoul disease focus of Chad": S6 Text. CHEERS Checklist.

### S6. CHEERS checklist

| Section/item | Item No | Recommendation | Reported on page no, line no |
|---|---|---|---|
| **Title** | | | |
| Title | 1 | Identify the study as an economic evaluation or use more specific terms such as "cost-effectiveness analysis", and describe the interventions compared. | Cover page, no line number |
| **Abstract** | | | |
| Abstract | 2 | Provide a structured summary of objectives, perspective, setting, methods (including study design and inputs), results (including base case and uncertainty analyses), and conclusions. | Abstract |
| **Introduction** | | | |
| Background and objectives | 3 | Provide an explicit statement of the broader context for the study. Present the study question and its relevance for health policy or practice decisions. | Introduction section, last paragraph in particular. |
| **Methods** | | | |
| Health economic analysis plan | 4 | Indicate whether a health economic analysis plan was developed and where available. | No previous protocol was published. |
| Study population | 5 | Describe characteristics of the study population (such as age range, demographics, socioeconomic, or clinical characteristics). | The population is described in the first and second paragraphs of the methods. |
| Settings and location | 6 | Provide relevant contextual information that may influence findings. | First subsection of the methods, "Settings and the historical control of gHAT". Illustrated in figure A in S1 Text. |
| Comparators | 7 | Describe the interventions or strategies being compared and state why they were chosen. | Second subsection of the methods, "Strategies", Table 1 and Figure 2. Supplemental section: S1.2.2. |
| Perspective | 8 | State the perspective(s) adopted by the study and the rationale. | Fifth subsection of the methods, "Costs". |
| Time horizon | 9 | State the time horizon over and why the horizon is appropriate. | Sixth subsection of the methods "Cost-effectiveness analysis", first paragraph. |
| Discount rate | 10 | Report the discount rate(s) and reason chosen. | Sixth subsection of the methods "Cost-effectiveness analysis", third paragraph. 3% is the recommended rate by WHO-CHOICE and the Gates Reference Case. |
| Selection of outcomes | 11 | Describe what outcomes were used as the measure(s) of benefit(s) and harm(s). | Fourth subsection of the methods, "Outcome metrics". Supplemental sections S1.2.3-S1.2.4 contain detailed explanations of how DALYs are calculated and how the natural history of HAT was considered. |

*(continued)*

| Section/item | Item No | Recommendation | Reported on page no, line no |
|---|---|---|---|
| Measurement of outcomes | 12 | Describe how outcomes used to capture benefit(s) and harm(s) were measured. | Outcomes were not measured, but were simulated. See the fourth subsection of the methods, "Outcome metrics". Effectiveness of AS, PS and VC strategies: model-based, of treatment: the literature. Described in detail in sections E, F, and S4 Text. Supplemental sections S1.2.3-S1.2.4 contain detailed explanations of how DALYs are calculated and how the natural history of HAT was considered. |
| Measurement and valuation of resources and costs | 12 | If applicable, describe the population and methods used to elicit preferences for outcomes. | The construction of program costs are detailed in section S1.3, in pages S1.9-S1.20. As can be seen, in a publication of this scope, detailing the cost inputs in the main body would be unfeasible. However, the resulting expected costs are in Tables 3 & 4 of the main body of the paper. |
| Currency, price date, and conversion | 15 | Report the dates of the estimated resource quantities and unit costs, plus the currency and year of conversion. | Our general approach for this is in the section "Principles for parameterization" in S4 Text in page S4.3, section S4.2, followed by the specific choices for each parameter. |
| Rationale and description of model | 16 | If modelling is used, describe in detail and why used. Report if the model is publicly available and where it can be accessed. | The decision analytic model is illustrated in Figure B, but the components models feeding into the decision analytic model are described as follows: the dynamic transmission (SEIRS) model is described briefly in the third methods section and section S1.2.1 as well as pictured in Figure C part A, and the treatment model is described briefly in the third methods section and shown in Figure C part B and described in Section S1.2.3. |
| Analytics & assumptions | 17 | Describe any methods for analysing or statistically transforming data, any extrapolation methods, and approaches for validating any model used. | The transmission and treatment models are discussed in the third methods subsection "Transmission and treatment models". Section S1.2 in S1 Text and the parameter glossary in S4 Text. |
| Characterising heterogeneity | 18 | Describe any methods used for estimating how the results of the study vary for subgroups. | There were no subgroups in the study. It was a disease ecology study coupled with an economic evaluation that looked at the impact of interventions on the transmission dynamics - which are inherently group-level metrics. |
| Characterising distributional effects | 19 | Describe how impacts are distributed across different individuals or adjustments made to reflect priority populations. | Because there were no subgroups, there were no distributional effects. The differential impacts of treatment on poorer or less poor individuals were beyond the scope of this paper. |

*(continued)*

| Section/item | Item No | Recommendation | Reported on page no, line no |
|---|---|---|---|
| Characterising uncertainty | 20 | Describe methods to characterise any sources of uncertainty in the analysis. | The epidemiological parameters are the posterior distributions of a model fitted to time-series data, and full details are available in another publication (Rock *et al* [1]). For the parameters to model health outcomes and costs, assumptions and estimates were parameterized according to conventions in the economic evaluation literature [2], taking care to sample from large distributions for aspects for which we knew very little. Epidemiological parameters are available on OSF `https://osf.io/rak9d/`. Health outcome and cost-effectiveness parameters: see Table 2 and Supplementary information S4 Text |
| Approach to engagement with patients and others affected by the study | 21 | Describe any approaches to engage patients or service recipients, the general public, communities, or stakeholders (such as clinicians or payers) in the others affected by the study. | Strategy components were determined along with the country director of PNLTHA, Peka Mallaye and other PNLTHA members, Justin Darnas and Severin Mbainda (co-authors). Implementation of simulations of AS and PS costs were aided by information from Paul Bessell, who has helped maintain the data of surveillance activities by FIND (Foundation for Innovative & Novel Diagnostics) and collaborators at the Liverpool School of Tropical Medicine, who have run field operations in Chad since 2014. |
| **Results** | | | |
| Study parameters | 22 | Report the values, ranges, references, and, if used, probability distributions for all parameters. Report reasons or sources for distributions used to represent uncertainty where appropriate. Providing a table to show the input values is strongly recommended. | Table 2 and described in more detail in S4 Text. |
| Incremental costs and outcomes | 23 | For each intervention, report mean values for the main categories of estimated costs and outcomes of interest, as well as mean differences between the comparator groups. If applicable, report incremental cost-effectiveness ratios. | Table 3 & 4 in the main body, and Table B for the sensitivity analysis in S3 Text. |

*(continued)*

| Section/item | Item No | Recommendation | Reported on page no, line no |
|---|---|---|---|
| Effect of uncertainty | 24 | Model-based economic evaluation: Describe the effects on the results of uncertainty for all input parameters, and uncertainty related to the structure of the model and assumptions. | Tables 3 & 4 in the main body, and Table B for the sensitivity analysis in S3 Text. In addition, cost-effectiveness acceptability curves are depicted in supplementary figures 4 in the main body and E in S3 Text for alternative horizons and with and without discounting. Then, an analysis of expected value of partial perfect information, shown in Fig F, shows the parameters that drove the uncertainty. The interpretation is in the results section and the discussion. |
| Effect of engagement with patients and others affected by the study | 25 | Report on any difference patient/service recipient, general public, community, or stakeholder involvement made to the approach or findings of the study | Our engagement with the stakeholders (co-authors) was iterative throughout the process. There are no result interpretations that were not affected by their involvement. |
| **Discussion** | | | |
| Study findings, limitations, generalisability, and current knowledge | 26 | Summarise key study findings and describe how they support the conclusions reached. Discuss limitations and the generalisability of the findings and how the findings fit with current knowledge. | Discussion. |
| **Other relevant information** | | | |
| Source of funding | 27 | Describe how the study was funded and the role of the funder in the identification, design, conduct, and reporting of the analysis. Describe other non-monetary sources of support. | Funding statement. |
| Conflicts of interest | 28 | Describe any potential for conflict of interest of study contributors in accordance with journal policy. In the absence of a journal policy, we recommend authors comply with International Committee of Medical Journal Editors recommendations. | Conflict of interest statement. |
